# The neurology and neuropsychiatry of COVID-19: a systematic review and meta-analysis of the early literature reveals frequent CNS manifestations and key emerging narratives

**DOI:** 10.1101/2021.02.24.21252335

**Authors:** Jonathan P Rogers, Cameron Watson, James Badenoch, Benjamin Cross, Matthew Butler, Jia Song, Danish Hafeez, Hamilton Morrin, Emma Rachel Rengasamy, Lucretia Thomas, Silviya Ralovska, Abigail Smakowski, Ritika Dilip Sundaram, Camille Kaitlyn Hunt, Mao Fong Lim, Daruj Aniwattanapong, Vanshika Singh, Zain Hussain, Stuti Chakraborty, Ella Burchill, Katrin Jansen, Heinz Holling, Dean Walton, Thomas A Pollak, Mark Ellul, Ivan Koychev, Tom Solomon, Benedict Daniel Michael, Timothy R Nicholson, Alasdair G Rooney

## Abstract

**Objectives:** There is accumulating evidence of the neurological and neuropsychiatric features of infection with SARS-CoV-2. In this systematic review and meta-analysis, we aimed to describe the characteristics of the early literature and estimate point prevalences for neurological and neuropsychiatric manifestations.

**Methods:** We searched MEDLINE, Embase, PsycInfo and CINAHL up to 18 July 2020 for randomised controlled trials, cohort studies, case-control studies, cross-sectional studies and case series. Studies reporting prevalences of neurological or neuropsychiatric symptoms were synthesised into meta-analyses to estimate pooled prevalence.

**Results:** 13,292 records were screened by at least two authors to identify 215 included studies, of which there were 37 cohort studies, 15 case-control studies, 80 cross-sectional studies and 83 case series from 30 countries. 147 studies were included in the meta-analysis. The symptoms with the highest prevalence were anosmia (43.1% [35.2—51.3], *n*=15,975, 63 studies), weakness (40.0% [27.9—53.5], *n*=221, 3 studies), fatigue (37.8% [31.6—44.4], *n*=21,101, 67 studies), dysgeusia (37.2% [30.0—45.3], *n*=13,686, 52 studies), myalgia (25.1% [19.8—31.3], *n*=66.268, 76 studies), depression (23.0 % [11.8—40.2], *n*=43,128, 10 studies), headache (20.7% [95% CI 16.1—26.1], *n*=64,613, 84 studies), anxiety (15.9% [5.6—37.7], *n*=42,566, 9 studies) and altered mental status (8.2% [4.4—14.8], *n*=49,326, 19 studies). Heterogeneity for most clinical manifestations was high.

**Conclusions:** Neurological and neuropsychiatric symptoms of COVID-19 in the pandemic’s early phase are varied and common. The neurological and psychiatric academic communities should develop systems to facilitate high-quality methodologies, including more rapid examination of the longitudinal course of neuropsychiatric complications of newly emerging diseases and their relationship to neuroimaging and inflammatory biomarkers.

## INTRODUCTION

COVID-19 stimulated a global academic response to examine the clinical sequelae and biology of the SARS-CoV-2 virus, including its neurological and neuropsychiatric impact. [1,2] Although the earliest reports naturally highlighted respiratory symptoms, [1] it was quickly recognised that SARS-CoV-2, like other coronaviruses, [2] can affect the central and peripheral nervous system. [3,4]

Many of the very earliest studies of the neurological and neuropsychiatric complications of SARS-CoV-2 infection were small retrospective case reports or series. [7,8] These initial studies were feasible to deliver quickly in the context of a new and poorly understood disease. Case reports [5,6] were superseded by case series [7,8], then case-control [9] and cohort studies [10,11], which suggested significant morbidity and mortality from neurological or neuropsychiatric complications. [12] Currently, large multi-centre prospective studies are underway [13] and already reporting. [14] We anticipate that the quality of evidence, and our knowledge, will improve considerably as these data continue to emerge rapidly.

In response to these signals, we aimed to develop a novel, sustainable platform to evaluate emerging knowledge of the neurology and neuropsychiatry of COVID-19. This also served to assist colleagues in keeping up to date with the literature relevant to their specialty, given the extraordinary volume and pace with which research is being published. In May 2020 we started logging literature on relevant symptoms, clinical associations, and putative underlying mechanisms in our blog, “The neurology and neuropsychiatry of COVID-19”, published weekly on the *Journal of Neurology, Neurosurgery and Psychiatry* website. [15] This catalogue of observational studies, reviews, editorials, and mechanistic studies has had over 27,000 global views, but it is essentially a library in which studies are narratively summarised and filed. We recognised the potential value of extending this platform to enable analytic summaries by synthesising evidence in the form of a systematic review and meta-analysis, which we termed **<u>S</u>**ystematically **<u>A</u>**nalyse and **<u>R</u>**eview **<u>S</u>**tudies of **<u>COV</u>**ID-19 **<u>Neuro</u>**logy and neuropsychiatry (**SARS-COV-Neuro**).

In the current report we aimed to answer two questions:

1. What were the key methodological characteristics of the early evolving literature on the neurological and neuropsychiatric consequences of COVID-19?
2. What was the prevalence of neurological and neuropsychiatric complications in COVID-19 patients in observational or interventional studies during this early period of evolving knowledge?

This review is the most comprehensive attempt yet to synthesise the data on the neurological and neuropsychiatric consequences of COVID-19. Other previous works are less up-to-date, incorporate fewer clinical parameters or have limited scope for meta-analysis. [2,16–19]

## METHODS

We conducted a systematic review and meta-analysis, based on a registered protocol (PROSPERO ID CRD42020200768) and reported according to PRISMA guidelines [20] (see Supplementary Table 1 for completed PRISMA checklist). A full list of author contributions is provided in Supplementary Methods 2.

The overall strategy was to combine synonyms for COVID-19 infection with synonyms for neurological and neuropsychiatric syndromes. We searched Ovid MEDLINE(R) and Epub Ahead of Print, In-Process & Other Non-Indexed Citations and Daily, EMBASE (via Ovid), APA PsycInfo (via OVID) and CINAHL (via EBSCO) from 1st January 2020 to 18th July 2020. Reference lists of other systematic reviews were examined and cross-checked against our database and eligibility criteria. The full search strategy is presented in supplementary methods 1.

We included any controlled trials, cross-sectional, case-control, cohort studies or case series reporting neuropsychiatric or neurological manifestations in confirmed or clinically suspected COVID-19 patients. We excluded non-English language reports. We excluded studies reporting on fewer than 10 infected patients to avoid the reporting biases common in small studies. Meta-analysis was conducted where a clinical manifestation was reported by three or more eligible studies. Studies were included in the meta-analysis only where they provided representative samples of patients with COVID-19 in whom the point prevalence of neurological or neuropsychiatric features could be estimated; studies where patient inclusion was based on neurological or neuropsychiatric complications (e.g. only those referred for clinical neuroimaging) were therefore excluded from the meta-analysis.

Screening of titles, abstracts and full texts for each article was conducted by two of the authors (CW, JS, AGR, BC, MB, DH, JB, ER), each blinded to the other’s ratings. Where there was disagreement about study inclusion, a third author who was a senior member of the team (AGR, MB, JS or JPR) arbitrated. Zotero was used for reference management and Rayyan QCRI was used for eligibility screening.

Data extraction was performed onto structured forms by two authors: one of the authors (ER, DH, BC, CW, HM, JB) entered the data, then a second author (AGR, MB, CH, AS, JB, JS, BC, ER, HM, DA, SR or MFL) checked the data. We recorded the methodological characteristics of studies and the frequency of neurological and neuropsychiatric manifestations reported by each study (see full list of variables extracted in Supplementary table 2). Where studies reported asthenia as a manifestation, this was coded as fatigue; where a paper reported both asthenia and fatigue, only the figures for fatigue were used.

Levels of evidence were assessed by use of the Oxford Centre for Evidence-based Medicine Levels of Evidence. [21] Quality of studies and risk of bias were assessed using the Newcastle-Ottawa Scale, including its adaptation for cross-sectional studies. [22,23] Quality was assessed by two authors in parallel with arbitration by a third author in cases of disagreement.

For the systematic review, we descriptively reported methodological characteristics of the evolving literature with analytic statistical tests where appropriate. All eligible studies were listed in a table with their study design, demographics and main findings.

For the meta-analysis, the primary outcome was point prevalence of neurological and neuropsychiatric manifestations with 95% confidence intervals. Given the potential for estimation errors with a double-arcsine transformation of proportion, [24] we used the *metafor* package in *R* version 4.0.2 to calculate generalised linear mixed models (GLMMs) for each outcome, [25,26] before then using the double arcsine transformation as a comparative sensitivity analysis. [27,28] Outcome proportions were transformed using a logit transformation. Between-study heterogeneity was calculated using the *I*^2^ statistic. We planned *a priori* to analyse the following subgroups: retrospective or prospective design, method of SARS-CoV-2 diagnosis, severity of COVID-19, and time-point in relation to infection. Ultimately, we only conducted subgroup analysis for retrospective or prospective design and severity of COVID-19 because of lack of consistently presented data for the other subgroups. In addition, due to high heterogeneity, we conducted an additional exploratory subgroup analysis examining country of origin. Subgroup analyses were conducted on the five clinical manifestations most commonly studied: anosmia, dysgeusia, fatigue, myalgia, and headache. Significance testing was performed to assess differences in reported frequencies by sub-group.

## RESULTS

De-duplicated searches returned a total of 13,292 titles. Abstract and full text screening generated a final list of 215 eligible studies (Figure 1). A complete list of all included studies is presented in Supplementary Table 3.

**Figure 1:**
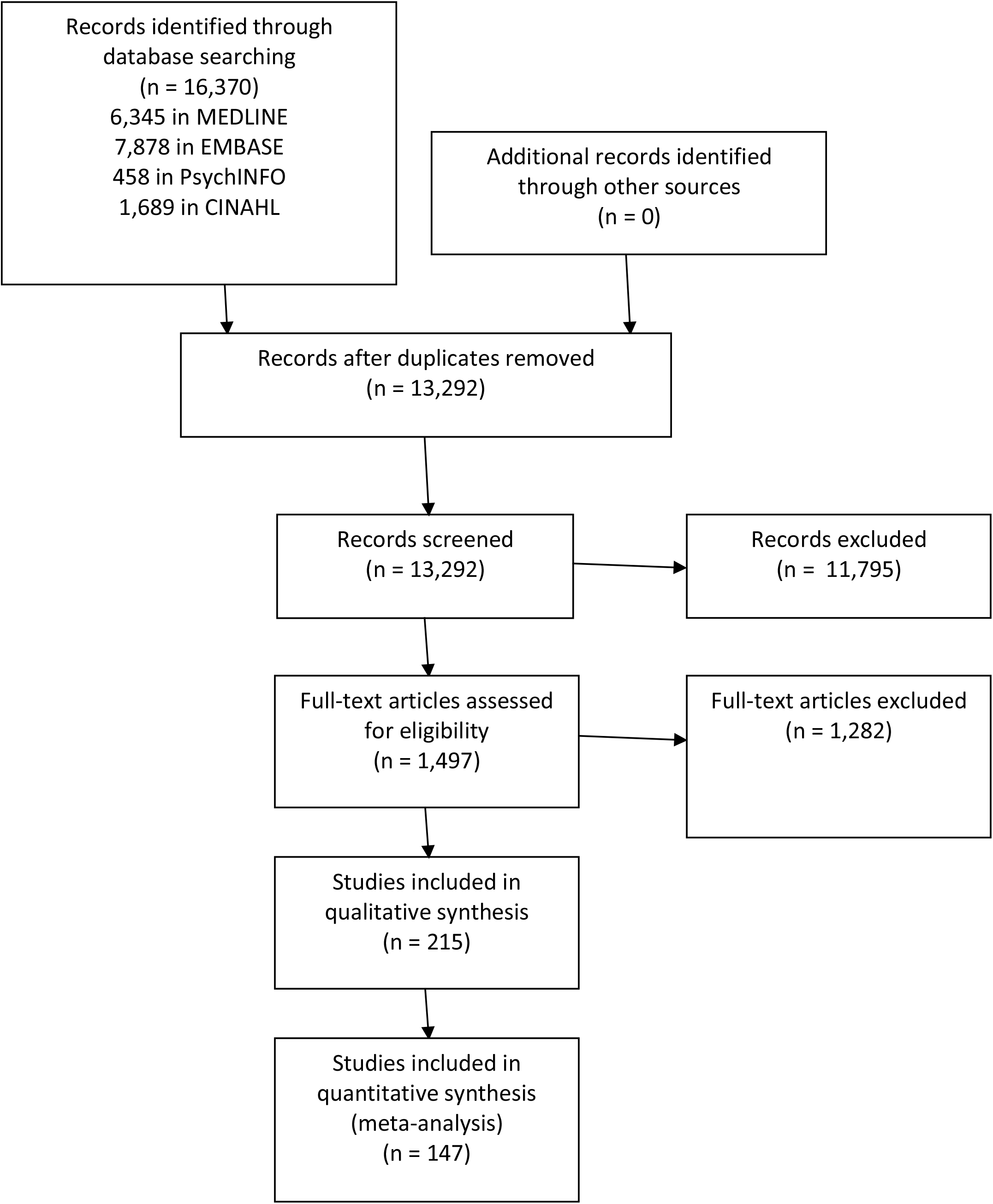
PRISMA flow diagram.

### Methodological characteristics of the literature

Methodological characteristics of the studies are summarised in Table 1. The most common study type was a case series (83 studies, 38.6%). To explore whether designs evolved in the first half of 2020, we considered studies that started data collection in December 2019 to February 2020 to be earlier and those between March and July 2020 to be later. Among the earlier studies, 37 out of 65 (57%) were case series, whereas this proportion fell to 40 out of 115 (34.8%) among the subsequent studies, *p*=0.004. Change in study design is illustrated in Figure 2. Overall, therefore, there was at least a two-month lag period from the first official report of the outbreak in Wuhan by the Chinese authorities (31st Dec 2019) to the first group of cohort studies.

**Table 1:**
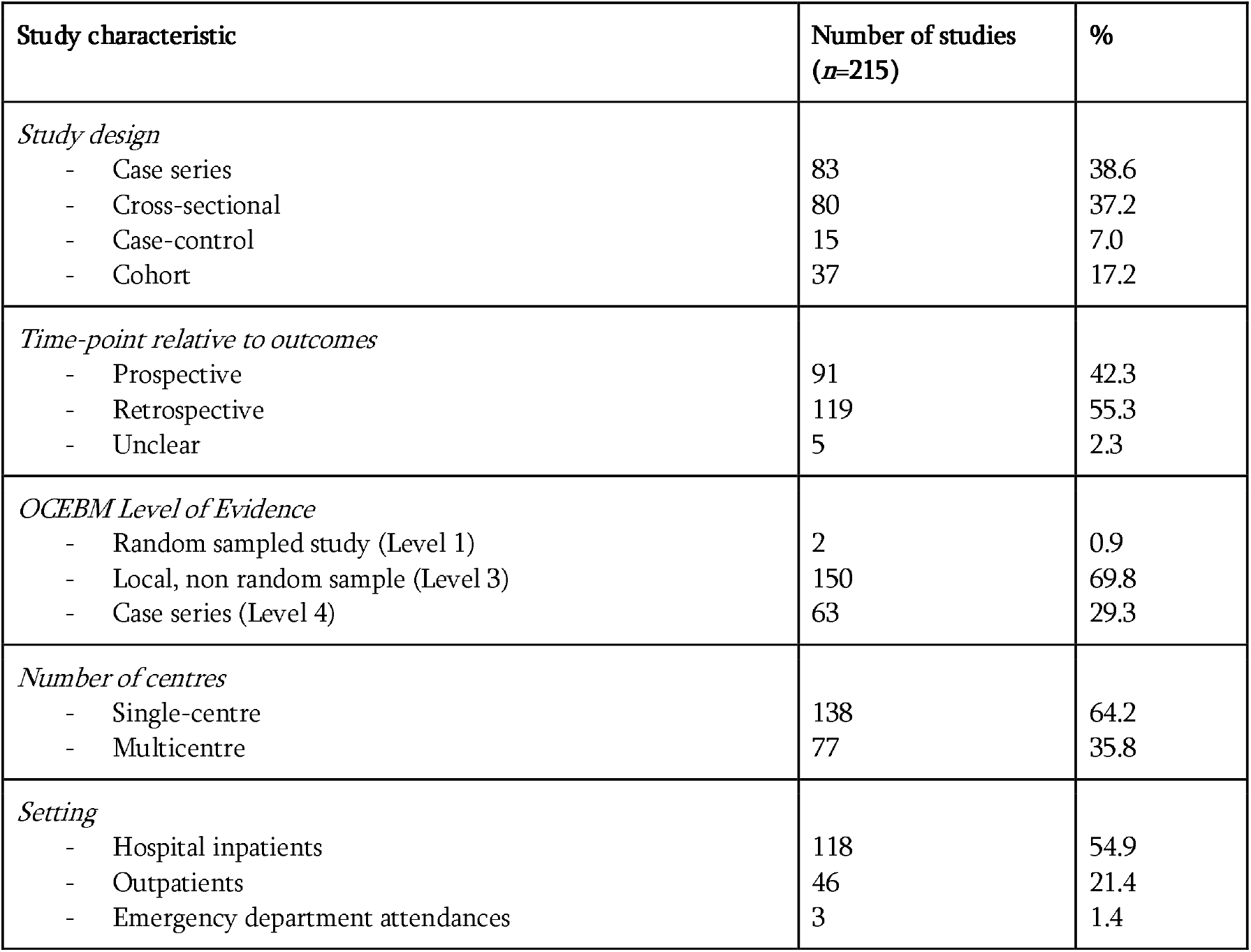
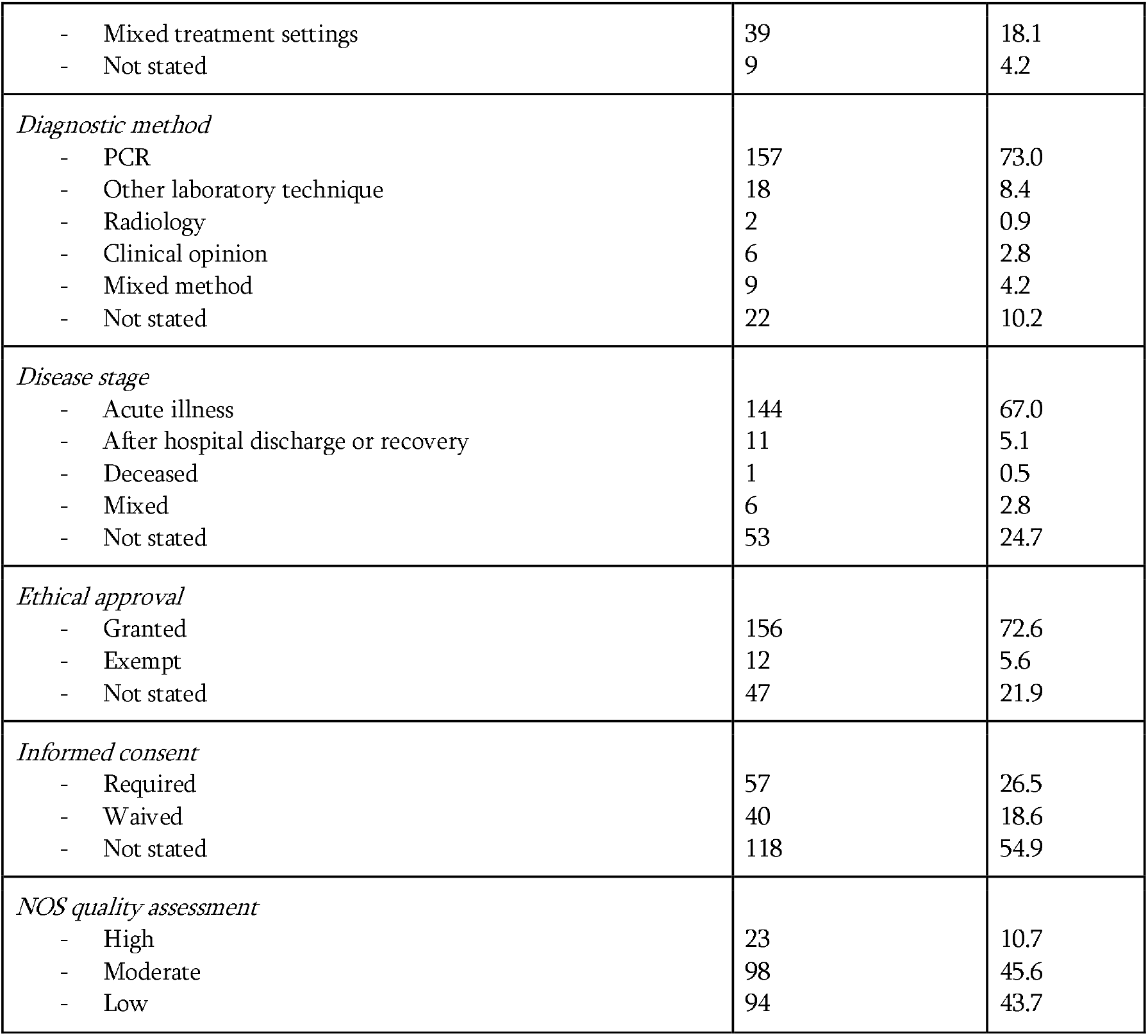
Methodological characteristics of included studies

**Figure 2:**
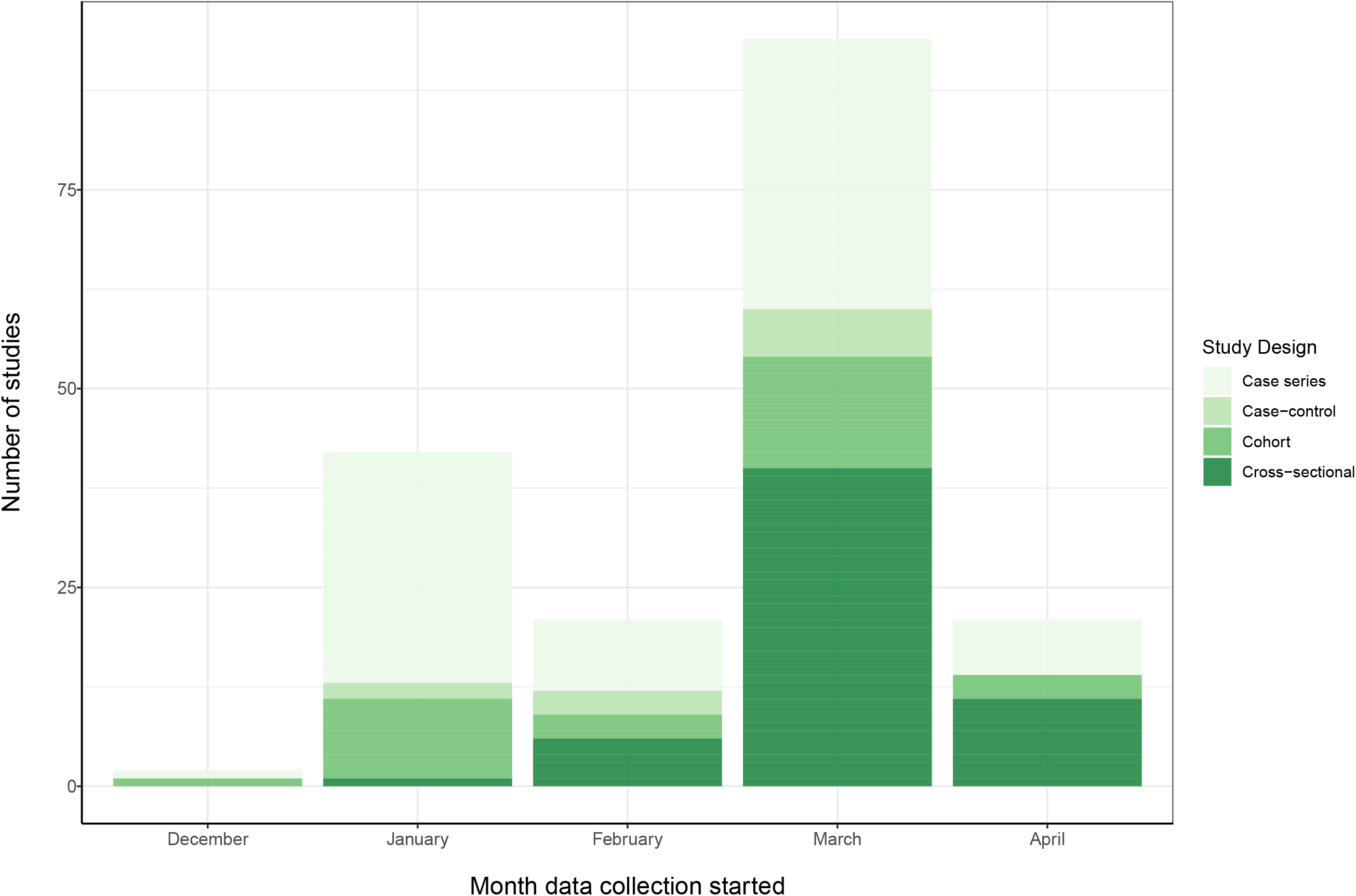
Study design trends.

Studies were written by a primary author affiliated with an institution from a total of 30 countries globally (Figure 3). The most frequent contributors were China *(n*=50 studies), USA (*n*=32 studies), Italy (*n*=28 studies), and France *(n*=23 studies). All but three studies starting recruitment in Jan 2020 were located in China. Globally, most studies (138, 64.2%) were single-centre without a significant shift towards multi-centre studies as the pandemic accelerated: where collection date was clear, 44/65 (67.7%) of earlier studies were single-centre, compared to 72/115 (62.6%) of late studies (*p*=0.49).

**Figure 3:**
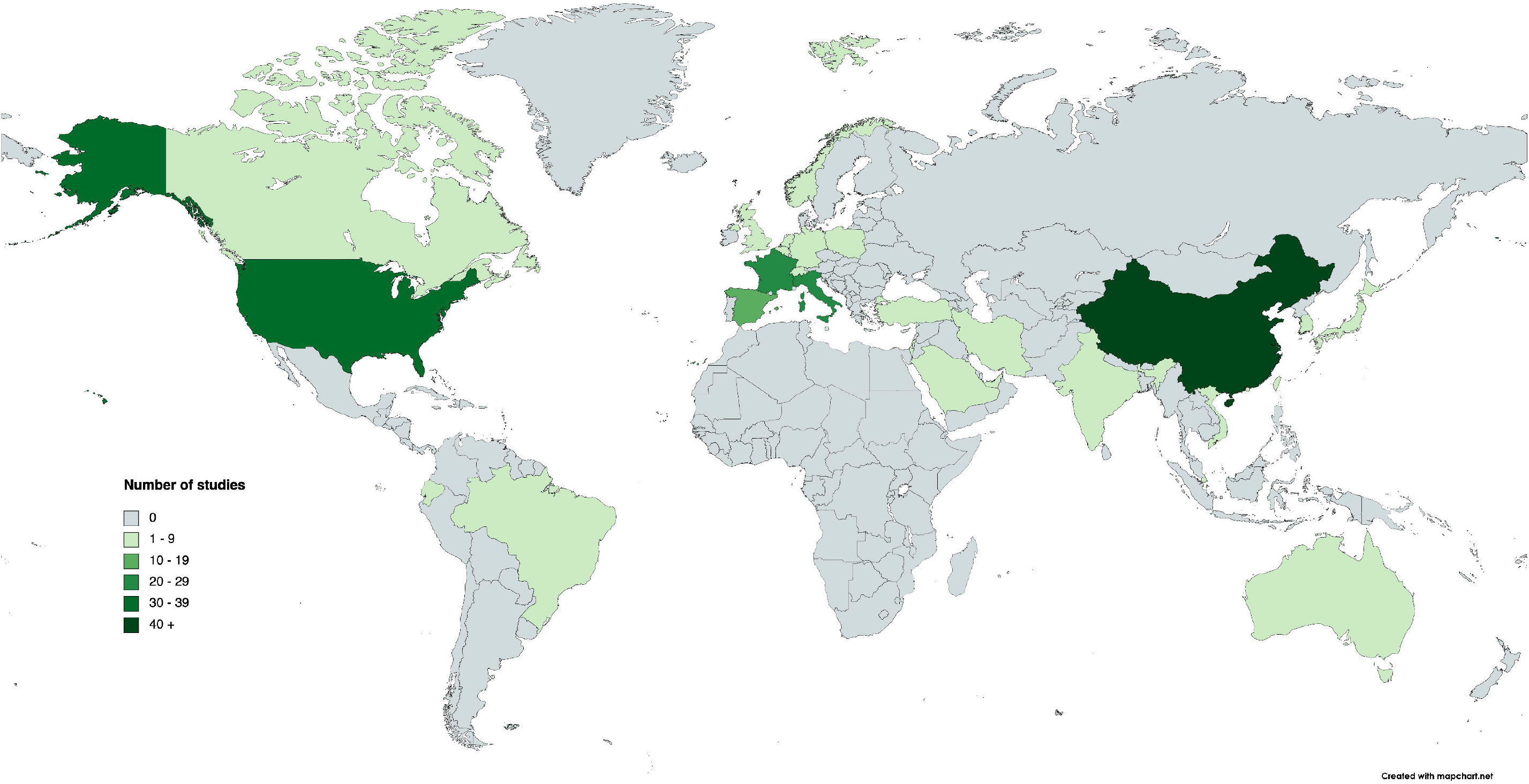
Geographical distribution of studies.

Studies were predominantly in hospitalised patients (118 studies, 54.9%) and during the acute illness (144, 67.0%). There were a total of 105,638 subjects. Number of subjects in each study varied between 10 and 40,469 (median 101, IQR 196). There were 18 studies with 1000 or more subjects.

There was evidence for ethical approval and informed consent in most studies, but this was waived in a minority, frequently because of the particular circumstances of the pandemic.

Quality assessment found only 23 (10.7%) studies to be of high quality, 98 (45.6%) were of moderate quality and 94 (43.7%) were of low quality.

### Prevalence of neuropsychiatric and neurological manifestations

Twenty neurological or neuropsychiatric manifestations were estimated by at least 3 studies, such that we included 147 studies (reporting on 99,905 infected patients) in the meta-analysis. Overall prevalences are shown in Table 2 with forest plots available in Figures 4-8 and Supplementary Figures 1-20. The most *often-studied* symptoms were headache (examined in 84 studies, *n*=64,613), myalgia (76 studies, *n*=66,268), fatigue (67 studies, n=21,101), anosmia (63 studies, 15,975) and dysgeusia (52 studies, *n*=13,686). The most *prevalent* symptoms were anosmia (43.1% [35.2-51.3], *n*=15,975 in 63 studies), weakness (40.0% [27.9-53.5], *n*=221 in 3 studies), fatigue (37.8% [31.6-44.4], *n*=21,101 in 67 studies) dysgeusia (37.2% [29.8-45.3], *n*=13,686 in 52 studies) and myalgia (25.1% [19.8-31.3], *n*=66,268 in 76 studies). Between-study heterogeneity was mostly high with *I*^2^≥90% for 13 manifestations, ≥50% and <90% for 2 manifestations and <50% for 5 manifestations. Most symptoms were recorded merely as ‘present’ or ‘absent’ by study authors. The robustness of the main analyses were assessed by repeating the analyses on headache, myalgia, anosmia, fatigue and dysgeusia using the standard random-effects model for meta-analysis with the Freeman-Tukey double arcsine transformation. The results were in line with the main analysis (see Supplementary table 4).

**Table 2:** Overall meta-analytic estimates of point prevalence of neurological or neuropsychiatric symptoms

| Symptom/Syndrome | Studies | *n* | Point prevalence (%) | 95% CI | *P* |
| --- | --- | --- | --- | --- | --- |
| Headache | 84 | 64,613 | 20.7 | 16.1 - 26.1 | 99.0% |
| Myalgia | 76 | 66,268 | 25.1 | 19.8-31.3 | 99.1% |
| Fatigue | 67 | 21,101 | 37.8 | 31.6 – 44.4 | 98.7% |
| Anosmia | 63 | 15,975 | 43.1 | 35.2 – 51.3 | 98.8% |
| Dysgeusia | 52 | 13,686 | 37.2 | 29.8 – 45.3 | 98.6% |
| Dizziness/vertigo | 26 | 47,619 | 6.4 | 4.0 - 10.0 | 97.1% |
| Altered mental status | 19 | 49,326 | 8.2 | 4.4 - 14.8 | 99.0% |
| Anosmia at follow-up | 11 | 3,182 | 11.8 | 5.5 – 23.5 | 98.5% |
| Depression | 10 | 43,128 | 23.0 | 11.8 - 40.2 | 99.3% |
| Anxiety | 9 | 42,566 | 15.9 | 5.6 - 37.7 | 99.5% |
| Sleep disorder | 8 | 42,221 | 23.5 | 12.0 - 40.9 | 98.9% |
| Ischaemic stroke | 8 | 5,258 | 1.9 | 1.3 – 2.8 | 61.7% |
| Other CVD | 6 | 43,701 | 1.6 | 0.3 – 7.9 | 98.7% |
| Dysgeusia at follow-up | 6 | 2,065 | 11.7 | 5.1– 25.0 | 96.7% |
| Seizure | 5 | 41,929 | 0.06 | 0.06 – 0.07 | 0.0% |
| Haemorrhagic stroke | 5 | 3,074 | 0.4 | 0.3 - 0.7 | 0.0% |
| Visual defect | 5 | 678 | 3.0 | 1.9 - 4.5 | 0.0% |
| Hearing impairment | 4 | 557 | 2.0 | 1.1 – 3.5 | 0.0% |
| Tinnitus | 4 | 455 | 3.5 | 1.7 – 7.4 | 51.8% |
| Weakness | 3 | 221 | 40.0 | 27.9 – 53.5 | 45.4% |

**Figure 4:**
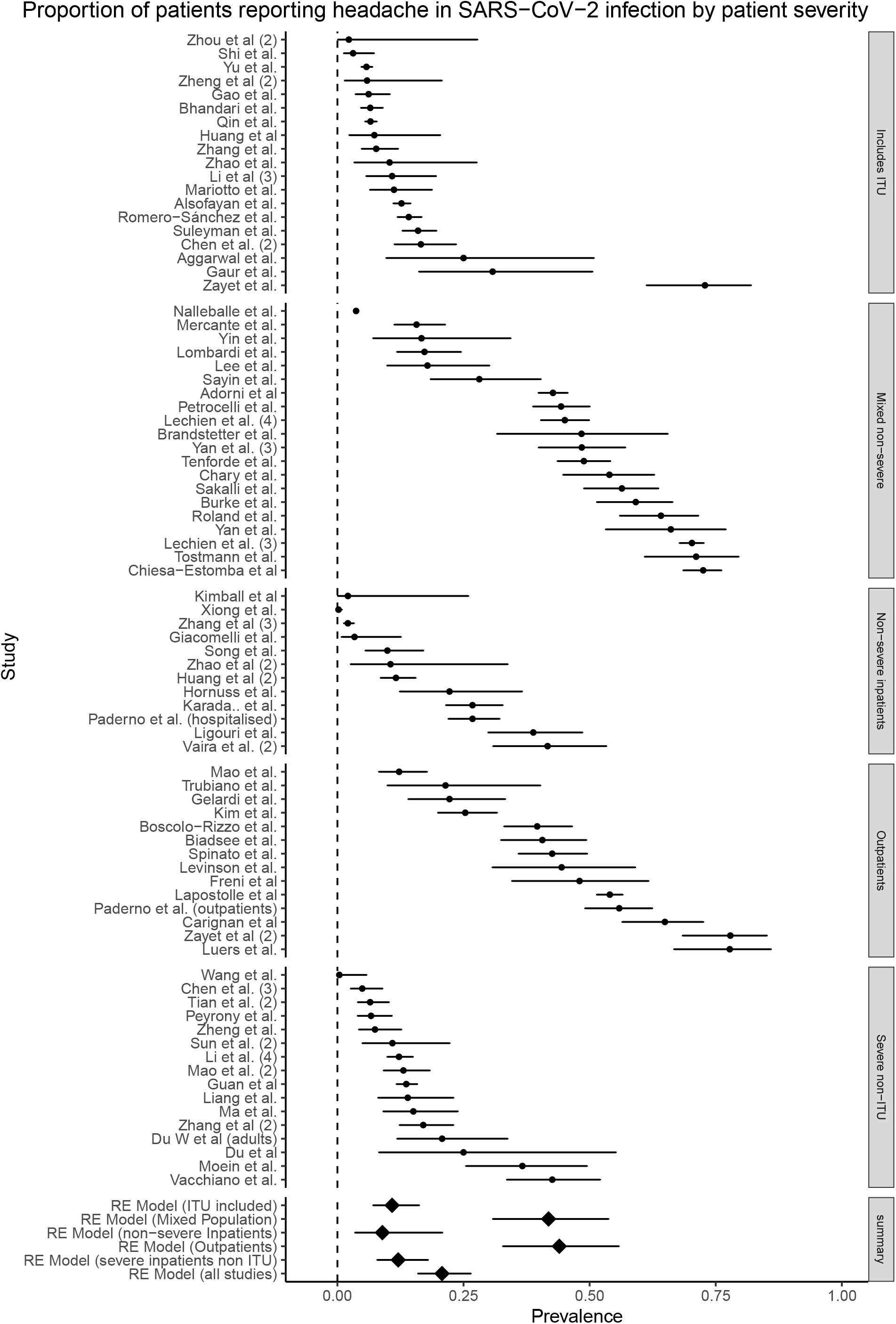
Forest plot for headache subgrouped by disease severity.

**Figure 5:**
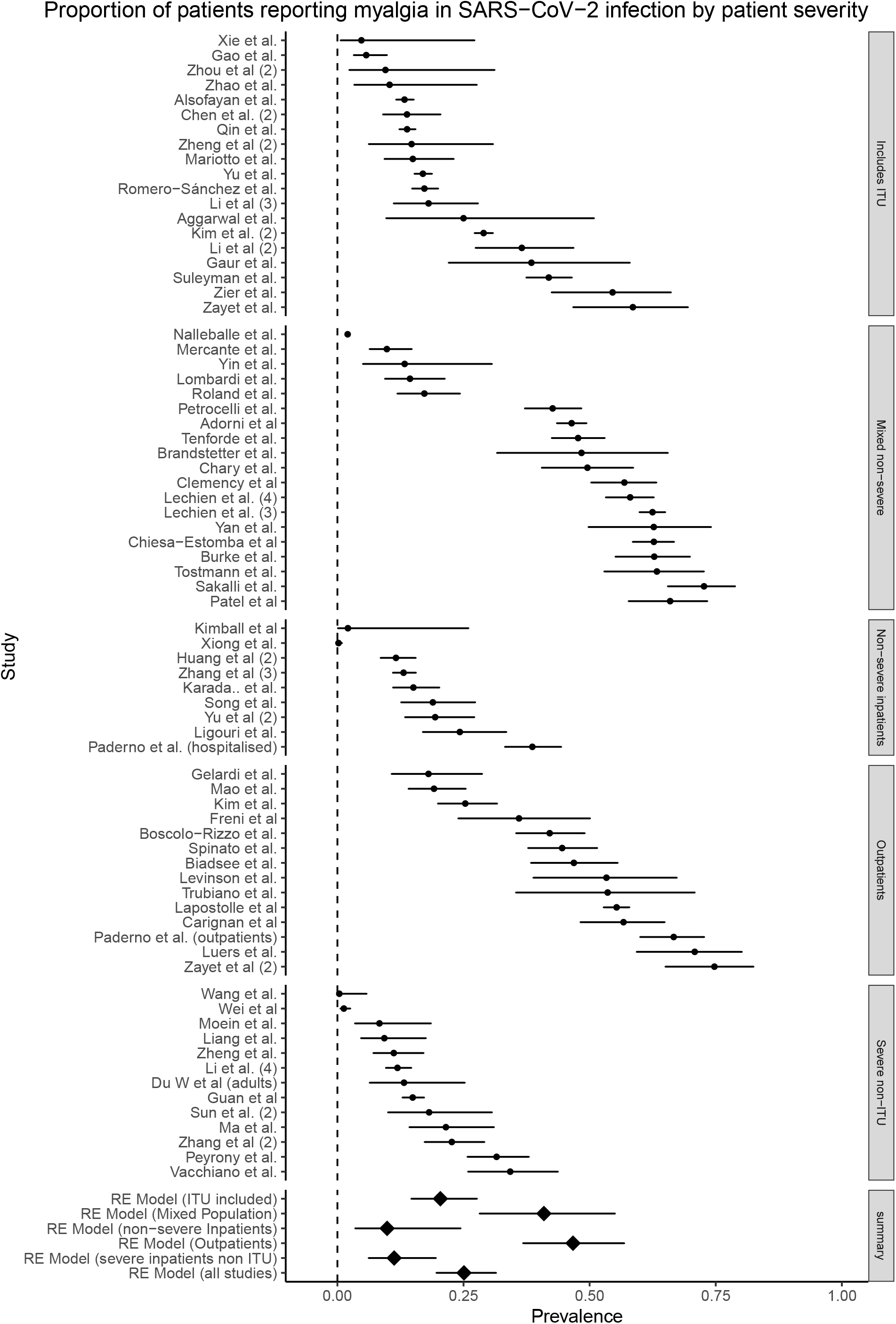
Forest plot for myalgia subgrouped by disease severity.

**Figure 6:**
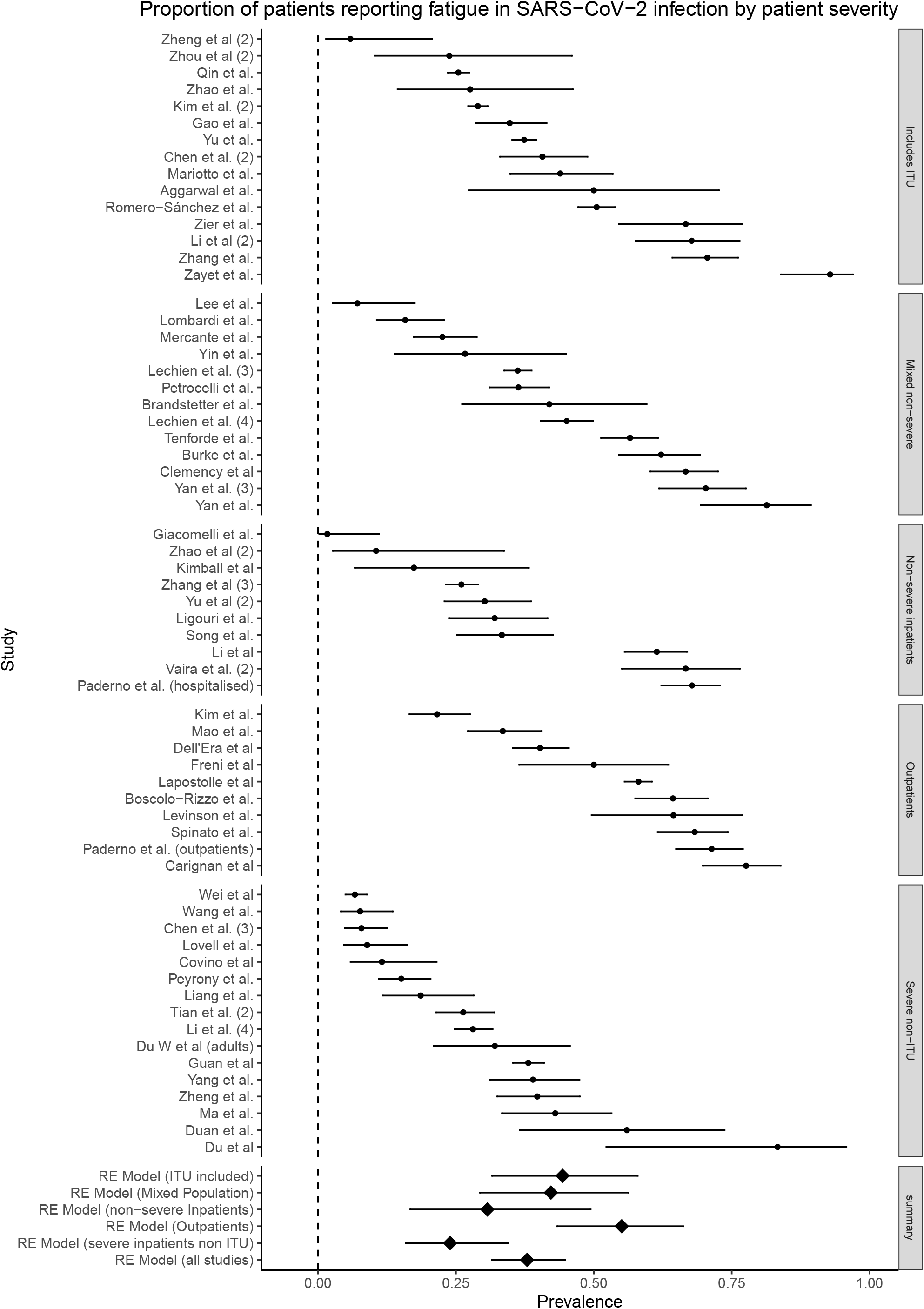
Forest plot for fatigue subgrouped by disease severity.

**Figure 7:**
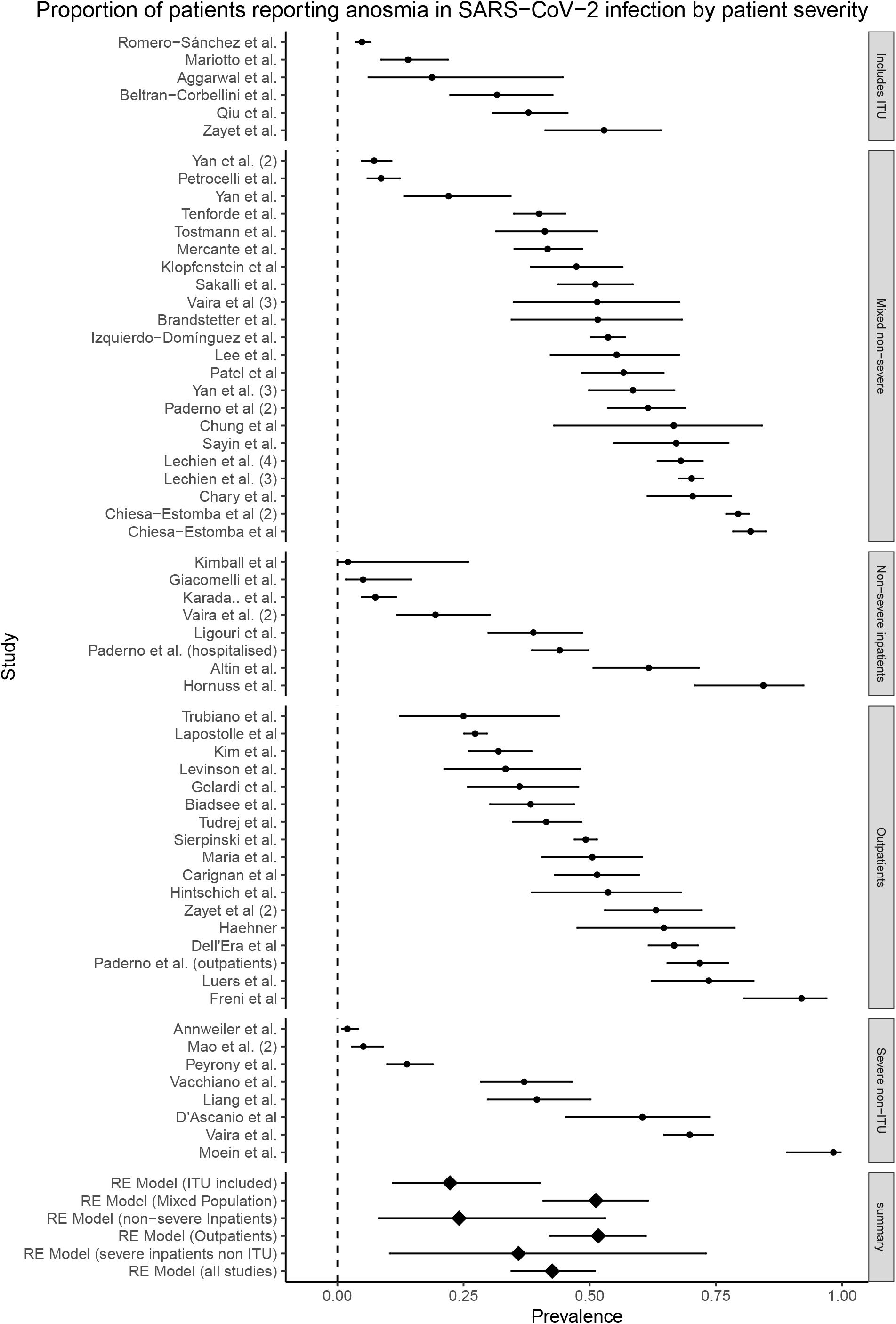
Forest plot for anosmia subgrouped by disease severity.

**Figure 8:**
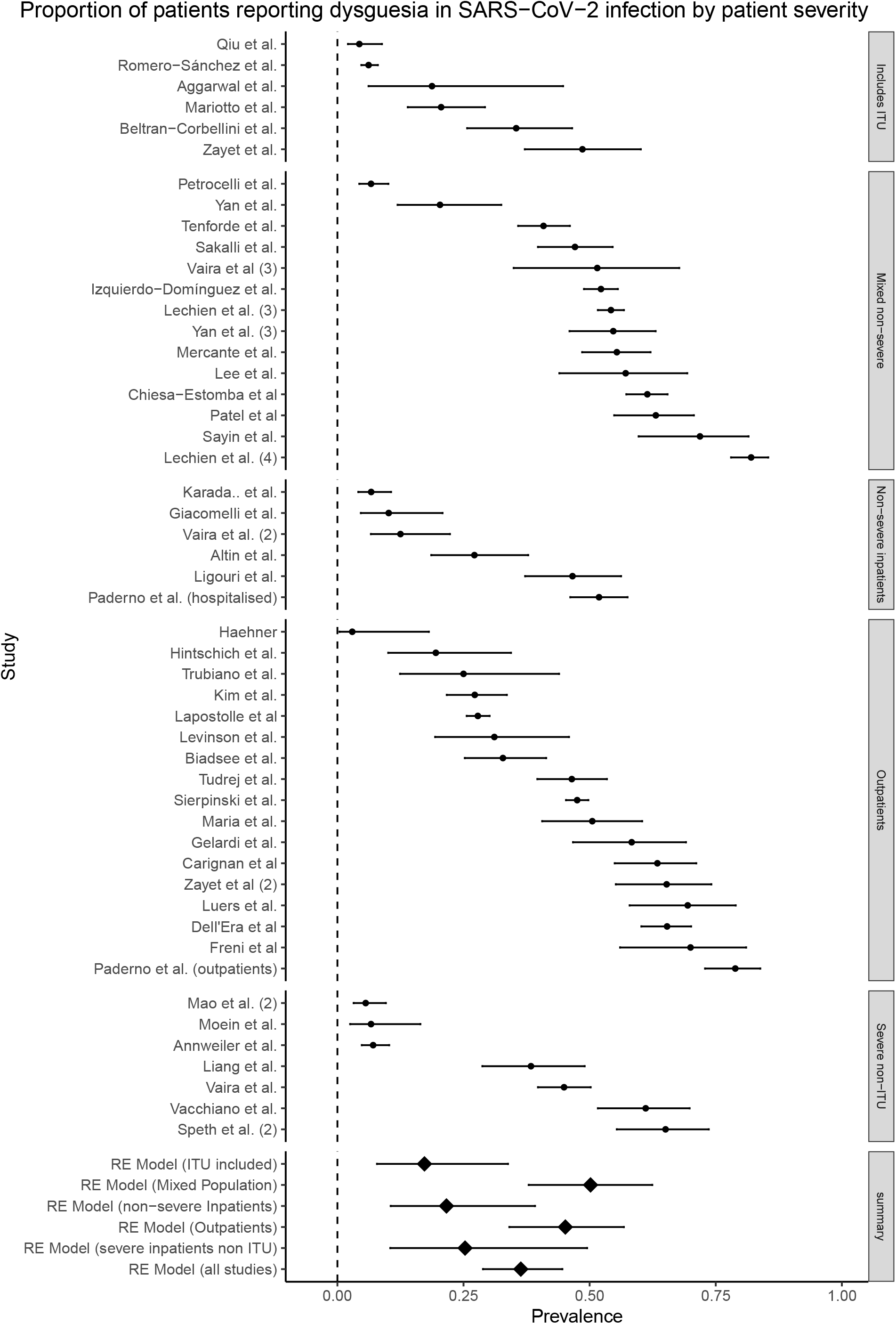
Forest plot for dysgeusia subgrouped by disease severity.

#### Subgroup analyses

Subgroup analysis was conducted by study design (prospective and retrospective; Table 3), case severity (outpatient, mixed non-severe, non-severe inpatients, severe but not admitted to ITU and admitted to ITU; Table 4) and country of origin (Supplementary Table 5). For headache, myalgia, anosmia and dysgeusia, there were significantly higher reported rates in prospective studies than in retrospective studies. In the severity subgroup analysis, compared to the ITU group, headache was more common in mixed non-severe and outpatient populations (*p*<0.001); myalgia was more common in mixed non-severe and outpatient populations (*p*=0.04 and <0.001 respectively); anosmia was more common in mixed non-severe and outpatient populations (*p*=0.05 and 0.04, respectively), and dysgeusia was more common in mixed non-severe populations (*p*=0.02); there were no significant differences between groups for fatigue.

**Table 3:** Subgroup analysis by study design for 5 most commonly studied clinical manifestations

| Manifestation | Retrospective |  |  | Prospective |  |  | *p* |
| --- | --- | --- | --- | --- | --- | --- | --- |
|  | Studies | Prevalence (95% CI) | *P* | Studies | Prevalence (95% CI) | *P* |  |
| Headache | 46 | 11.1 (8.0-15.3) | 98.3 | 34 | 37.5 (29.3-46.5) | 98.2 | <0.001 |
| Myalgia | 42 | 16.8 (11.8-23.1) | 99.1 | 30 | 38.6 (29.6-48.5) | 98.5 | <0.001 |
| Anosmia | 16 | 22.3 (11.4-39.0) | 98.0 | 44 | 50.8 (42.5-59.1) | 98.6 | <0.005 |
| Dysgeusia | 13 | 22.3 (11.0-40.2) | 97.8 | 36 | 42.4 (34.4-50.9) | 98.5 | 0.04 |
| Fatigue | 41 | 33.5 (26.1-41.8) | 98.8 | 24 | 43.3 (33.2-54.1) | 98.5 | 0.14 |

**Table 4:** Subgroup analysis by case severity for 5 most commonly studied clinical manifestations

| Manifestat | Outpatients | Mixed non-severe | Non-severe inpatients | Severe non-ITU | Includes ITU |
| --- | --- | --- | --- | --- | --- |

| ion | Studi<br>es | Prevale<br>nce<br>(95%<br>CI) | *P* | Studi<br>es | Prevale<br>nce<br>(95%<br>CI) | *P* | Studi<br>es | Prevale<br>nce<br>(95%<br>CI) | *P* | Studi<br>es | Prevale<br>nce<br>(95%<br>CI) | *P* | Studi<br>es | Prevale<br>nce<br>(95%<br>CI) | *P* |
| --- | --- | --- | --- | --- | --- | --- | --- | --- | --- | --- | --- | --- | --- | --- | --- |
| Headache | 14 | 44.0<br>(32.8-55.8) | 96.<br>7 | 20 | 41.9<br>(30.9-53.7) | 99.<br>1 | 12 | 8.9 (3.5-20.8) | 97.<br>9 | 16 | 12.1<br>(7.9-17.9) | 94.<br>3 | 19 | 10.8<br>(7.1-16.2) | 96.<br>7 |
| Myalgia | 14 | 46.7<br>(36.8-56.8) | 95.<br>7 | 19 | 40.9<br>(28.2-55.0) | 99.<br>3 | 9 | 9.9 (3.6-24.4) | 98.<br>6 | 13 | 11.3<br>(6.2-19.5) | 97.<br>0 | 19 | 20.4<br>(14.7-27.6) | 97.<br>9 |
| Anosmia | 17 | 51.7<br>(42.2-61.1) | 97.<br>1 | 22 | 51.2<br>(40.8-61.4) | 98.<br>2 | 8 | 24.1<br>(8.2-53.1) | 98.<br>0 | 8 | 35.9<br>(10.4-73.0) | 99.<br>0 | 6 | 22.3<br>(11.0-40.1) | 95.<br>2 |
| Dysgeusia | 17 | 45.2<br>(34.1-56.8) | 98.<br>0 | 14 | 50.2<br>(37.9-62.4) | 98.<br>3 | 6 | 21.6<br>(10.6-39.2) | 95.<br>3 | 7 | 25.3(10.<br>5-49.5) | 97.<br>9 | 6 | 17.3<br>(7.8-33.8) | 94.<br>8 |
| Fatigue | 10 | 55.1<br>(43.3-66.3) | 97.<br>8 | 13 | 42.2<br>(29.3-56.3) | 98.<br>1 | 10 | 30.7(16.<br>7-49.4) | 98.<br>0 | 16 | 24.0<br>(15.9-34.4) | 97.<br>3 | 15 | 44.3<br>(31.5-58.0) | 99.<br>1 |

## DISCUSSION

To our knowledge, this is the largest and most comprehensive systematic review of the neurological and neuropsychiatric manifestations of COVID-19. We identified 215 studies, published between January and July 2020, with a total population of 105,638, containing a large variation in the size of studies. We uncovered some general findings about the methodological characteristics of early-evolving literature in response to a novel pathogen. Studies varied substantially in design, geographical location, treatment setting, illness stage, sample size, diagnostic method and clinical manifestations studied. More studies were retrospective than prospective and case series comprised a significant minority of the early literature. In terms of country of origin, after the first few weeks of the pandemic in which the literature was dominated by studies from China, a wide range of research was produced from 30 countries, among which less economically developed countries were mostly absent. Most studies confirmed formal ethical review and most required informed consent, but these requirements were waived in a subset of cases.

In our review we summarise point prevalence of 20 neurological and neuropsychiatric complications of COVID-19. The most frequently *studied* symptoms were heavily weighted towards non-specific features of systemic illness, such as headache, myalgia, fatigue, anosmia and dysgeusia. It was predominantly these more non-specific symptoms that were found to have the highest prevalences, ranging from 20.7% [16.1-26.1] to 43.1% [35.2-51.3] (headache and anosmia, respectively). Of note, more specific neurological and neuropsychiatric symptoms such as altered mental status, depression, anxiety, sleep disorder, stroke and seizures were less frequently studied. However, the core psychiatric disorders of depression (23.0% [11.8-40.2]) and anxiety (15.9% [5.6-37.7]) appeared to be highly prevalent. The reported prevalence of major neurological disorders such as ischaemic stroke (1.9% [1.3-2.8]), haemorrhagic stroke (0.4% [0.3-0.7%]) and seizure (0.06% [0.06-0.07]) were substantially lower. Subgroup analyses suggested that study design (prospective versus retrospective), severity of illness and country of origin of a study affected the prevalence figures obtained. Importantly, for myalgia, fatigue, anosmia and dysgeusia, prevalence rates were substantially higher in prospective studies compared to retrospective studies.

There are several limitations to our study, relating both to the quality of the underlying evidence and to the data synthesis. Major limitations in the study design were the frequent absence of comparison groups, limiting conclusions about the specificity of symptoms to COVID-19; retrospective study designs, which meant that only those symptoms that happened to be enquired about were included; and small sample sizes, which risk reporting bias. In terms of populations, the frequent use of hospital inpatients is unrepresentative of the majority of patients with COVID-19, who are not admitted to hospital. Regarding clinical manifestations, the main limitations were reliance on self-report measures, which risk recall biases; lack of baseline assessment, which prevents estimation of incidence; and a focus on non-specific neuropsychiatric symptoms rather than on major neurological and neuropsychiatric disorders. In particular, some of the most commonly studied symptoms (such as weakness and fatigue) have some conceptual overlap, [29] so it is possible that the prevalences found in this review may be underestimated. Terminology connoting altered mental status varied, with terms such as delirium and encephalopathy chosen in different studies, despite existing recommendation on standardisation of the nomenclature. [30] The finding that only 14.4% of the studies were of high quality limits the strength of any conclusions that can be drawn. In terms of the data synthesis, we were limited by excluding studies not published in English, which may particularly have reduced the number of important studies included from China, and the generalisability of our results may be limited by the geographical scope of the studies. Furthermore, the high heterogeneity between studies, even after subgroup analyses, suggests that variation in populations, outcomes and measurement techniques might account for much of the differences between studies. Finally, the cross-sectional nature and the focus on acute presentations of most studies reported to-date limit our ability to draw conclusions about the long-term impact of neuropsychiatric post-COVID-19 symptom burden. Future, well-designed prospective cohorts, such as the UK-based Post-hospitalisation COVID-19 study (https://www.phosp.org/), may be able to address this gap in the knowledge.

There are several implications of this review for future research. Firstly, while retrospective studies are important in identifying associations in large patient populations, they are likely to underestimate the prevalence of important symptoms. This may particularly be the case with some neuropsychiatric disorders such as depression and delirium, which are known to be generally under-recognised. [31,32] Therefore, even in the context of a pandemic, there is a need to improve the speed with which the academic community can produce prospectively designed studies, which are based on registered protocols and use validated and objective measures. Standardised case definitions and record forms for common neurological manifestations of viral infections were produced by the Brain Infections Global Network from early in the pandemic [33] and made freely available. These have been modified by other international groups, [34] and are being incorporated into the WHO case report forms. [35] More studies are required of those not admitted to hospital. In terms of the clinical manifestations measured, there appeared to be an inverse relationship between the severity of a clinical manifestation and its frequency of study, with headache, myalgia and anosmia receiving much more attention than depression, stroke and seizures. A re-balancing would be welcome with greater attention given to major neurological and neuropsychiatric disorders. Finally, the occasional waivers of ethical review and the more frequent waivers of informed consent in these studies illustrates that some aspects of study review may be overly burdensome - and therefore potentially neglected - during a pandemic. Whilst we acknowledge the need for proper ethical and institutional oversight, COVID-19 may be an opportunity for this process to be streamlined across the field, especially for non-interventional studies, where the risks to participants are minimal, so that studies during a pandemic (and beyond) can start quickly and inform urgent policy needs.

There are several clinical implications of our study. Firstly, practitioners should be aware that neurological and neuropsychiatric symptoms are very common with four (anosmia, weakness, dysgeusia and fatigue) estimated to occur in more than 30% of patients. Secondly, these non-specific neurological and neuropsychiatric symptoms appear to be the most common. Neuropsychiatric disorders such as anxiety and depression occupy an intermediate space with prevalence of between 15.9% [5.6-37.7] and 23.0% [11.8-40.2]), while major neurological disorders such as stroke and seizures are much rarer. However, because of the very high number of individuals infected with SARS-CoV-2 worldwide, even less frequent symptoms may still result in a substantial increase in the burden of disease. This means that services for those with common mental illnesses and neurological rehabilitation should be resourced and equipped for an increase in case numbers. Many of these disorders can become chronic, so the neurological and psychiatric impact of the pandemic may substantially outlast the current phase. Thirdly, given the multitude of symptoms reported, neurological and neuropsychiatric comorbidity is likely to be the norm rather than the exception in COVID-19, so there must be accessible advice and input from these specialties for patients who are acutely unwell. Finally, although there is a relative lack of data on non-hospitalised patients, the data available suggest that several symptoms, such as anosmia, dysgeusia, fatigue, headache and myalgia, are common even among those with milder illness. Although long-term evidence from this earliest literature was sparse, it gives some initial indication that the symptoms described in ‘long COVID’ may be a continuation of some of those experienced in the acute phase of the illness. [36]

In conclusion, COVID-19 is accompanied by a wide range of neurological and neuropsychiatric symptoms from the common, such as fatigue and anosmia, to the more infrequent but severe, such as stroke and seizure. There is substantial psychiatric morbidity, but a lack of control groups limits to what extent causality can be attributed.

## Data Availability

Data will be made available to interested parties following reasonable request to the first author.

## Acknowledgements

We wish to thank the many healthcare workers who have contributed inestimably to the care of these patients but are seldom recognised in the research literature.

## Supplementary material

Supplementary methods 1: Full search strategy

Supplementary methods 2: Author contributions

Supplementary table 1: PRISMA checklist

Supplementary table 2: List of extracted variables

Supplementary table 3: Full list of included studies

Supplementary table 4: comparison of proportion estimation models

Supplementary table 5: Subgroup analysis by country of origin

Supplementary figures 1-20: Forest plots for individual symptoms

## REFERENCES

1 Wu F, Zhao S, Yu B, et al. A new coronavirus associated with human respiratory disease in China. Nature 2020;579:265–9. doi:10.1038/s41586-020-2008-3

2 Rogers JP, Chesney E, Oliver D, et al. Psychiatric and neuropsychiatric presentations associated with severe coronavirus infections: a systematic review and meta-analysis with comparison to the COVID-19 pandemic. Lancet Psychiatry 2020;7:611–27. doi:10.1016/S2215-0366(20)30203-0

3 Wu Y, Xu X, Chen Z, et al. Nervous system involvement after infection with COVID-19 and other coronaviruses. Brain Behav Immun 2020;87:18–22. doi:10.1016/j.bbi.2020.03.031

4 Troyer EA, Kohn JN, Hong S. Are we facing a crashing wave of neuropsychiatric sequelae of COVID-19? Neuropsychiatric symptoms and potential immunologic mechanisms. Brain Behav Immun 2020;87:34–9. doi:10.1016/j.bbi.2020.04.027

5 Moriguchi T, Harii N, Goto J, et al. A first case of meningitis/encephalitis associated with SARS-Coronavirus-2. Int J Infect Dis IJID Off Publ Int Soc Infect Dis 2020;94:55–8. doi:10.1016/j.ijid.2020.03.062

6 Zhao H, Shen D, Zhou H, et al. Guillain-Barré syndrome associated with SARS-CoV-2 infection: causality or coincidence? Lancet Neurol 2020;19:383–4. doi:10.1016/S1474-4422(20)30109-5

7 Toscano G, Palmerini F, Ravaglia S, et al. Guillain-Barré Syndrome Associated with SARS-CoV-2. 2020. http://search.ebscohost.com/login.aspx?direct=true&db=jlh&AN=144329289&site=ehost-live

8 Helms J, Kremer S, Merdji H, et al. Neurologic Features in Severe SARS-CoV-2 Infection. N Engl J Med 2020;382:2268–70. doi:10.1056/NEJMc2008597

9 Belani P, Schefflein J, Kihira S, et al. COVID-19 Is an Independent Risk Factor for Acute Ischemic Stroke. AJNR Am J Neuroradiol Published Online First: 2020. doi:10.3174/ajnr.A6650

10 Vacchiano V, Riguzzi P, Volpi L, et al. Early neurological manifestations of hospitalized COVID-19 patients. Neurol Sci 2020;41:2029–31. doi:10.1007/s10072-020-04525-z

11 Karadaş Ö, Öztürk B, Sonkaya AR. A prospective clinical study of detailed neurological manifestations in patients with COVID-19. Neurol Sci 2020;41:1991–5. doi:10.1007/s10072-020-04547-7

12 Ellul MA, Benjamin L, Singh B, et al. Neurological associations of COVID-19. Lancet Neurol 2020;19:767–83. doi:10.1016/S1474-4422(20)30221-0

13 Lechien JR, Chiesa-Estomba CM, Place S, et al. Clinical and epidemiological characteristics of 1420 European patients with mild-to-moderate coronavirus disease 2019. J Intern Med 2020;288:335–44. doi:10.1111/joim.13089

14 Varatharaj A, Thomas N, Ellul MA, et al. Neurological and neuropsychiatric complications of COVID-19 in 153 patients: a UK-wide surveillance study. Lancet Psychiatry 2020.

15 Butler, Matt, Watson, Cameron, Rooney, Ally, et al. The Neurology and Neuropsychiatry of COVID-19. JNNP Blog. 2020.<https://blogs.bmj.com/jnnp/2020/05/01/the-neurology-and-neuropsychiatry-of-covid-19/> (accessed 23 Jan 2021).

16 Favas TT, Dev P, Chaurasia RN, et al. Neurological manifestations of COVID-19: a systematic review and meta-analysis of proportions. Neurol Sci Off J Ital Neurol Soc Ital Soc Clin Neurophysiol 2020;41:3437–70. doi:10.1007/s10072-020-04801-y

17 Islam MA, Alam SS, Kundu S, et al. Prevalence of Headache in Patients With Coronavirus Disease 2019 (COVID-19): A Systematic Review and Meta-Analysis of 14,275 Patients. Front Neurol 2020;11:562634. doi:10.3389/fneur.2020.562634

18 Pinzon RT, Wijaya VO, Buana RB, et al. Neurologic Characteristics in Coronavirus Disease 2019 (COVID-19): A Systematic Review and Meta-Analysis. Front Neurol 2020;11:565. doi:10.3389/fneur.2020.00565

19 Wang L, Shen Y, Li M, et al. Clinical manifestations and evidence of neurological involvement in 2019 novel coronavirus SARS-CoV-2: a systematic review and meta-analysis. J Neurol 2020.

20 Moher D, Shamseer L, Clarke M, et al. Preferred reporting items for systematic review and meta-analysis protocols (PRISMA-P) 2015 statement. Syst Rev 2015;4:1. doi:10.1186/2046-4053-4-1

21 OCEBM Levels of Evidence Working Group. The Oxford Levels of Evidence 2. OCEBM Levels Evid. https://www.cebm.ox.ac.uk/resources/levels-of-evidence/ocebm-levels-of-evidence (accessed 23 Jan 2021).

22 Ottawa Hospital Research Institute. The Newcastle-Ottawa Scale (NOS) for assessing the quality of nonrandomised studies in meta-analyses. http://www.ohri.ca/programs/clinical_epidemiology/oxford.asp (accessed 20 Jan 2021).

23 Modesti PA, Reboldi G, Cappuccio FP, et al. Panethnic Differences in Blood Pressure in Europe: A Systematic Review and Meta-Analysis. PLOS ONE 2016;11:e0147601. doi:10.1371/journal.pone.0147601

24 Schwarzer G, Chemaitelly H, Abu-Raddad LJ, et al. Seriously misleading results using inverse of Freeman-Tukey double arcsine transformation in meta-analysis of single proportions. Res Synth Methods 2019;10:476–83. doi:10.1002/jrsm.1348

25 Hamza TH, van Houwelingen HC, Stijnen T. The binomial distribution of meta-analysis was preferred to model within-study variability. J Clin Epidemiol 2008;61:41–51. doi:10.1016/j.jclinepi.2007.03.016

26 Stijnen T, Hamza TH, Özdemir P. Random effects meta-analysis of event outcome in the framework of the generalized linear mixed model with applications in sparse data. Stat Med 2010;29:3046–67. doi:https://doi.org/10.1002/sim.4040

27 R Core Team. R: A Language and Environment for Statistical Computing. 2017.https://www.R-project.org/

28 Viechtbauer W. Conducting Meta-Analyses in R with the metafor Package. J Stat Softw2010;36:1–48. doi:10.18637/jss.v036.i03

29 Saguil A. Evaluation of the Patient with Muscle Weakness. Am Fam Physician 2005;71:1327–36.

30 Slooter AJC, Otte WM, Devlin JW, et al. Updated nomenclature of delirium and acute encephalopathy: statement of ten Societies. Intensive Care Med 2020;46:1020–2. doi:10.1007/s00134-019-05907-4

31 Chin YC, Koh GCH, Tay YK, et al. Underdiagnosis of delirium on admission and prediction of patients who will develop delirium during their inpatient stay: a pilot study. Singapore Med J 2016;57:18–21. doi:10.11622/smedj.2016007

32 Balestrieri M, Bisoffi G, Tansella M, et al. Identification of depression by medical and surgical general hospital physicians. Gen Hosp Psychiatry 2002;24:4–11. doi:10.1016/S0163-8343(01)00176-1

33 COVID-Neuro Network • Brain Infections Global. <https://braininfectionsglobal.tghn.org/covid-neuro-network/>(accessed 9 Feb 2021).

34 Winkler A.S., Knauss S., Schmutzhard E., et al. A call for a global COVID-19 Neuro Research Coalition. Lancet Neurol 2020;19:482–4. doi:10.1016/S1474-4422%2820%2930150-2

35 Global COVID-19 Clinical Platform: Rapid core case report form (CRF). https://www.who.int/publications-detail-redirect/WHO-2019-nCoV-Clinical_CRF-2020.4 (accessed 9 Feb 2021).

36 CarfÍ A, Bernabei R, Landi F. Persistent Symptoms in Patients After Acute COVID-19. JAMA J Am Med Assoc 2020;324:603–5. doi:10.1001/jama.2020.12603

