## supplementary methods 1 for "The neurology and neuropsychiatry of COVID-19: a systematic review and meta-analysis of the early literature reveals frequent CNS manifestations and key emerging narratives"

**Supplementary methods 1: Full search strategy**

Per phase one of our strategy and for the current manuscript we populated the database with a search extending from 1^st^ Jan 2020 to 18^th^ July 2020. Future phases will update the database on a progressively shortening basis. To enable this we will continuously run the processes described below. Successive iterations of the database will be version and date-controlled.

**Information sources**

We searched Ovid MEDLINE(R) and Epub Ahead of Print, In-Process & Other Non-Indexed Citations and Daily, EMBASE (via Ovid), APA PsycInfo (via OVID) and CINAHL (via EBSCO) from 1st Jan 2020 to 18th July 2020. Individualised search strategies for each database were deposited with CRD. Reference lists of systematic review articles were examined and cross-checked against our database and eligibility criteria. We noted foreign language papers but did not translate these.

**Search Strategy**

**MEDLINE**

(Coronavirus or corona virus or coronavirinae or coronaviridae or betacoronavirus or Covid19 or Covid 19 or Covid-19 or nCoV or CoV 2 or CoV2 or CoV-2 or Sarscov2 or SARS-CoV-2 or 2019nCoV or novel CoV or Wuhan virus or ((Wuhan or Hubei or Huanan) and (respiratory or pneumonia or virus))).mp. or exp Coronavirus/ or exp Coronavirus Infections/ or Coronaviridae.mp.

AND

(neurol* or nervous or brain or CNS or encephal* or mening* or Cranial* or myeli* or demyeli* or ADEM or ataxi* or dysphasi* or aphasi* or stroke or guillain-barre or Miller-Fisher or paresis or palsy or cerebr* or crani* or epilep* or seizure or headache* or migraine* or dysgeusia or anosmia or taste or smell or psych* or neuropsych* or mania or manic or psycho* or delusion* or hallucin* or functional or catatoni* or cognit* or dement* or delir* or depress* or anxi* or obsess* or post-traum* or posttraum* or PTSD or behaviour or behavior or fatigue* or MRI or CT or neuroimag* or scan* or neurotrop* or neuroinvas* or neuropath* or cerebrospinal* or cerebro-spinal* or confus* or conscious* or letharg* or psychomotor* or psycho-motor* or agitat*).mp. or exp Neurology/ or exp Nervous System/ or exp Nervous System Diseases/ or exp Neurologic Manifestations/ or exp Psychiatry/ or exp Mental Disorders/ or exp Mental Processes/ or exp Behavioral Symptoms/ or exp Psychological Phenomena/

**EMBASE**

(Coronavirus or corona virus or coronavirinae or coronaviridae or betacoronavirus or Covid19 or Covid 19 or Covid-19 or nCoV or CoV 2 or CoV2 or CoV-2 or Sarscov2 or SARS-CoV-2 or 2019nCoV or novel CoV or Wuhan virus or ((Wuhan or Hubei or Huanan) and (respiratory or pneumonia or virus))).mp. or exp Coronavirinae/ or exp Coronavirus Infection/ or exp Coronaviridae/ or exp Coronaviridae infection/ or exp SARS Coronavirus/

AND

(neurol* or nervous or brain or CNS or encephal* or mening* or Cranial* or myeli* or demyeli* or ADEM or ataxi* or dysphasi* or aphasi* or stroke or guillain-barre or Miller-Fisher or paresis or palsy or cerebr* or crani* or epilep* or seizure or headache* or migraine* or dysgeusia or anosmia or taste or smell or psych* or neuropsych* or mania or manic or psycho* or delusion* or hallucin* or functional or catatoni* or cognit* or dement* or delir* or depress* or anxi* or obsess* or post-traum* or posttraum* or PTSD or behaviour or behavior or fatigue* or MRI or CT or neuroimag* or scan* or neurotrop* or neuroinvas* or neuropath* or cerebrospinal* or cerebro-spinal* or confus* or conscious* or letharg* or psychomotor* or psycho-motor* or agitat*).mp. or exp Neuroscience/ or exp Nervous System/ or exp Neurologic Disease/ or exp Psychiatry/ or exp Mental Disease/ or exp Behavior/ or exp Mental Function/ or exp Confusion/ or exp Psychophysiology/

**PsycINFO**

(Coronavirus or corona virus or coronavirinae or coronaviridae or betacoronavirus or Covid19 or Covid 19 or Covid-19 or nCoV or CoV 2 or CoV2 or CoV-2 or Sarscov2 or SARS-CoV-2 or 2019nCoV or novel CoV or Wuhan virus or (Wuhan or Hubei or Huanan)).mp.

AND

(neurol* or nervous or brain or CNS or encephal* or mening* or Cranial* or myeli* or demyeli* or ADEM or ataxi* or dysphasi* or aphasi* or stroke or guillain-barre or Miller-Fisher or paresis or palsy or cerebr* or crani* or epilep* or seizure or headache* or migraine* or dysgeusia or anosmia or taste or smell or psych* or neuropsych* or mania or manic or psycho* or delusion* or hallucin* or functional or catatoni* or cognit* or dement* or delir* or depress* or anxi* or obsess* or post-traum* or posttraum* or PTSD or behaviour or behavior or fatigue* or MRI or CT or neuroimag* or scan* or neurotrop* or neuroinvas* or neuropath* or cerebrospinal* or cerebro-spinal* or confus* or conscious* or letharg* or psychomotor* or psycho-motor* or agitat*).mp OR exp Psychiatry/ OR exp Mental Disorders/ OR exp Sensory System Disorders/ OR exp Sense Organ Disorders/ OR exp Nervous System Disorders/ OR exp Neurosciences/ OR exp Neurocognitive Disorders or exp Emotional States

**CINAHL**

(Coronavirus or corona virus or coronavirinae or coronaviridae or betacoronavirus or Covid19 or Covid 19 or Covid-19 or nCoV or CoV 2 or CoV2 or CoV-2 or Sarscov2 or SARS-CoV-2 or 2019nCoV or novel CoV or Wuhan virus or ((Wuhan or Hubei or Huanan) and (respiratory or pneumonia or virus))) OR (MH "Coronavirus+") OR (MH "Coronavirus Infections+") OR (MH "Coronaviridae+")

AND

( (neurol* or nervous or brain or CNS or encephal* or mening* or Cranial* or myeli* or demyeli* or ADEM or ataxi* or dysphasi* or aphasi* or stroke or guillain-barre or Miller-Fisher or paresis or palsy or cerebr* or crani* or epilep* or seizure or headache* or migraine* or dysgeusia or anosmia or taste or smell or psych* or neuropsych* or mania or manic or psycho* or delusion* or hallucin* or functional or catatoni* or cognit* or dement* or delir* or depress* or anxi* or obsess* or post-traum* or posttraum* or PTSD or behaviour or behavior or fatigue* or MRI or neuroimag* or scan* or neurotrop* or neuroinvas* or neuropath* or cerebrospinal* or cerebro-spinal* or confus* or conscious* or letharg* or psychomotor* or psycho-motor* or agitat*) OR ) OR (MH "Neurology") OR (MH "Nervous System+") OR (MH "Nervous System Diseases+") OR (MH "Neurologic Manifestations+") OR (MH "Psychiatry+") OR (MH "Mental Processes+") OR (MH "Diagnosis, Neurologic+") OR ( (MH "Behavioral and Mental Disorders+") )
