## supplementary methods 2 for "The neurology and neuropsychiatry of COVID-19: a systematic review and meta-analysis of the early literature reveals frequent CNS manifestations and key emerging narratives"

**Supplementary Methods 2: Author contributions**

| **Author** | **Contribution(s)** |
| --- | --- |
| *All authors* | - Made a substantial intellectual contribution to the study - Approved the final manuscript |
| Dr Jonathan P Rogers | - Led and coordinated final phase of study - Wrote manuscript - Conducted quality assessment - Sorted references - Calculated descriptive statistics - Consulted on study inclusion - Arbitrated with quality assessment - Checked completed manuscript |
| Dr Cameron Watson | - Conducted meta-analysis - Screened studies for eligibility - Extracted data |
| Mr James Badenoch | - Screened studies for eligibility - Extracted data - Checked data extraction - Conducted quality assessment - Checked adherence to PRISMA guidelines - Made PRISMA flowchart - Sorted references - Drafted limitations of study - Checked completed manuscript |
| Dr Benjamin Cross | - Screened studies for eligibility - Extracted data - Checked data extraction - Conducted quality assessment |
| Dr Matthew Butler | - Screened studies for eligibility - Consulted on screening studies - Checked data extraction - Conducted quality assessment - Compared to other systematic reviews - Drafted limitations of study - Checked citations - Checked completed manuscript |
| Dr Jia Song | - Screened studies for eligibility - Consulted on screening studies - Checked data extraction - Conducted quality assessment - Checked completed manuscript |
| Mr Danish Hafeez | - Screened studies for eligibility - Extracted data - Checked data extraction - Conducted quality assessment - Assisted in creating figure illustrating change in study design |
| Dr Hamilton Morrin | - Extracted data - Checked data extraction - Conducted quality assessment |
| Dr Emma Rachel Rengasamy | - Screened studies for eligibility - Extracted data - Checked data extraction - Assisted in writing results - Made supplementary table with complete list of studies |
| Miss Lucretia Thomas | - Screened studies for eligibility - Extracted data - Checked data extraction - Conducted quality assessment |
| Dr Silviya Ralovska | - Extracted Data - Checked data extraction - Conducted quality assessment |
| Ms Abigail Smakowski | - Checked data extraction - Conducted quality assessment - Supervised quality assessment - Arbitrated quality assessment |
| Miss Ritika Dilip Sundaram | - Extracted data - Checked data extraction - Conducted quality assessment |
| Ms Camille Kaitlyn Hunt | - Checked data extraction - Conducted quality assessment |
| Dr Mao Fong Lim | - Checked data extraction - Conducted quality assessment - Arranged funding statements |
| Dr Daruj Aniwattanapong | - Checked data extraction |
| Ms Vanshika Singh | - Conducted quality assessment - Drafted implications for clinicians |
| Dr Zain Hussain | - Conducted quality assessment - Assisted with creating tables of results |
| Miss Stuti Chakraborty | - Conducted quality assessment |
| Miss Ella Burchill | - Conducted quality assessment - Adapted to house style |
| Katrin Jansen | - Supported with meta-analysis methods - Conducted some of the meta-analyses |
| Prof Dr Heinz Holling | - Supported with meta-analysis methods - Advised on meta-analysis |
| Dr Dean Walton | - Conducted quality assessment - Provided senior review of manuscript |
| Dr Thomas A Pollak | - Conducted quality assessment - Provided senior review of manuscript |
| Dr Mark Ellul | - Conducted quality assessment - Provided senior review of manuscript |
| Dr Ivan Koychev | - Conducted quality assessment - Provided senior review of manuscript |
| Professor Tom Solomon | - Provided senior review of manuscript |
| Dr Benedict Daniel Michael | - Provided senior review of manuscript |
| Dr Timothy R Nicholson | - Conceived the study - Provided senior leadership and advice throughout - Conducted quality assessment - Provided senior review of manuscript |
| Dr Alasdair G Rooney | - Conceived the study - Led and coordinated early phases of study - Wrote manuscript - Screened studies - Consulted on screening studies - Checked data extraction - Conducted OCEBM ratings - Conducted quality assessment - Provided senior review of manuscript |
