## supplementary table 2 for "The neurology and neuropsychiatry of COVID-19: a systematic review and meta-analysis of the early literature reveals frequent CNS manifestations and key emerging narratives"

**Supplementary table 2: Comprehensive list of fields which were extracted from eligible studies**

(*asterisk denotes variables for which outcome measures were collected*)

| Data items | Prevalences | Mechanistic data | Analytical statistics |
| --- | --- | --- | --- |
| Title of study | Headache | C-reactive protein | Results from statistical tests reported by a study |
| CEBM Levels of Evidence | Myalgia | D-dimer |  |
| Date of date collection | Asthenia | Antibodies |  |
| Funding source | Dizziness/ vertigo | Cytokines |  |
| Country of origin | Tinnitus | Creatine kinase |  |
| Ethics statement | Hearing impairment | Computed tomography |  |
| Number of recruitment settings | Ataxia | Magnetic resonance imaging |  |
| Study design (including retrospective/ prospective) | Paresis | Electroencephalography |  |
| Number of infected participants | Obsessions | Cerebrospinal fluid analysis |  |
| Number of control participants | Pasly | Post-mortem findings |  |
| Description of control group | Paraesthesia/numbness |  |  |
| Population age (mean/median, sd/range) | Speech/aphasia |  |  |
| Population sex (number male) | Focal neurological deficit |  |  |
| Method of COVID-19 diagnosis | Seizures |  |  |
| Stage of COVID-19 pathway | Neuralgia |  |  |
| Severity of COVID-19 | Ophthalmoplegia |  |  |
| COVID-19 outcome | Visual defect |  |  |
| Follow-up period | Fatigue* |  |  |
| Time-point in relation to infection | Impaired smell* |  |  |
| Treatments for neuropsych manifestations | Impaired taste* |  |  |
| Neuropsychiatric and other relevant comorbidities | Cognitive impairment* |  |  |
| Methodological limitations as noted by the authors | Depression* |  |  |
| temporal onset of neuropsych complication (in relation to COVID-19 disease) | Anxiety* |  |  |
| Method of ascertaining prevalence | Post-traumatic stress disorder* |  |  |
| Nature of sample | Impaired consciousness/delirium/encephalopathy* |  |  |
|  | Stroke |  |  |
|  | Meningoencephalitis |  |  |
|  | Guillain-Barre syndrome |  |  |
|  | Central demyelinating disorders |  |  |
|  | Motor neuropathy |  |  |
|  | Migraine |  |  |
|  | Sleep disorder |  |  |
|  | Catatonia |  |  |
|  | Dementia |  |  |
|  | Mania |  |  |
|  | Functional neurological disorder |  |  |
|  | Psychosis |  |  |
|  | Post-intensive care unit neuropsychiatric abnormality |  |  |
|  | Myopathy |  |  |
|  | Movement disorder |  |  |
