## supplementary table 3 for "The neurology and neuropsychiatry of COVID-19: a systematic review and meta-analysis of the early literature reveals frequent CNS manifestations and key emerging narratives"

**Supplementary Table 1: Summary table of studies reporting neuropsychiatric outcomes of COVID-19 infected patients.**

|  | **Study** | **Setting** | **Population** | **Design** | **Infected sample (cases)** | **Mean Age (S.D.)** | **Male (%)** | **Outcomes** |
| --- | --- | --- | --- | --- | --- | --- | --- | --- |
| (1) | Nie et al. (2020) | China | Inpatient | Cross-sectional | 78 | 58.4 (13) | 33 (42.3%) | Depression 28 (35.9%), Anxiety 30 (38.5%), Depression and Anxiety 19 (24.3%) |
| (2) | Radmanesh et al. (2020) | USA | Mixed | Case series | 242 | 68.7 (16.5) | 150 | Headache 5 (2%), Weakness 2 (0.8%), Dizziness/Vertigo 79 (32.6%), Focal neurological deficit 30 (12.4%), Altered mental status 102 (42.1%), Acute haemorrhagic stroke 11 (4.5%), Nonspecific white matter microangiopathy 134 (55.4%), Chronic infarct 47 (19.4%), Acute or subacute ischemic infarct 13 (5.4%) |
| (3) | Wei Y et al. (2020) | China | Outpatient | Cross-sectional | 628 | 53 (14.8) | 296 | Myalgia 8 (1.3%), Dizziness/Vertigo 1 (0.2%), Fatigue 42 (6.7%) |
| (4) | Haehner et al. (2020) | Germany | Outpatient | Cross-sectional | 34 | - | 227 | Impaired smell onset 22 (64.7%), Impaired taste onset 22 (2.94%) |
| (5) | Beltran-Corbellini et al. (2020) | Spain | Inpatient | Cross-sectional | 79 | 61.6 (17.4) | 48 | Impaired smell and taste 31 (39.2%), Impaired smell onset 25 (31.7%), Impaired smell at follow-up 13 (16.5%), Impaired taste at onset 28 (35.4%) |
| (6) | Renaud et al. (2020) | France | Outpatient | Case series | 347 | 37* (-) | 113 | Impaired smell onset 345 (99.4%), Impaired taste at onset 123 (35.4%) |
| (7) | Gao et al. (2020) | China | Inpatient | Case series | 210 | 71* (-) | 101 | Headache 13 (6%), Myalgia 12 (6%), Fatigue 73 (5%), Ischaemic stroke 3 (1%) |
| (8) | Abalo-Lojo et al. (2020) | Spain | - | Case series | 131 | 50.4 (-) | 56 | Headache 51 (38.9%), Myalgia 61 (46.6%), Asthenia 76 (58%), Impaired smell onset 77 (58.8%), Impaired taste at onset 74 (56.5%) |
| (9) | Adorni et al. (2020) | Italy | Volunteers | Cross-sectional | 170175  856 | 47.4 (14.5) | 68767 | Headache 485 (56.7%), Myalgia 527 (61.6%), Smell and/or taste 507 (59.2%) |
| (10) | Roland et al. (2020) | USA | Volunteers (online) | Cross-sectional | 157 | 40 (13) | 51 | Headache 93 (64%), Myalgia 25 (77%), Smell or taste 95 (66%) |
| (11) | Aggarwal et al. (2020) | USA | Inpatient | Case series | 16 | 65.5 (Range 38-95) | 12 (75%) | Headache 4 (25%), Myalgia 4 (25%), Dizziness/Vertigo 3 (19%), Fatigue 8 (50%), Impaired smell 3 (19%), |
| (12) | Alsofayan Y.M. et al. (2020) | Saudi Arabia | Mixed | Cross-sectional | 1519 | 36 * (-) | 825 | Headache 193 (27.3%), Myalgia 202 (28.6%) |
| (13) | Romero-Sanchez et al. (2020) | Spain | Inpatient | Case series | 841 | 66.4 (15.0) | 473 | Headache 119 (14.1%), Myalgia 145 (17.2%), Asthenia 425 (51%), Dizziness/Vertigo 51 (6.1%), Seizures 6 (0.7%), Ophthalmoplegia 1 (0.1%), Impaired smell onset 41 (4.9%), Impaired taste at onset 52 (6.2%), Depression 44 (5.2%), Anxiety 68 (8.1%), Impaired consciousness 165 (19.6%), Ischaemic stroke 11 (1.3%), Haemorrhagic stroke 3 (0.4%), Meningoencephalitis 1 (0.1%), Guillain-Barre syndrome and variants 1 (0.1%), Sleep disorders 109 (13%), Psychosis 11 (1.3%), Myopathy 26 (3.1%), Movement disorder 6 (0.7%) |
| (14) | Sakalli et al. (2020) | Turkey | Mixed | Cohort | 172 | 37.8 (12.5) | 84 | Headache 97 (56.4%), Myalgia 125 (72.7%), Anosmia and Dysgeusia 72 (41.9%), Impaired smell onset 88 (51.2%), Impaired smell follow-up 68 (39.5%), Impaired taste onset 81 (47.1%), Impaired taste follow-up 62 (36%) |
| (15) | Salmon Ceron et al. (2020) | France | Outpatient | Cohort | 51 | 34* (Range 22-61) | 24 | Headache 38 (69.1%), Myalgia 17 (30.9%), Asthenia 28 (56%), Impaired smell onset 55 (100%), Impaired smell follow-up 35 (63.6%), Impaired taste onset 46 (83.6%) |
| (16) | Chen et al. (2020) | China | Inpatient | Case series | 145 | 47.5 (14.6) | 54.50% | Headache 24 (16.6%), Myalgia 20 (13.8%), Dizziness/Vertigo 29 (20%), Hearing impairment 2 (1.4%). Fatigue 59 (40.7%), Anorexia 62 (42.8%), Hypoacusis 2 (1.4%) |
| (17) | Chen et al. (2020) | China | Inpatient | Cohort | 203 | 54 * (IQR 41-68) | 108 (53.2%) | Headache 10 (4.9%), Dizziness/Vertigo 4 (2%), Myalgia or Arthralgia 54 (26.6%), Fatigue 16 (7.9%), Anorexia 6 (3%) |
| (18) | Liu et al. (2020) | China | Inpatient | Case-control | 21 | 43.1 (2.6) | 9 | Suspected Depression 21 (100%), Suspected Anxiety 21 (100%) |
| (19) | Chiesa-Estomba et al. (2020) | South America | Online/outpatient | Cross-sectional | 542 | 34 (11) (Range 18-88) | 218 (40.2) | Headache 393 (72.5%), Myalgia 340 (62.7%), Total anosmia 78 (14.4%) Impaired smell onset 444 (81.9%), Impaired smell follow-up 183 (33.8%), Impaired taste onset 333 (61.4%) |
| (20) | Lombardi et al. (2020) | Italy | Healthcare workers | Case-control | 139 | 44.5 (-) | 57 | Headache 24 (17.3%), Myalgia 20 (14.4%), Asthenia 22 (15.8%), Ophthalmoplegia 13 (9.3%), Anosmia and Dysgeusia 20 (14.3%) |
| (21) | Oualha et al. (2020) | France | Inpatient | Case series | 27 | 6* (Range 0.2-17.8) | 10 | Paresis 1 (4%), Focal neurological deficit 1 (4%), Stupor and Reduced GCS 1 (4%), Haemorrhagic stroke 2 (7.4%), Meningoencephalitis 1 (4%) |
| (22) | Paderno et al. (2020) | USA | Mixed | Cross-sectional | 508 | *Group A* (hospital) = 61.9 (12.8), *Group B* (home) = 44.7 (12.1) | Group A (hospital) = 204 Group B (home) = 81 | Headache 198 (38.9%), Myalgia 256 (50.4%), Asthenia 352 (69.3%), Impaired smell onset 283 (71.8%), Impaired smell follow-up 29 (19.7%), Impaired taste onset 321 (63.1%), Impaired taste follow-up 35 (19.7%) |
| (23) | Kim et al. (2020) | South Korea | Inpatient | Cohort | 2491 | 62* (25) | 1326 | Myalgia 722 (29.1%), Fatigue 722 (29.1%), Encephalitis 151 (6.1%), Central demyelinating disorders 151 (6.1%) |
| (24) | Kim et al. (2020) | South Korea | Community | Cross-sectional | 213 | 26 * (-) | 66 | Headache 54 (25.3%), Myalgia 54 (25.3%), Dizziness/Vertigo 32 (15%), Fatigue 46 (21.6%), Impaired smell onset 68 (31.9%), Impaired taste onset 58 (27.2%) |
| (25) | Kimball et al. (2020) | USA | Inpatient | Cohort | 23 | 80.7 (8.4) | 7 | Dizziness/Vertigo 1 (4.4%), Fatigue 4 (17.4%) |
| (26) | Klopfenstein et al. (2020) | France | Mixed | Case series | 114 | - | - | Anosmia and Dysgeusia 46 (38%). Impaired smell onset 54 (47.4%) |
| (27) | Lovato et al. (2020) | Italy | Telephone survey- Mild/mixed | Cross-sectional | 121 | 46.7 (-) | 49 | Anosmia and Dysgeusia 121 (83%) |
| (28) | Chiesa-Estomba et al. (28) | South America | Mixed | Cohort | 1221 | 41 (13) | - | Impaired smell onset 970 (79.4%), Impaired smell follow-up 381 (50.7%) |
| (29) | Chougar et al. (29) | France | Inpatient | Case series | 308 | 61.8 (14.9) | 64 | Fatigue 33 (10.7%), Focal neurological deficit 45 (14.6%), Seizures 16 (5.2%) , Visual defect 6 (19.5%), Impaired smell onset 28 (%), Impaired consciousness 89 (28.9%), Guillain-Barre syndrome and variants 3 (1%) |
| (30) | Chung et al. (2020) | Hong Kong | Inpatient (mild) | Cross-sectional | 18 | 28 (19) | 41 | Anosmia and Dysgeusia 9 (52.9%), Impaired smell onset 12 (67%) |
| (31) | Gaur et al. (2020) | India | Inpatient | Cohort | 26 | 37.6 (-) | 16 (61.5%) | Headache 8 (30.8%), Myalgia 10 (38.5%), |
| (32) | Altin et al. (2020) | Turkey | Inpatient | Case-control | 81 | 54.2 (17.0) | 41 (50.6%) | Impaired smell onset 50 (61.7%), Impaired taste onset 22 (27.2%) |
| (33) | Annweiler et al. (2020) | France | Mixed | Cross-sectional | 353 | 84.7 (7) | 160 (45.3%) | Impaired smell onset 7 (2%), Impaired taste onset 25 (7.1%), Altered consciousness 37 (10.5%) |
| (34) | Gómez-Iglesias et al. (2020) | Spain | Population | Cross-sectional | 909 | 34.7 (-) | 283 (31.1%) | Myalgia 296 (32.5%), Smell and Taste 824 (90.7%), Impaired smell onset 888 (97.7%), Impaired taste onset 845 (93%) |
| (35) | Clemency et al. (2020) | USA | Outpatients – HCWs | Cross-sectional | 225 | - | - | Myalgia 128 (56.8%), Smell and Taste 110 (48.9%), Fatigue 150 (66.7%) |
| (36) | Coelho et al. (2020) | USA | General population | Cohort | 260 | - | - | Headache 76 (81.7%), Myalgia 75 (80.6%), Fatigue 81 (87.1%), Smell or Taste 3 (3.2%) |
| (37) | Gorzkowski et al. (2020) | France | Mixed | Cohort | 229 | 39.7 (13.7) | 82 (35.8%) | Smell or Taste 140 (61.1%), Impaired smell at onset 124 (54.2%), Impaired smell at follow-up 6 (2.6%), Impaired taste at onset 140 (61.1%) |
| (38) | Han et al. (2020) | China | Inpatient | Cohort | 32 | 44 (-) | 12 (37.5%) | Myalgia or Fatigue 13 (52%) |
| (39) | Lovell et al. (2020) | UK | Inpatients (referred to palliative care) | Case series | 101 | 82 (IQR 72-89) | 64 (63.3%) | Seizures 1 (1%), Fatigue 9 (8.9%), Delirium 24 (23.8%) |
| (40) | Lu et al. (2020) | China | Discharged or deceased | Cohort | 304 | 44* (IQR 33-59) | 182 (59.9%) | Seizures 2 (0.7%), Anxiety 1 (0.3%), Shock 8 (2.60%), Other cerebrovascular accident 4 (1.3%) |
| (41) | Sayin et al. (2020) | Turkey | Mixed | Case-control | 64 | 38.6 (10.1) | 48 (75%) | Headache 18 (39.1%), Smell or Taste 46 (71.9%), Impaired smell onset 43 (67.2%), Impaired taste onset 46 (71.9%) |
| (42) | Schmithausen et al. (2020) | Germany | Mild cases – home quarantined | Cross-sectional | 41 | 40* (Range 18-92) | 20 (49%) | Smell and/or taste 28 (68%) |
| (43) | Scullen et al. (2020) | USA | Inpatient | Case series | 27 | 59.8 (Range 35-91) | 14 (52%) | Headache 2 (2.63%), Weakness 1 (3.,7%), Focal neurological deficit 10 (13.2%), Impaired taste at onset 1 (1.32%), Altered mental status 26 (34,2%), Other cerebrovascular accident 4 (14.8%) |
| (44) | Belani et al. (2020) | USA | Inpatients undergoing neuroimaging for suspected stroke | Case-control | 34 | - | - | Stroke 19 (56%) |
| (45) | Bhandari et al. (2020) | India | Inpatient | Cohort | 522 | 35.4 (-) | 318 (60.91%) | Headache 139 (26.7%) |
| (46) | Hernández-Fernández et al. (2020) | Spain | Inpatient | Case series | 1683 | 66.8 (-) | 18 (1.1%) | Ischaemic stroke 17 (1.0%), Haemorrhagic stroke 5 (0.3%), Other cerebrovascular accident 1 (0.1%) |
| (47) | Covino et al. (2020) | Italy | Inpatient | Case series | 69 | 84* (IQR 82-89) | 37 (53.6%) | Fatigue 8 (11.6%) |
| (48) | Biadsee et al. (2020) | Israel | Self-isolating/non-hospitalised | Cross-sectional | 128 | 36.3 (Range 18-73) | 58 (45.3%) | Headache 52 (40%), Myalgia 60 (47%), Weakness 61 (48%), Impaired smell onset 49 (38%), Impaired taste onset 42 (32%) |
| (49) | Hintschich et al. (2020) | Germany | Home quarantine | Case-control | 41 | 37 (-) | 12 (29.3%) | Smell and Taste 18 (44%), Impaired smell onset 22 (54%), Impaired taste onset 8 (20%) |
| (50) | Sierpinski et al. (2020) | Poland | Self-isolating | Cross-sectional | 1942 | 50* (-) | 773 (39.8%) | Smell and Taste 825 (42.5%), Impaired smell onset 956 (49.2%), Impaired taste onset 923 (47.5%) |
| (51) | Paderno et al. (2020) | Italy | Mixed | Cohort | 151 | 45 (Range 18-70) | 56 (37%) | Impaired smell onset (complete anosmia) 93 (74%), Impaired taste (complete dysgeusia) 95 (70%) |
| (52) | Hornuss et al. (2020) | Germany | Inpatient | Case-control | 45 | 56 (16.9) | 25 (55.6%) | Headache 10 (22%), Impaired smell onset 38 (84.4%) |
| (53) | Boscolo-Rizzo et al. (2020) | Italy | Outpatient - mild | Cross-sectional | 202 | 56*(-) | 99 (49%) | Headache 80 (39.6%), Myalgia 85 (42%), Dizziness/vertigo 25 (12.3%), Smell or Taste 113 (55.9%), Fatigue. 130 (64.3%) |
| (54) | Huang et al. (2020) | China | Inpatient | Cohort | 41 | 49 (-) | 30 (73.2%) | Headache 3 (8%), Myalgia or Fatigue 18 (44%) |
| (55) | Huang et al. (2020) | China | Inpatient | Case series | 336 | 43 (IQR 30-54) | 182 (54.1%) | Headache 39 (11.6%), Myalgia 39 (11.6%), Weakness OR Fatigue 83 (24.7%), Impaired consciousness/delirium/encephalopathy 1 (0.3%) |
| (56) | Parra et al. (2020) | Spain | Inpatient | Case series | 10 | 54.1(10.7) | 6 (60%) | Psychosis 10 (100%) |
| (57) | Luers et al. (2020) | Germany | Outpatient | Cross-sectional | 72 | 38 (-) | 41(56.9%) | Headache 56 (77.8%), Myalgia 51 (70.8%), Impaired smell onset 53 (73.6%), Impaired taste onset 50 (69.4%) |
| (58) | Somekh et al. (2020) | Israel | Outpatient | Cross-sectional | 73 | - | - | Smell or Taste 37 (51%) |
| (59) | Song et al. (2020) | China | Inpatient | Case series | 111 | 55*( /IQR 44-67) | 62(55.9%) | Headache 11 (9.9%), Myalgia 21 (18.9%), Fatigue 37 (33.3%) |
| (60) | Immovilli et al. (2020) | Italy | Inpatient | Case series | 19 | 76.1 (8.8) | 10 (52.6%) | Ischaemic stroke 17 (89.5%), Haemorrhagic stroke 2 (10.5%) |
| (61) | Pastor et al. (2020) | Spain | Inpatient | Case-control | 20 | 63.9 (-) | 17 (85%) | Impaired consciousness/delirium/encephalopathy 18 (90%) |
| (62) | Izquierdo-Domínguez et al. (2020) | Spain | Mixed | Cross-sectional | 846 | 56.8 (15.7) | 446 (52.1%) | Smell and Taste 399 (47.2%), Impaired smell onset 454 (53.7%), Impaired taste onset 442 (52.2%) |
| (63) | Ma et al. (1) (2020) | China | Inpatients | Case series | 93 | 67* (/IQR 54-72) | 51 (54.8%) | Headache 14 (15.1%), Myalgia 20 (21.5%) |
| (64) | Ma et al. (2) (2020) | China | Inpatient | Cross-sectional | 770 | 50.4 (13.1) | 370 (48.0%) | Depression 332 (43.1%) |
| (65) | Mao et al. (2020) | China | Outpatient | Case series | 188 | 46* (-) | 94 (50%) | Headache 23 (12%), Myalgia 36 (19%), Fatigue 63 (34%) |
| (66) | Marinho et al. (2020) | Brazil | Outpatients (majority physicians) | Case series | 12 | - (Range 25-69) | 6 (50%) | Asthenia 12 (100%), Impaired smell onset 11 (91.7%) |
| (67) | Mariotto et al. (2020) | Italy | Inpatient | Cross-sectional | 107 | 65.8 (11.9) | 82 (76.6%) | Headache 12 (17.4%), Myalgia 16 (23.2%), Dizziness/vertigo 6 (8.7%), Fatigue 47 (68.1%), Impaired smell onset 15 (21.7%), Impaired taste onset 22 (31.9%), Impaired consciousness/delirium/encephalopathy 13 (18.8%) |
| (68) | Mathian et al. (2020) | France |  | Case series | 17 | 53.5* (Range 27-69) | 4 (23.5%) | Headache 10 (59%), Myalgia 8 (47%), Impaired smell onset 5 (29%), Impaired taste onset 5 (29%), Impaired consciousness/delirium/encephalopathy 1 (6%) |
| (69) | McLoughlin et al. (2020) | UK | Inpatient | Cross-sectional | 71 | 61 (Range 24-91) | 51(71.8%) | Impaired consciousness/delirium/encephalopathy 31 (43.7%), Sleep disorders 16 (22.5%), Psychosis 5 (7%) |
| (70) | Patel et al. (2020) | UK | Mixed | Cross-sectional | 141 | 45.6 (-) | 83 | Myalgia 93 (66%), Impaired smell onset 80 (56.7%), Impaired taste onset 89 (63.1%) |
| (71) | Mehrpour et al. (2020) | Iran | Inpatient | Case series | 10 | 75.6 (9.6) | 5 (50%) | Ischaemic stroke 9 (90%), Haemorrhagic stroke 1 (10%) |
| (72) | Mei et al. (2020) | China | Inpatient | Case series | 494 | 40 /IQR 27-59) | 266 (53.8%) | Smell and Taste 39 (7.9%) |
| (73) | Brandstetter et al. (2020) | Germany | Healthcare workers | Cross-sectional | 31 | - | 5 (16%) | Headache 15 (48.4%), Myalgia 15 (48.4%) Fatigue 13 (41.9%) |
| (74) | Jäckel et al. (2020) | Germany | Inpatient | Case series | 20 | 65.5 (11.0) | 16 (80%) | Delirium 13 (65%) |
| (75) | Jalessi et al. (2020) | Iran | Post-discharge | Cross-sectional | 92 | 52.9 (13.3) | 62 (67.4%) | Headache 20 (21.7%), Myalgia 57 (62%), Smell and Taste 15 (16.3%), Impaired smell onset 22 (23.9%), Impaired smell follow-up 1 (1%) |
| (76) | Joffily et al. (2020) | Brazil | Outpatient | Cross-sectional | 159 | (70% under 39) | 50 (31.4%) | Smell and Headache 116 (73%), Impaired smell onset 159 (100%), Impaired smell follow-up 88 (55.3%), Impaired taste onset 147 (92.5%) |
| (77) | Kai Chua et al. (2020) | Singapore | Presenting to ED with acute respiratory symptoms | Case series | 31 | - | - | Impaired smell onset 7 (22.6%) |
| (78) | Kandemirli et al. (2020) | Turkey / USA | ICU | Case series | 235 | - | - | 50 (21%) developed neurological symptoms |
| (79) | Karadaş et al. (2020) | Turkey | Inpatient | Cohort | 239 | 46.5 (15.4) | 133 (55.7%) | Headache 64 (26.7%), Myalgia 36 (15.1%), Dizziness/vertigo 16 (6.7%), Tinnitus 5 (2.1%), Hearing impairment 3 (1.3%), Visual defect 8 (3.3%), Smell and Taste 11 (4.6%), Impaired smell onset 18 (7.5%), Impaired taste onset 16 (6.7%), Impaired consciousness/delirium/encephalopathy 23 (9.6%), Ischaemic stroke 7 (2.9%), Haemorrhagic stroke 2 (0.8%), Guillain-Barre syndrome 1 (0.4%), Sleep disorders 30 (12.6%) |
| (80) | Carignan et al. (2020) | Canada | Outpatient | Case-control | 134 | 57.1 (/IQR 41-65) | 64 (47.8%) | Headache 87 (64.9%), Myalgia 76. (56.7%), Asthenia 104 (77.6%), Dizziness/vertigo 27 (20.1%) Visual defect 6 (4.5%), Smell and/or Taste 87 (64.9%), Impaired smell onset 69 (51.5%), Impaired taste onset 85 (63.4%) |
| (81) | [Meini et al.](https://link.springer.com/article/10.1007%2Fs00405-020-06102-8) (2020) | Italy | Discharged from ITU 1 month ago | Cross-sectional | 100 | 65 (15) | 39 (39%) | Smell and Taste 28 (28%), Impaired smell onset 29 (29%), Impaired smell follow-up 5 (5%), Impaired taste onset 41 (41%), Impaired taste follow-up 7 (7%) |
| (82) | [Menni et al.](https://www.nature.com/articles/s41591-020-0916-2) (2) (2020) | UK / USA | App users | Cohort | 7178 | 41.6(-) | (-) | Smell and Taste 4668 (65%), Fatigue 2093(29.2%), Impaired consciousness/delirium/encephalopathy 1323 (18.4%) |
| (83) | [Korkmaz et al](https://www.ncbi.nlm.nih.gov/pmc/articles/PMC7324269/). (2020) | Turkey | Paediatric cases presenting to ED | Case series | 79 | 9.5 (IQR 3–15) | 48 (60.8%) | Headache 11(14%), Fatigue or Myalgia 15 (19%), Impaired smell onset 1 (1%) |
| (84) | Mercante et al. (2020) | Italy | Mixed | Cross-sectional | 204 | 52.6 (14.4) | 110 (53.9%) | Headache 32 (15.7%), Myalgia 20 (9.9%), Dizziness/vertigo 43 (21.1%), Fatigue 46 (22.6%), Impaired smell onset 85 (41.7%), Impaired taste onset 113 (55.4%) |
| (85) | Speth et al. (2020) | Switzerland | Mixed | Cross-sectional | 114 | 44.6 (16.1) | 52 (45.60%) | PHQ-2 and GAD-2 scores increased significantly from baseline (adjusted incidence rate ratio 1.40 [1.10 - 1.78], *p*=0.006) |
| (86) | Speth et al. (2) (2020) | Switzerland | Mixed | Cross-sectional | 103 | 46.8 (15.9) | 50 (48.5%) | Impaired smell onset 63 (61.2%), Impaired tastel onset 67 (65%) |
| (87) | Kremer et al. (2020) | France | Inpatient | Case series | 37 | 61 (12) | 30 (81.0%) | Headache 4 (11%), Focal neurological deficit 4 (11%), Seizures 5 (14%), Impaired consciousness/delirium/encephalopathy 27 (73%), Haemorrhagic stroke 11 (29.7%), Other cerebrovascular accident 9 (24%), Sleep disorders 15 (41%), Post ICU neuropsychiatric abnormality 15 (41%) |
| (88) | Spoldi et al. (2020) | Italy | Inpatient undergoing head CT | Cross-sectional | 63 | 77.8 (17.8) | 39 (61.9%) | Olfactory cleft opacification 16 (25%) |
| (89) | Sun et al. (2020) | China | Mixed | Case series | 55 | 44 (IQR 34-56) | 31 (56.4%) | Headache 6 (10.9%), Myalgia 10 (18.2%), Dizziness/Vertigo 6 (10.9%) |
| (90) | [Sweid et al.](https://covid19.elsevierpure.com/en/publications/cerebral-ischemic-and-hemorrhagic-complications-of-coronavirus-di) (2020) | USA | Acute cerebrovascular disease | Case series | 22 | 59.5 (16) | 10 (45.4%) | Ischaemic stroke 19 (86.4%), Haemorrhagic stroke 3 (13.6%) |
| (91) | Chary et al. (2020) | France | Mixed | Cohort | 115 | 47 (-) | 34 (29.6%) | Headache 62 (54%), Myalgia 57 (50%), Smell and Taste 38 (33%), Impaired smell onset 81 (70%), Impaired smell follow-up 29 (25.2%) |
| (92) | Lapostolle et al. (2020) | France | Outpatient | Cross-sectional | 1487  (data missing for 35) | 44(M),41 (F)*  IQR 32-57 (M)  IQR 30-54 (F) | 700 (47.7%) | Headache 803 (55.3%), Myalgia 823 (6.7%), Asthenia 864 (59.5%), Impaired smell onset 406 (28%), Impaired taste onset 414 (28.5%) |
| (93) | Lechien et al. (2020) | Belgium | General population | Cross-sectional | 78 | 40.6 (11.2) | 32 (41.0%) | Impaired smell onset 78 (100%), Impaired taste onset 53 (67.9%) |
| (94) | Lechien et al. (2) (2020) | Belgium | Outpatient | Cross-sectional | 16 | 36 (10) | 8 (50%) | Headache 5 (31.3%), Myalgia 4 (25%), Asthenia 7 (44%) |
| (95) | Lechien et al. (3) (2020) | Europe | Mixed | Cross-sectional | 702 | 40.3 (11.8) | 206 (29.3%) | Headache 447 (63.7%), Myalgia 371 (52.9%), Asthenia 519 (73.9%) |
| (96) | Lechien et al. (4) (2020) | Europe | Mixed | Cross-sectional | 1420 | 39.2 (12.1) | 458 (32.3%) | Headache 998 (70.3%), Myalgia 887 (62.5%), Asthenia 514 (36 %), Smell and Taste 875 (61.6%), Impaired smell onset 997 (70.2%), Impaired smell follow-up 429 (30.2%), Impaired taste onset 770 (54.2%), Impaired taste follow-up 332(23.4%) |
| (97) | Lechien et al. (5) (2020) | Europe | Mixed | Cross-sectional | 2013 | 39.5 (12.1) | 684 (34.0%) | Headache 1411 (70.1%), Myalgia 1244 (61.8%), Impaired smell onset 1751 (87%), Impaired smell follow-up 282 (14%), Impaired taste onset 1135 (56.4%) |
| (98) | Lechien et al. (6) (2020) | Belgium | Outpatient | Cross-sectional | 86 | 41.7 (11.8) | 30 (34.9%) | Headache 42 (60%), Myalgia 30 (42.9%), Asthenia 51 (72.9 %), Speech/aphasia 17 (24.3%), Visual defects 18 (25.7%), Fatigue 51 (73%), Impaired smell onset 86 (100%), Impaired taste onset 44 (51%) |
| (99) | Khan et al. (2020) | UAE | Inpatient | Case series | 22 | 46.3 (11.1) | 20 (90.9%) | Ischaemic stroke 22 (100%) |
| (100) | Khira et al. (2020) | USA | Inpatients who had CT angiogram | Case series | 31 | 64.9 (15.7) | 68 | Stroke 31 (100%) |
| (101) | Lee et al. (2020) | Canada | Inpatient | Case-control | 56 | 38 (/IQR 32–53) | 23 (41.10%) | Headache 10 (17.9%), Smell and Taste 20 (35.7%), Fatigue 4 (7.1%), Impaired smell onset 31 (55.4%), Impaired taste onset 32 (57.1%) |
| (102) | Lee et al. (2) (2020) | USA | General population | Cross-sectional | 61 | - | - | Dysfunctional coronavirus anxiety associated with coronavirus infection (OR 3.04 [1.28-7.25]) |
| (103) | Lee et al. (3) (2020) | Korea | Inpatients | Case series | 98 | 72* (IQR 68-79) | 44 (44.9%) | Focal neurological deficit 7 (9%) |
| (104) | Lee et al. (4) (2020) | Korea | Mixed | Cohort | 3,191 | 44 (e/IQR 25.–58.) | 1,161 (36.4%) | Smell and Taste 488 (15.3%) |
| (105) | Li et al. (2020) | China | Inpatient | Case series | 219 | 53.3 (15.9) | 89 (40.6%) | Ischaemic stroke 10 (4.6%), Haemorrhagic stroke 1 (0.5%) |
| (106) | Merkler et al. (2020) | USA | Mixed | Case series | 1916 | 64* (IQR 51-76) | 1101 (57.5%) | Ischaemic stroke 31 (1.6%) |
| (107) | Moein et al. (2020) | Iran | Inpatient | Case-control | 60 | 46.6 (12.2) | 40 (66.7%) | Headache 22 (37%), Myalgia 5 (8%),Tinnitus 1 (2%), Smell OR Taste 21 (35%), Impaired smell onset 59 (96.3%) , Impaired taste onset 4 (7%), |
| (108) | Moro et al. (2020) | Italy | Online- survey of neurologists | Cross-sectional | - | - | - | Most frequently reported neurological findings: headache (61.9%), myalgia (50.4%), anosmia (49.2%), ageusia (39.8), impaired consciousness (29.3%), psychomotor agitation (26.7%), encephalopathy (21.0%), acute cerebrovascular disorders (21.0%) |
| (109) | Nalleballe et al. (2020) | USA | Mixed | Cohort | 40,469 | - | 18,364 (45.4%) | Headache 150 (3.7%), Myalgia 821 (2%), Dizziness/vertigo 379 (0.9%), Focal neurological deficit 392 (1%), Seizures 258 (0.6%), Smell and Taste 477 (1.2%), Depression 1549 (3.8%), Anxiety 1869 (4.6%), Impaired consciousness/delirium/encephalopathy 937 (2.3%), Other cerebrovascular accident 406 (1.%), Sleep disorders 1394 (3.4%), Other movement disorders 277 (0.7%), Suicidal ideation 63(0.2%) |
| (110) | Li et al. (2020) | China | Inpatient | Cross-sectional | 280 | 104 (51-65) | 135 (48.2%) | Fatigue 172 (61.4%), Depression 114 (40.7%), Anxiety 174 (62.1%), Sleep disorders 178 (63.6%) |
| (111) | Li et al. (2) (2020) | China | Inpatient | Case series | 93 | 51 (17.5) | 41 (44.1%) | Myalgia 34 (37%), Fatigue 63 (68%) |
| (112) | Li et al. (3) (2020) | China | Inpatient | Case series | 83 | 45.5 (12.3) | 44 (53.0%) | Headache 9 (10.8%), Myalgia 15 (18.1%) |
| (113) | Paterson et al. (2020) | UK | Inpatient | Case series | 43 | Summarised according to clinical presentation) Encephalopathy: 57.5, inflammatory CNS syndromes: 53, stroke: 62.5, GBSL 56, plexopathy: 60, miscellaneous and uncharacterised: 20 | Reported % male: Encephalopathy: 40, inflammatory CNS syndromes: 33, stroke: 75, GBS: 100, plexopathy: 100, miscellaneous and uncharacterised: 40 | Encephalopathy 10 (23%), inflammatory CNS syndromes 12 (28%), ischaemic stroke 8 (19%), peripheral neurological disorders 8 (19%), miscellaneous CNS disorders 5 (12%) |
| (114) | D'Ascanio et al. (2020) | Italy | Mixed | Case-control | 43 | 58.1 (15.6) | 29 (67.4%) | Impaired smell onset 26 (60.4%) |
| (115) | Dawson et al. (2020) | USA | Household study of patients with positive swabs and those they lived with | Cross-sectional | 42 | 31* (Range <1 - >90) | 69 (-) | Headache 32 (76%), Myalgia 24 (57%), Light-headed 3 (7%), Smell OR Taste 16 (38%), Fatigue 15 (36%), Impaired smell onset 18 (43%), Impaired taste onset 24 (57%) |
| (116) | Dell'Era et al. (2020) | Italy | Outpatient | Cross-sectional | 355 | 50* (IQR 40-59.5) | 192 (54.1%) | Fatigue 143 (40.3%), Impaired smell onset 237 (66.8%) , Impaired smell follow-up 88 (24.8%), Impaired taste onset 232 (65.4%), Impaired taste follow-up 84 (36.2%) |
| (117) | Tabata et al. (2020) | Japan | Inpatient | Case series | 104 | 68*(IQR 47-75) | 54 (51.9%) | - |
| (118) | Tian et al. (2020) | China | Inpatient | Case series | 751 | 64 (IQR 57-69) | 372 (49.5%) | Headache 37 (5%) |
| (119) | Tostmann et al. (2020) | Netherlands | Healthcare workers | Cross-sectional | 90 | - | 19 (21.1%) | Headache 64 (71.1%), Myalgia 57 (63.3%), Impaired smell onset 37 (46.8%) |
| (120) | Du et al. (2020) | China | Inpatient | Case series | 12 | 45.3 (Range 23-79) | 7 (58.3%) | Headache 3 (25%), Fatigue 10 (83%) |
| (121) | Du W et al (2020) | China | Inpatient | Case series | 67 | 34.1* (0-65) | 32 (47.8%) | Headache 1 (7.1%), Myalgia 1 (7.1%), Fatigue 1 (7.1%) |
| (122) | Trubiano et al. (2020) | Australia | Outpatient | Cross-sectional | 28 | 55* (IQR 46-64) | 14 (50%) | Headache 6 (21.4%), Myalgia 15 (53.6%), Smell and Taste 3 (10.7%), Impaired smell onset 7(25%) |
| (123) | Guan et al. (2020) | China | Inpatient | Case series | 1099 | 47 (IQR 35-58) | 637 (58%) | Headache 150 (13.6%), Myalgia 164 (14.9%), Fatigue 419 (38.1%) |
| (124) | Escalard et al (2020) | France | Inpatient | Case series | 10 | 59.5 (IQR 54-72) | 8 (80%) | Ischaemic stroke 10 (100%) |
| (125) | Farahani et al. (2020) | Iran | Inpatient | Cohort | 10 | 50* (-) | 7 (70%) | Headache 10 (100%), Fatigue 10 (100%) |
| (126) | Freni et al. (2020) | Italy | Outpatient | Cohort | 50 | 37.7 (17.9) | 30 (60.0%) | Headache 24 (48%), Myalgia 18 (36%), Asthenia 25 (50%), Impaired smell onset 46 (92%), Impaired smell follow-up 9 (18%), Impaired taste onset 35 (70%), Impaired taste follow-up 4 (8%) |
| (127) | Galanopoulou et al. (2020) | USA | Inpatient | Case series | 22 | 63.2 (11.9) | 14 (63.6%) | Seizures 14 (63.6%), Impaired consciousness/delirium/encephalopathy 15 (68.2%) |
| (128) | Tudrej et al. (2020) | France | Outpatient | Cross-sectional | 198 | 45*(IQR 28) | 284 (143.0%) | Smell and Taste 58 (29.3%), Impaired smell onset 82 (41.4%), Impaired taste onset 92 (46.5%) |
| (129) | Vacchiano et al. (2020) | Italy | Inpatient | Cohort | 108 | 59*(Range 18-83) | 62 (57%) | Headache 46 (43%), Myalgia 37 (34%), Dizziness/vertigo 11 (10%), Impaired smell onset 40 (37%), Impaired taste onset 66 (61%) |
| (130) | Vaira et al. (2020) | Italy | Mixed | Cross-sectional | 345 | 48.5 (12.8) | 146 (42.3%) | Impaired smell onset 241(70%), Impaired taste onset 155 (44.9%) |
| (131) | Vaira et al. (2020) | Italy | Inpatient | Cross-sectional | 72 | 49.2 (13.7) | 27 (37.5%) | Headache 30 (41.6%), Asthenia 48 (66.7%), Smell and Taste 30 (41.7%), Impaired smell onset 14 (14.4%), Impaired taste onset 9 (12.5%), Impaired taste follow-up 9 (12.5%) |
| (132) | Boscolo-Rizzo et al. (2020) | Italy | Self-isolating | Cross-sectional | 54 | - | - | Smell or Taste 34 (63%) |
| (133) | Burke et al. (2020) | USA | Mixed | Case series | 164 | 50 (-) | 92 (56%) | Headache 97 (59%), Myalgia 103 (63%), Smell or Taste 36 (24%), Fatigue 102 (62%) |
| (134) | Cecchetti et al. (2020) | Italy | Inpatients referred for EEG | Case series | 18 | - | 11 (61.1%) | Seizures 0 (0%) |
| (135) | Chen et al. (2020) | China | Inpatient | Case series | 12 | 14.5 (IQR 9--16) | 6 (50.0%) | Dizziness/vertigo 2 (16.7%), Fatigue 1 (8.3%) |
| (136) | Chen et al. (2) (2020) | China | Inpatient | Case series | 113 | 68 (IQR 62-77) | 83 (73.5%) | Headache 11 (10%), Myalgia 21 (19%), Dizziness/vertigo 10 (9%), Fatigue 64 (57%), Impaired consciousness/delirium/encephalopathy 25 (22%) |
| (137) | Denis et al. (2020) | France | Other | Cross-sectional | 2,477,174 | 39.1, 37*, (IQR 15-99) | - | Asthenia 1,154363 (46.6%), Smell or Taste 325,910 (17.1%) |
| (138) | Duan et al. (2020) | China | Inpatient | Case series | 25 | 52 (19.3) | 15 (60.0%) | Fatigue 14 (56%) |
| (139) | Fraisse et al. (2020) | France | Inpatient (ICU only) | Case series | 92 | 61*(IQR 55-70) | 73 (79.3%) | Ischaemic stroke 2 (2.2%), Haemorrhagic stroke 2 (2.2%) |
| (140) | Gaborieau et al. (2020) | France | Inpatient | Cohort | 157 | 0.5*(IQR 0.125 – 10) | 94 (59.9%) | Smell or Taste 7 (4.5%) |
| (141) | Garrazzino et al. (2020) | Italy | Mixed | Case series | 168 | 5 (IQR 0.3-9.6) | 94 (56.0%) | Seizures 3 (1.8%), Fatigue 3 (1.8%) |
| (142) | Guo et al. (2020) | China | Inpatient | Case-control | 103 | 42.5 (12.5) | 59 (57.3%) | Depression 62 (60.2%), Anxiety 59 (55.3%), PTSD 1 (10%) |
| (143) | Kaye et al. (2020) | USA | Other | Cross-sectional | 237 | 39.6 (14.6) | 108 (45.6%) | Headache 88 (37%), Impaired smell onset 173 (73%), Impaired smell follow-up 65 (27%) |
| (144) | Li et al. (2020) | China | Inpatient | Case series | 655 | - | 367 (56.0%) | Headache 80 (12.2%), Myalgia 78 (11.9%), Fatigue 184 (28.1%) |
| (145) | Liang et al. (2020) | China | inpatients | Cross-sectional | 86 | 25.5 (Range 6-57) | 44 (51.2%) | Headache 12 (14%), Myalgia 8 (9.3%), Tinnitus 3 (3.5%), Smell and Taste and Tinnitus 44 (51.2%), Fatigue 16 (18.6%), Impaired smell onset 34 (39.5%), Impaired taste onset 33 (38.4%) |
| (146) | Liu et al. (2020) | Taiwan | Imported cases from abroad | Case series | 321 | - | 151 (47%) | Headache 34 (10.6%), Myalgia 14 (12.5%), Dizziness/vertigo 6 (1.9%), Smell or Taste 42 (13.1%), Fatigue 52 (16%), Ophthalmic symptoms 6 (1.9%) |
| (147) | Luo et al. (2020) | China | Mixed | Case series | 60 | 43.6 (M) (2.7) 50.0 (F) (1.8) | 32 (53.3%) | Headache 8 (12.7%), Dizziness/vertigo 12 (20%), Fatigue 23 (38%), Impaired taste onset 8 (12.7%) |
| (148) | Mao et al. (2020) | China | Inpatients | Case series | 214 | 52.7 (15.5) | 87 (40.7%) | Headache 28 (13.1%), Dizziness/vertigo 36 (16.8%), Ataxia 1 (0.5%), Seizures 1 (0.5%), Neuralgia 5 (2.3%), Visual defects 3 (1.4%), Impaired smell onset 11 (5.1%), Impaired taste onset 12 (5.6%), Impaired consciousness 16 (7.5%), Other cerebrovascular accident 6 (2.8%), Myopathy 23 (10.7%) |
| (149) | Myrstad et al. (2020) | Norway | Inpatients | Cohort | 66 | 67.9 (Range 30-95) | 38 (57.6% | Impaired consciousness/delirium/encephalopathy 11 (16.7%), New confusion 11 (16.7%) |
| (150) | Naeini et al. (2020) | Iran | Mixed | Cross-sectional | 49 | 45.1 (12.2) | 27 (55.1%) | Headache 35 (71.4%), Fatigue 31 (63%), Impaired smell onset 49 (100%), Impaired taste onset 37 (75.5%) |
| (151) | Nguyen et al. (2020) | Vietnam | Outpatient | Cross-sectional | 1387 | - | - | Participants with COVID-19 more likely to have depression (OR 2.88, *p*<0.001) and lower average HRQOL (*p*<0.001) |
| (152) | Pati et al. (2020) | USA | Inpatient (severe) | Case series | 10 | 61.3 (-) | 5 (50.0%) | Cerebral Performance Category <2 (poor outcome): 5 (50%); 3-5 (good outcome): 5 (50%) |
| (153) | Paz et al. (2020) | Ecuador | Outpatient | Cross-sectional | 306 | 38.3 (11.0) | 149 (48.7%) | Depression and anxiety 49 (16%), Depression 17 (22.9%), Anxiety 74 (24.2%) |
| (154) | Petrescu et al. (2020) | France | Inpatients undergoing EEG | Case series | 36 | 69.1 (-) | 26 (72.2%) | Confusion or fluctuating alertness 23 (58%), delayed awakening after ceasing sedation 6 (15%) |
| (155) | Petrocelli et al. (2020) | Italy | Healthcare workers | Cohort | 300 | 43.6 (12.2) | 75 (25.0%) | Headache 133 (44.3%), Myalgia 128 (42.7%), Asthenia 109 (36.3%), Smell and Taste 164 (54.7%), Impaired smell onset 26 (8.7%), Impaired taste onset 20 (6.7%) |
| (156) | Peyrony et al. (2020) | France | Inpatient | Cohort | 225 | 62 (whole population) (IQR 48-71) | 241 (107%) | Headache 15 (6.7%), Myalgia 71 (31.6%), Dizziness/vertigo 8 (3.6%), Fatigue 34 (15.1%), Impaired smell onset 31 (13.8%), Impaired consciousness/delirium/encephalopathy 15 (6.7%) |
| (157) | Pinna et al. (2020) | USA | Inpatient | Case series | 50 | 59.6 (-) | 29 (58.0%) | Headache 12 (24%), Ataxia 1 (2%), Palsy 3 (6%), Paraesthesia 1 (2%), Seizures 13 (26%), Hypogeusia and Dysgeusia 5 (10%), Impaired smell onset 3 (6%), Altered mental status 30 (60%), Ischaemic stroke 10 (20%) , Haemorrhagic stroke 4 (8%), SAH 4 (8%), TIA 1 (2%), Hypoxic ischemic brain injury 7 (14%), Meningoencephalitis 2 (4%), Myopathy 6 (12%), Short term memory loss 12 (24%), Extra -ocular muscle abnormalities 5 (10%) |
| (158) | Pons-Escoda et al. (2020) | Spain | Inpatient | Case series | 103 | 74 (IQR 50-90) | 63 (61.2%) | Seizures 3 (2.9%), Ischaemic stroke 3 (2.9%), Haemorrhagic stroke 8 (7.7%), Hematoma 6 (5.8%), Guillain-Barre syndrome 1 (1%) |
| (159) | Porta-Etessam et al. (2020) | Spain | Healthcare workers previously infected | Cross-sectional | 112 | 43.4 (11.4) | 21 (18.8%) | Headache 112 (100%), Photophobia 32 (28.6%), Impaired smell onset 11 (9.8%) |
| (160) | Qin et al. (2020) | China | Inpatient | Case series | 1875 | 63 (IQR 51-70) | 945 (50.4%) | Headache 123 (6.6%) , Myalgia 259 (13.8%), Dizziness/vertigo 87 (4.6%), Fatigue 477 (25.4%), Impaired consciousness/delirium/encephalopathy 16 (0.9%), Phonophonbia 46 (41.1%), Allodynia 5 (4.5%), Autonomic Symptoms 22 (19.6%) |
| (161) | Qiu et al. (2020) | China, France, Germany | Inpatient | Case series | 161 | 38.8 (IQR 23-53) | 92 (57.1%) | Smell and Taste 93 (24%), Impaired smell onset 61 (15%), Impaired taste onset 7 (2%) |
| (162) | Radmanesh et al. (2020) | USA | Inpatient | Case series | 11 | 53 (Range 38-64) | 9 (81.8%) | Impaired consciousness/delirium/encephalopathy 11 (100%) |
| (163) | Reddy et al. (2020) | USA | Inpatient | Case series | 12 | 56.3 | 6 (50.0%) | Ischaemic stroke 10 (83.3%), Haemorrhagic stroke 2 (16.7%) |
| (164) | Shi et al. (2020) | China | Inpatient (ICU only) | Case series | 161 | 59.4 (16.6) | 104 (64.6%) | Headache 5 (3.1%), Fatigue and Myalgia 17 (10.6%) |
| (165) | Soltani et al. (2020) | Iran | Inpatient (Paeds) | Cohort | 30 | 6 (5) | 14 (46.7%) | Impaired consciousness/delirium/encephalopathy 4 (13.3%) |
| (166) | Spinato et al. (2020) | USA | Outpatients (mild) | Cross-sectional | 202 | 56 (IQR 45-67) | 97 (48.0%) | Headache 86 (42.6%), Myalgia 90 (44.6%), Smell or Taste 130 (64.4%), Fatigue 138 (68.3%) |
| (167) | [Su et al.](https://pubmed.ncbi.nlm.nih.gov/32208917/) (2020) | - | Inpatients (Resp sx) | Case series | 14 | 42.9 (Range 30-72) | 8 (57.1%) | Fatigue 3 (21.4%) |
| (168) | Sun et al. (2020) | China | Inpatients (>60) | Case-control | 244 | Discharged: 67* (IQR 64-72)  Deceased: 72*  (IQR 66-78) | 51 (41.5%) discharged  82 (67.8%) deceased | Consciousness disorders 32 (13.1%) |
| (169) | Tenforde et al. (2020) | USA | Mixed | Cross-sectional | 350 | 43* (IQR 32-57) | 165 (47.1%) | Headache 171 (60%) Myalgia 167 (58%), Smell and/or Taste 163 (56%), Fatigue 198 (69%), Impaired smell onset 140 (49%), Impaired taste onset 143 (50%), Confusion 41 (14%) |
| (170) | Vaira et al. (2020) | Italy | Healthcare workers | Cross-sectional | 33 | 51.8 (-) | 11 (33.3%) | Smell and Taste 13 (39.4%), Impaired smell onset 17 (51.5%), Impaired taste onset 17 (51.5%) |
| (171) | Varatharaj et al. (2020) | UK | Inpatient | Case series | 153 | 71 (Range 23-94) | 73 (47.7%) | Focal neurological deficit 1 (0.8%), Seizures 1 (0.8%), Cognitive impairment 6 (4.8%), Depression 3 (2.4%), Impaired consciousness/delirium/encephalopathy 9 (7.2%), Ischaemic stroke 57 (45.65), Haemorrhagic stroke 9 (7.2%), Other cerebrovascular accident 11 (8.8%), Meningoencephalitis 7 (5.6%), Guillain-Barre syndrome 4 (3.2%), Catatonia 1 (0.8%), Mania 1 (0.8%), Psychosis 10 (8%), Myasthenic crisis 1 (0.8%) |
| (172) | Vargas-Gandica et al. (2020) | Germany, USA,  Venezuela, Bolivia | Mixed | Case series | 10 | 48 (-) | 3 (30.0%) | Headache 3 (30%), Weakness 4 (40%), Impaired smell onset 8 (80%), Impaired taste onset 9 (90%), Polyarthralgia 2 (20%) |
| (173) | Vespignani et al. (2020) | France | Inpatient | Case series | 26 | Specified for only 5/26 patients:  67 (-) | Specified for only 5/26 patients:  4 (15.4%) | Seizures 1 (20%), Impaired smell onset 1 (20%) |
| (174) | Wang et al. (2020) | China | Inpatients | Cohort | 131 | 49* (Range 18-88) | 59 (45%) | Dizziness/vertigo 2 (1.5%), Fatigue 10 (7.6%) |
| (175) | Wu et al. (2020) | France | Inpatient | Cross-sectional | 14 | - | 7 (50.0%) | Fatigue 8 (57.1%), Anxiety 13 (92.9%) |
| (176) | Xiao et al. (2020) | China | Inpatient | Cross-sectional | 66 | - | 34 (51.5%) | Headache 11 (16.7%) Myalgia 16 (24.2%), Fatigue 26 (39.3%), Ischaemic stroke 5 (3.3%) |
| (177) | Xie et al. (2020) | China | Inpatient | Case series | 21 | 54 (15.4) | 13 (61.9%) | Myalgia 1 (4.8%), Weakness 4 (19%) |
| (178) | Xiong et al. (2020) | China | Inpatient | Cohort | 917 | 48.7 (17.1) | 504 (55.0%) | Headache 2 (0.2%), Myalgia 2 (0.2%), Syncope 3 (0.3%), Neuralgia 1 (0.1%), Delirium 7 (0.8%), Other cerebrovascular accident 10 (1.1%) |
| (179) | Yan et al. (2020) | USA | Mixed | Cross-sectional | 59 | - | 29 (49.2%) | Headache 39 (66.1%), Myalgia 37 (62.7%), Fatigue 48 (81.4%), Impaired smell onset 13 (22%), Impaired smell follow-up 40 (67.8%), Impaired taste onset 12 (20.3%), Impaired taste follow-up 42 (71.2%) |
| (180) | Yan et al. (2) (2020) | USA | Mixed | Cross-sectional | 316 | - | - | Impaired smell onset 23 (7.3%), Impaired smell follow-up 18 (23%) |
| (181) | Yang et al. (2020) | China | Inpatient | Case series | 136 | 56 (/IQR 44-64) | 66 (48.5%) | Dizziness/vertigo 12 (8.8%), Fatigue 53 (39%), Sleep disorders 49 (36%) |
| (182) | Yin et al. (2020) | China | Mixed | Cohort | 30 | 52.7 (15.1) | 19 (63.3%) | Headache 5 (16.7%), Myalgia 4 (13.3%), Fatigue 8 (26.7%) |
| (183) | Yu et al. (2020) | China | Inpatient | Case series | 1859 | 59 (IQR 45-68) | 934 (50.2%) | Headache 107 (6%), Myalgia 315 (17%), Fatigue 695 (37%) |
| (184) | Yu et al. (2020) | China | Inpatient | Case series | 129 | 64 (/IQR 56-69) | 56 (43.4%) | Myalgia 25 (19.4%), Fatigue 39 (30.2%) |
| (185) | Yuan et al. (2020) | China | Self-isolating | Cross-sectional | 96 | 47.1 (13.2) | 47 (49.0%) | Depression 42 (43.8%) |
| (186) | Zarghami et al. (2020) | Iran | Mixed | Cross-sectional | 82 | 40.3 (14.4) (Hospitalised)  43.6 (15.8) (Non-Hospitalised) | 32 (39.0%) | Depression 3 (3.7%), Anxiety 5 (6.1%), Sleep disorder 24 (29.3%) |
| (187) | Zayet et al. (2020) | France | Mixed | Cross-sectional | 70 | 56.7 (19.3)  (Range/19-96) | 29 (41.4%) | Headache 51 (72.9%), Myalgia 41 (58.6%), Tinnitus 7 (10%), Hearing loss 4 (5.7%), Blurred vision 3 (4.3%), Fatigue 65 (92.9%), Impaired smell onset 37 (52.9%), Impaired taste onset 34 (48.6%) |
| (188) | Zayet et al. (2) (2020) | France | Outpatient | Case series | 95 | 39.8 (12.2)  (Range 18-73) | 16 (16.8%) | Headache 74 (77.7%), Myalgia 71 (74.7%), Smell and/or Taste 70 (73.7%), Impaired smell onset 60 (63.2%), Impaired taste onset 62 (65.3%) |
| (189) | Zhang et al. (2020) | China | Mixed | Cohort | 221 | 55*(IQR 39-67) | 108 (48.9%) | Headache 17 (7.7%), Fatigue 156 (70.6%) |
| (190) | Zhang et al. (2) (2020) | China | Inpatient | Cohort | 194 | 48.3*(IQR 33-56) | 108 (55.7%) | Headache 33 (17.0%), Myalgia 44 (22.7%) |
| (191) | Zhang et al. (3) (2020) | China | Inpatient | Case series | 869 | 51* (IQR 40-58) | 377 (43.4%) | Headache 18 (2.1%), Myalgia 114 (13.1%), Dizziness/vertigo 12 (1.4%), Fatigue 226 (26%) |
| (192) | Zhao et al. (2020) | China | Inpatient | Cohort | 29 | 56 (IQR 32 -66) | 14 (48.3%) | Headache 3 (10.3%), Myalgia 3 (10.3%), Fatigue 8 (27.6%) |
| (193) | Zhao et al. (2) (2020) | China | Inpatient | Case series | 19 | 48*(IQR 27-56) | 11 (57.9%) | Headache 2 (10.5%), Fatigue 2 (10.5%) |
| (194) | Zheng et al. (2020) | China | Inpatient | Cohort | 161 | 45*(IQR 33.5-57) | 80 (49.7%) | Headache 12 (7.5%), Myalgia 18 (11.2%), Fatigue 64 (39.8%) |
| (195) | Zheng et al. (2) (2020) | China | Inpatient | Case series | 34 | 66*(IQR 58-76) | 23 (67.6%) | Headache 2 (5.9%), Myalgia 5 (14.7%), Fatigue 2 (5.9%) |
| (196) | Zhou et al. (2020) | China | Inpatient | Case-control | 29 | 47 (10.5) | 18 (62.1%) | No significant differences between total scores on Trail Making Test, Sign Coding Test, Continuous Performance Test and Digital Span Test among recovered COVID-19 patients compared to healthy controls. |
| (197) | Zhou et al. (2) (2020) | China | Inpatient | Cohort | 21 | 66.1 (13.9) | 13 (61.9%) | Myalgia 2 (9.5%), Fatigue 5 (23.8%) |
| (198) | Mahammedi et al. (2020) | Italy | Inpatient | Case series | 108 | 71*(IQR 60.5-79) | 69 (63.9%) | Headache 13 (12%), Myalgia 13 (12%), Ataxia 2 (2%), Seizures 10 (9%), Neuralgia 3 (3%), Impaired smell onset 2 (2%), Impaired consciousness/delirium/encephalopathy 64 (59%), Ischaemic stroke 34 (31%), Haemorrhagic stroke 6 (5.6%), Guillain-Barre syndrome 2 (20%), Central demyelination 2 (10%) |
| (199) | Solomon et al. (2020) | USA | Inpatient | Case series | 18 | 62 (IQR 53-75) | 14 (77.8%) | Headache 2 (11.1%), Myalgia 3 (16.7%), Impaired taste onset 1 (5.5%) |
| (200) | Levinson et al. (2020) | Israel | Isolation unit | Cross-sectional | 45 | 34 (Range 15-82) | 23 (51.1%) | Headache 20 (48%), Myalgia 24 (57%), Dizziness/vertigo 9 (21%), Fatigue 29 (69%), Impaired smell onset 15 (36%), Impaired smell follow-up 4 (8.9%), Impaired taste onset 14 (33%), Impaired taste follow-up 2 (2.2%) |
| (201) | Kotabagi et al. (2020) | UK | Inpatient (Pregnant women) | Cross-sectional | 11 | 31(Range 18-39) | 0 (0.0%) | PHQ-9 and GAD-7 scores highest at time of institution of lockdown rules |
| (202) | Yaghi et al. (2020) | USA | Inpatient | Case series | 3556 | 62.5 (IQR 52-69.3) (in events) | 23 (71.9%) (in events) | Stroke 32 (0.9%) |
| (203) | Yan et al. (2020) | USA | Mixed | Case series | 128 | 53.5* (IQR 40-65) | 9 (7.0%) | Headache 62 (48.4%), Fatigue 90 (70.3%), Impaired smell onset 75 (58.6%), Impaired taste onset 70 (54.7%) |
| (204) | Tian et al. (2020) | China | Inpatient | Case series | 262 | 47.5*(Range 1-94) | 127 (48.5%) | Headache 17 (6.5%), Fatigue 69 (26.3%) |
| (205) | Helms et al. (2020) | France | Inpatient (with ARDS) | Case series | 58 | 63*(-) | - | Focal neurological deficit 39 (67%), Cognitive impairment 14 (36%), Confusion 26 (44.8%), Ischaemic stroke 3 (5%) |
| (206) | Suleyman et al. (2020) | USA | Mixed | Case series | 463 | 57.5 (16.8) | 204 (44.1%) | Headache 74 (16%), Myalgia 194 (42%) |
| (207) | Zier et al. (2020) | USA | Inpatient | Case series | 66 | 58*(Range 23-87) | 43 (65.2%) | Myalgia 36 (55%), Fatigue 44 (67%) |
| (208) | Dogra et al. (2020) | USA | Inpatient (ICH) | Case series | 33 | 62*(Range 37-83) | 26 (78.8%) | Speech/aphasia 1 (3%), Focal neurological deficit 7 (21.2%), Seizures 2 (6.1%), Impaired consciousness/delirium/encephalopathy 17 (51.1%), Haemorrhagic stroke 31 (93.9%), Other cerebrovascular accident 11 (33.3%), Meningoencephalitis 8 (24.2%) |
| (209) | Lechien et al. (2020) | Europe | Mixed | Cross-sectional | 417 | 36.9 (11.4) | 154 (36.9%) | Headache 188(45%), Myalgia 242 (58%), Asthenia 188 (45%), Impaired smell onset 284 (79.6%), Impaired taste onset 342 (88.8%) |
| (210) | Parma et al. (2020) | USA | Outpatients | Cross-sectional | 4039 | 41.4 (12.2) | 1118 (27.7%) | Mean reductions in smell (-79.7±28.7), taste (-69.0±32.6) and chemesthetic function (-37.3±36.2) during COVID-19 |
| (211) | De Maria et al. (2020) | Italy | outpatients | Cross-sectional | 95 | - | - | Impaired smell onset 48 (50.5%), Impaired taste onset 48 (50.5%) |
| (212) | Ligouri et al. (2020) | Italy | Inpatient | Cross-sectional | 103 | 55 (14.7) | 59 (57.3%) | Headache 40 (38.3%), Myalgia 25 (24.7%), Dizziness/vertigo 27 (26.2%), Hearing impairment 2 (1.9%), Paraesthesia 6 (5.8%), Fatigue 33 (32%), Impaired smell onset 40 (38.3%), Impaired taste onset 48 (46.6%), Depression 39 (37.9%), Anxiety 34 (33%), Impaired consciousness/delirium/encephalopathy 23 (22.2%), Sleep disorders 51 (49.5%) |
| (213) | Wee et al. (2020) | Singapore | Mixed | Cohort | 154 | - | - | Smell or Taste 35 (22.7%) |
| (214) | Giacomelli et al. (2020) | Italy | Inpatient | Cross-sectional | 59 | 60* (IQR 50-74) | 40 (67.8%) | Headache 2 (3.4%), Asthenia 1 (1.7%), Smell and Taste 11 (28.9%), Impaired smell onset 3 (5.1%), Impaired taste onset 6 (10.2%) |
| (215) | Gelardi et al. (2020) | Italy | Outpatient | Cohort | 72 | 49.7 (Range 19-70) | 39 (54.2%) | Headache 16 (22%) , Myalgia 13 (18%), Weakness 29 (40%), Impaired smell onset 26 (36.1%), Impaired taste onset 42 (58.3%) |

Outcomes refer to all COVID-19 infected patients. Mean age given, where median age used, illustrated by *, IQR/Range specified.

HCW = Health Care Workers. ICH = intracranial haemorrhage. ICU = Intensive Care Unit. MRI = Magnetic resonance imaging. ED = Emergency Department. CT = Computed tomography. CT Angiogram = Computed tomography angiography. SAH = Subarachnoid haemorrhage. TIA = Transient ischaemic attack. CNS = Central Nervous System.

REFERENCES:

1.	Nie X-D, Wang Q, Wang M-N, Zhao S, Liu L, Zhu Y-L, et al. Anxiety and depression and its correlates in patients with coronavirus disease 2019 in Wuhan. Int J Psychiatry Clin Pract. 2020;(9709509):1–6.

2.	Radmanesh A, Raz E, Zan E, Derman A, Kaminetzky M. Brain Imaging Use and Findings in COVID-19: A Single Academic Center Experience in the Epicenter of Disease in the United States. AJNR Am J Neuroradiol. 2020;41(7):1179–83.

[3.	Wei Y., Lu Y., Xia L., Yuan X., Li G., Li X., et al. Analysis of 2019 novel coronavirus infection and clinical characteristics of outpatients: An epidemiological study from a fever clinic in Wuhan, China. J Med Virol [Internet]. 2020; Available from: http://onlinelibrary.wiley.com/journal/10.1002/(ISSN)1096-9071](https://www.zotero.org/google-docs/?1sevUe)

4.	Haehner A, Draf J, Dräger S, de With K, Hummel T, de With K. Predictive Value of Sudden Olfactory Loss in the Diagnosis of COVID-19. ORL. 2020 Jul;82(4):175–80.

5.	Beltran-Corbellini A, Chico-Garcia JL, Martinez-Poles J, Rodriguez-Jorge F, Natera-Villalba E, Gomez-Corral J, et al. Acute-onset smell and taste disorders in the context of COVID-19: a pilot multicentre polymerase chain reaction based case-control study. Eur J Neurol. 2020;

6.	Renaud M, Leon A, Trau G, Fath L, Ciftci S, Bensimon Y, et al. Acute smell and taste loss in outpatients: all infected with SARS-CoV-2?. Rhinology. 2020;(347242).

7.	Gao S, Jiang F, Jin W, Shi Y, Yang L, Xia Y, et al. Risk factors influencing the prognosis of elderly patients infected with COVID-19: a clinical retrospective study in Wuhan, China. Aging. 2020;12(101508617).

8.	Abalo-Lojo JM, Pouso-Diz JM, Gonzalez F. Taste and Smell Dysfunction in COVID-19 Patients. Ann Otol Rhinol Laryngol. 2020;(407300):3489420932617.

9.	Adorni F, Prinelli F, Bianchi F, Giacomelli A, Pagani G, Bernacchia D, et al. Self-reported symptoms of SARS-CoV-2 infection in a non-hospitalized population: results from the large Italian web-based EPICOVID19 cross-sectional survey. JMIR Public Health Surveill. 2020;(101669345).

10.	Roland LT, Gurrola JG 2nd, Loftus PA, Cheung SW, Chang JL. Smell and taste symptom-based predictive model for COVID-19 diagnosis. Int Forum Allergy Rhinol. 2020;10(7):832–8.

11.	Aggarwal S, Garcia-Telles N, Aggarwal G, Lavie C, Lippi G, Henry BM. Clinical features, laboratory characteristics, and outcomes of patients hospitalized with coronavirus disease 2019 (COVID-19): Early report from the United States. Diagn Berl Ger. 2020;7(2):91–6.

[12.	Alsofayan Y.M., Althunayyan S.M., Khan A.A., Hakawi A.M., Assiri A.M. Clinical characteristics of COVID-19 in Saudi Arabia: A national retrospective study. J Infect Public Health [Internet]. 2020; Available from: http://www.elsevier.com/wps/find/journaldescription.cws_home/716388/descriptio](https://www.zotero.org/google-docs/?1sevUe)

13.	Manuel Romero-Sánchez C, Díaz-Maroto I, Fernández-Díaz E, Sánchez-Larsen Á, Layos-Romero A, García-García J, et al. Neurologic manifestations in hospitalized patients with COVID-19: The ALBACOVID registry. Neurology. 2020 Aug 25;95(8):e1060–70.

14.	Sakalli E, Temirbekov D, Bayri E, Alis EE, Erdurak SC, Bayraktaroglu M. Ear nose throat-related symptoms with a focus on loss of smell and/or taste in COVID-19 patients. Am J Otolaryngol. 2020;41(6):102622.

15.	Salmon Ceron D, Bartier S, Hautefort C, Nguyen Y, Nevoux J, Hamel A-L, et al. Self-reported loss of smell without nasal obstruction to identify COVID-19. The multicenter Coranosmia cohort study. J Infect. 2020 Oct;81(4):614–20.

16.	Chen Q, Zheng Z, Zhang C, Zhang X, Wu H, Wang J, et al. Clinical characteristics of 145 patients with corona virus disease 2019 (COVID-19) in Taizhou, Zhejiang, China. Infection. 2020 Aug;48(4):543–51.

17.	Chen T., Dai Z., Mo P., Li X., Ma Z., Song S., et al. Clinical characteristics and outcomes of older patients with coronavirus disease 2019 (COVID-19) in Wuhan, China (2019): a single-centered, retrospective study. J Gerontol A Biol Sci Med Sci. 2020;

18.	Liu X, Lin H, Jiang H, Li R, Zhong N, Su H, et al. Clinical characteristics of hospitalised patients with schizophrenia who were suspected to have coronavirus disease (COVID-19) in Hubei Province, China. Gen Psychiatry. 2020;33(2):e100222.

19.	Chiesa-Estomba C.M., Lechien J.R., Portillo-Mazal P., Martinez F., Cuauro-Sanchez J., Calvo-Henriquez C., et al. Olfactory and gustatory dysfunctions in COVID-19. First reports of Latin-American ethnic patients. Am J Otolaryngol - Head Neck Med Surg. 2020;41(5):102605.

20.	Lombardi A, Consonni D, Carugno M, Bozzi G, Mangioni D, Muscatello A, et al. Characteristics of 1573 healthcare workers who underwent nasopharyngeal swab testing for SARS-CoV-2 in Milan, Lombardy, Italy. Clin Microbiol Infect Off Publ Eur Soc Clin Microbiol Infect Dis. 2020;(dy9, 9516420).

21.	Oualha M, Bendavid M, Berteloot L, Corsia A, Lesage F, Vedrenne M, et al. Severe and fatal forms of COVID-19 in children. Arch Pediatr Organe Off Soc Francaise Pediatr. 2020;27(5):235–8.

22.	Paderno A, Schreiber A, Grammatica A, Raffetti E, Tomasoni M, Gualtieri T, et al. Smell and taste alterations in COVID-19: a cross-sectional analysis of different cohorts. Int Forum Allergy Rhinol. 2020;(101550261).

23.	Kim L, Garg S, O’Halloran A, Whitaker M, Pham H, Anderson EJ, et al. Risk Factors for Intensive Care Unit Admission and In-hospital Mortality among Hospitalized Adults Identified through the U.S. Coronavirus Disease 2019 (COVID-19)-Associated Hospitalization Surveillance Network (COVID-NET). Clin Infect Dis Off Publ Infect Dis Soc Am. 2020;

24.	Kim G-U, Kim M-J, Ra SH, Lee J, Bae S, Jung J, et al. Clinical characteristics of asymptomatic and symptomatic patients with mild COVID-19. Clin Microbiol Infect Off Publ Eur Soc Clin Microbiol Infect Dis. 2020;26(7):948.e1-948.e3.

25.	Kimball A, Hatfield KM, Arons M, James A, Taylor J, Spicer K, et al. Asymptomatic and Presymptomatic SARS-CoV-2 Infections in Residents of a Long-Term Care Skilled Nursing Facility - King County, Washington, March 2020. MMWR Morb Mortal Wkly Rep. 2020;69(13):377–81.

26.	Klopfenstein T, Kadiane-Oussou NJ, Toko L, Royer P-Y, Lepiller Q, Gendrin V, et al. Features of anosmia in COVID-19. Med Mal Infect. 2020;(0311416).

27.	Lovato A, Galletti C, Galletti B, de Filippis C. Clinical characteristics associated with persistent olfactory and taste alterations in COVID-19: A preliminary report on 121 patients. Am J Otolaryngol. 2020;41(5):102548.

28.	Chiesa-Estomba CM, Lechien JR, Radulesco T, Michel J, Sowerby LJ, Hopkins C, et al. Patterns of smell recovery in 751 patients affected by the COVID-19 outbreak. Eur J Neurol. 2020;

29.	Chougar L, Shor N, Weiss N, Galanaud D, Leclercq D, Mathon B, et al. Retrospective Observational Study of Brain Magnetic Resonance Imaging Findings in Patients with Acute SARS-CoV-2 Infection and Neurological Manifestations. Radiology. 2020;202422.

30.	Chung TW-H, Sridhar S, Zhang AJ, Chan K-H, Li H-L, Wong FK-C, et al. Olfactory Dysfunction in Coronavirus Disease 2019 Patients: Observational Cohort Study and Systematic Review. Open Forum Infect Dis. 2020;7(6):ofaa199.

31.	Gaur A., Meena S.K., Bairwa R., Meena D., Nanda R., Sharma S.R., et al. Clinico-radiological Presentation of COVID-19 Patients at a Tertiary Care Center at Bhilwara Rajasthan, India. J Assoc Physicians India. 2020;68(7):29–33.

32.	Altin F, Cingi C, Uzun T, Bal C. Olfactory and gustatory abnormalities in COVID-19 cases. Eur Arch Oto-Rhino-Laryngol Off J Eur Fed Oto-Rhino-Laryngol Soc EUFOS Affil Ger Soc Oto-Rhino-Laryngol - Head Neck Surg. 2020;(anb, 9002937).

33.	Annweiler C, Sacco G, Salles N, Aquino J-P, Gautier J, Berrut G, et al. National French survey of COVID-19 symptoms in people aged 70 and over. Clin Infect Dis Off Publ Infect Dis Soc Am. 2020;(a4j, 9203213).

34.	Gomez-Iglesias P, Porta-Etessam J, Montalvo T, Valls-Carbo A, Gajate V, Matias-Guiu JA, et al. An Online Observational Study of Patients With Olfactory and Gustory Alterations Secondary to SARS-CoV-2 Infection. Front Public Health. 2020;8(101616579):243.

35.	Clemency BM, Varughese R, Scheafer DK, Ludwig B, Welch JV, McCormack RF, et al. Symptom Criteria for COVID-19 Testing of Heath Care Workers. Acad Emerg Med Off J Soc Acad Emerg Med. 2020;27(6):469–74.

36.	Coelho DH, Kons ZA, Costanzo RM, Reiter ER. Subjective Changes in Smell and Taste During the COVID-19 Pandemic: A National Survey-Preliminary Results. Otolaryngol-Head Neck Surg. 2020 Aug;163(2):302–6.

37.	Gorzkowski V, Bevilacqua S, Charmillon A, Jankowski R, Gallet P, Rumeau C, et al. Evolution of olfactory disorders in COVID-19 patients. The Laryngoscope. 2020;(8607378).

38.	Han Y-N, Feng Z-W, Sun L-N, Ren X-X, Wang H, Xue Y-M, et al. A comparative-descriptive analysis of clinical characteristics in 2019-coronavirus-infected children and adults. J Med Virol. 2020;(i9n, 7705876).

39.	Lovell N, Maddocks M, Etkind SN, Taylor K, Carey I, Vora V, et al. Characteristics, Symptom Management, and Outcomes of 101 Patients With COVID-19 Referred for Hospital Palliative Care. J Pain Symptom Manage. 2020;60(1):e77–81.

40.	Lu L, Xiong W, Liu D, Liu J, Yang D, Li N, et al. New onset acute symptomatic seizure and risk factors in coronavirus disease 2019: A retrospective multicenter study. Epilepsia. 2020;61(6):e49–53.

41.	Sayin İ, Yaşar KK, Yazici ZM. Taste and Smell Impairment in COVID-19: An AAO-HNS Anosmia Reporting Tool-Based Comparative Study. Otolaryngol-Head Neck Surg. 2020 Sep;163(3):473–9.

42.	Schmithausen RM, Dohla M, Schobetaler H, Diegmann C, Schulte B, Richter E, et al. Characteristic Temporary Loss of Taste and Olfactory Senses in SARS-CoV-2-positive-Individuals with Mild Symptoms. Pathog Immun. 2020;5(1):117–20.

43.	Scullen T, Keen J, Mathkour M, Dumont AS, Kahn L. Coronavirus 2019 (COVID-19)-Associated Encephalopathies and Cerebrovascular Disease: The New Orleans Experience. World Neurosurg. 2020;(101528275).

44.	Belani P, Schefflein J, Kihira S, Rigney B, Delman BN, Mahmoudi K, et al. COVID-19 Is an Independent Risk Factor for Acute Ischemic Stroke. AJNR Am J Neuroradiol. 2020;(3).

45.	Bhandari S, Singh A, Sharma R, Rankawat G, Banerjee S, Gupta V, et al. Characteristics, Treatment Outcomes and Role of Hydroxychloroquine among 522 COVID-19 hospitalized patients in Jaipur City: An Epidemio-Clinical Study. J Assoc Physicians India. 2020;68(6):13–9.

46.	Hernandez-Fernandez F, Valencia HS, Barbella-Aponte RA, Collado-Jimenez R, Ayo-Martin O, Barrena C, et al. Cerebrovascular disease in patients with COVID-19: neuroimaging, histological and clinical description. Brain J Neurol. 2020;(0372537).

47.	Covino M, De Matteis G, Santoro M, Sabia L, Simeoni B, Candelli M, et al. Clinical characteristics and prognostic factors in COVID-19 patients aged >=80 years. Geriatr Gerontol Int. 2020;(101135738).

48.	Biadsee A, Biadsee A, Kassem F, Dagan O, Masarwa S, Ormianer Z. Olfactory and Oral Manifestations of COVID-19: Sex-Related Symptoms-A Potential Pathway to Early Diagnosis. Otolaryngol-Head Neck Surg. 2020 Oct;163(4):722–8.

49.	Hintschich CA, Wenzel JJ, Hummel T, Hankir MK, Kuhnel T, Vielsmeier V, et al. Psychophysical tests reveal impaired olfaction but preserved gustation in COVID-19 patients. Int Forum Allergy Rhinol. 2020;(101550261).

50.	Sierpinski R, Pinkas J, Jankowski M, Zgliczynski WS, Wierzba W, Gujski M, et al. Sex differences in the frequency of gastrointestinal symptoms and olfactory or taste disorders in 1942 nonhospitalized patients with coronavirus disease 2019 (COVID-19). Pol Arch Intern Med. 2020;130(6):501–5.

51.	Paderno A, Mattavelli D, Rampinelli V, Grammatica A, Raffetti E, Tomasoni M, et al. Olfactory and Gustatory Outcomes in COVID-19: A Prospective Evaluation in Nonhospitalized Subjects. Otolaryngol--Head Neck Surg Off J Am Acad Otolaryngol-Head Neck Surg. 2020;(7909794, 8508176, on7, on8):194599820939538.

52.	Hornuss D, Lange B, Schroter N, Rieg S, Kern WV, Wagner D. Anosmia in COVID-19 patients. Clin Microbiol Infect Off Publ Eur Soc Clin Microbiol Infect Dis. 2020;(dy9, 9516420).

53.	Boscolo-Rizzo P, Borsetto D, Fabbris C, Spinato G, Frezza D, Menegaldo A, et al. Evolution of Altered Sense of Smell or Taste in Patients With Mildly Symptomatic COVID-19. JAMA Otolaryngol-- Head Neck Surg. 2020;(101589542).

54.	Huang C., Wang Y., Li X., Ren L., Zhao J., Hu Y., et al. Clinical features of patients infected with 2019 novel coronavirus in Wuhan, China. The Lancet. 2020;395(10223):497–506.

55.	Huang D, Wang T, Chen Z, Yang H, Yao R, Liang Z. A novel risk score to predict diagnosis with coronavirus disease 2019 (COVID-19) in suspected patients: A retrospective, multicenter, and observational study. J Med Virol. 2020;(i9n, 7705876).

56.	Parra A, Juanes A, Losada CP, Alvarez-Sesmero S, Santana VD, Marti I, et al. Psychotic symptoms in COVID-19 patients. A retrospective descriptive study. Psychiatry Res. 2020;291:113254.

57.	Luers JC, Rokohl AC, Loreck N, Wawer Matos PA, Augustin M, Dewald F, et al. Olfactory and Gustatory Dysfunction in Coronavirus Disease 19 (COVID-19). Clin Infect Dis Off Publ Infect Dis Soc Am. 2020;

58.	Somekh I, Yakub Hanna H, Heller E, Bibi H, Somekh E. AGE-DEPENDENT SENSORY IMPAIRMENT IN COVID-19 INFECTION AND ITS CORRELATION WITH ACE2 EXPRESSION. Pediatr Infect Dis J. 2020;

59.	Song S, Wu F, Liu Y, Jiang H, Xiong F, Guo X, et al. Correlation Between Chest CT Findings and Clinical Features of 211 COVID-19 Suspected Patients in Wuhan, China. Open Forum Infect Dis. 2020;7(6):ofaa171.

60.	Immovilli P, Terracciano C, Zaino D, Marchesi E, Morelli N, Terlizzi E, et al. Stroke in COVID-19 patients-A case series from Italy. Int J Stroke Off J Int Stroke Soc. 2020;(101274068):1747493020938294.

61.	Pastor J, Vega-Zelaya L, Martin Abad E. Specific EEG Encephalopathy Pattern in SARS-CoV-2 Patients. J Clin Med. 2020;9(5).

62.	Izquierdo-Dominguez A, Rojas-Lechuga MJ, Chiesa-Estomba C, Calvo-Henriquez C, Ninchritz-Becerra E, Soriano-Reixach M, et al. Smell and taste dysfunctions in COVID-19 are associated with younger age in ambulatory settings - a multicenter cross-sectional study. J Investig Allergol Clin Immunol. 2020;(9107858):0.

63.	Ma S., Lai X., Chen Z., Tu S., Qin K. Clinical characteristics of critically ill patients co-infected with SARS-CoV-2 and the influenza virus in Wuhan, China. Int J Infect Dis. 2020;96:683–7.

64.	Ma Y-F, Li W, Deng H-B, Wang L, Wang Y, Wang P-H, et al. Prevalence of depression and its association with quality of life in clinically stable patients with COVID-19. J Affect Disord. 2020;275:145–8.

65.	Mao B., Liu Y., Chai Y.-H., Jin X.-Y., Lu H.-W., Yang J.-W., et al. Assessing risk factors for SARS-CoV-2 infection in patients presenting with symptoms in Shanghai, China: a multicentre, observational cohort study. Lancet Digit Health. 2020;2(6):e323–30.

66.	Marinho PM, Marcos AAA, Romano AC, Nascimento H, Belfort R, Belfort RJ. Retinal findings in patients with COVID-19. Lancet. 2020;395(10237):1610–1610.

67.	Mariotto S, Savoldi A, Donadello K, Zanzoni S, Bozzetti S, Carta S, et al. Nervous system: subclinical target of SARS-CoV-2 infection. J Neurol Neurosurg Psychiatry. 2020;(2985191r, jbb).

68.	Mathian A., Mahevas M., Rohmer J., Roumier M., Cohen-Aubart F., Amador-Borrero B., et al. Clinical course of coronavirus disease 2019 (COVID-19) in a series of 17 patients with systemic lupus erythematosus under long-Term treatment with hydroxychloroquine. Ann Rheum Dis. 2020;79(6):837–9.

69.	Mcloughlin BC, Miles A, Webb TE, Knopp P, Eyres C, Fabbri A, et al. Functional and cognitive outcomes after COVID-19 delirium. Eur Geriatr Med. 2020;(101533694).

70.	Patel A, Charani E, Ariyanayagam D, Abdulaal A, Denny SJ, Mughal N, et al. New-onset anosmia and ageusia in adult patients diagnosed with SARS-CoV-2 infection. Clin Microbiol Infect Off Publ Eur Soc Clin Microbiol Infect Dis. 2020;(dy9, 9516420).

71.	Mehrpour M, Shuaib A, Farahani M, Hatamabadi HR, Fatehi Z, Ghaffari M, et al. Coronavirus disease 2019 and stroke in Iran: a case series and effects on stroke admissions. Int J Stroke Off J Int Stroke Soc. 2020;(101274068):1747493020937397.

72.	Mei X, Zhang Y, Zhu H, Ling Y, Zou Y, Zhang Z, et al. Observations about Symptomatic and Asymptomatic infections of 494 patients with COVID-19 in Shanghai,China. Am J Infect Control. 2020;(4).

73.	Brandstetter S, Roth S, Harner S, Buntrock-Dopke H, Toncheva AA, Borchers N, et al. Symptoms and immunoglobulin development in hospital staff exposed to a SARS-CoV-2 outbreak. Pediatr Allergy Immunol Off Publ Eur Soc Pediatr Allergy Immunol. 2020;(bu6, 9106718).

74.	Jackel M, Bemtgen X, Wengenmayer T, Bode C, Biever PM, Staudacher DL. Is delirium a specific complication of viral acute respiratory distress syndrome?. Crit Care Lond Engl. 2020;24(1):401.

75.	Jalessi M, Barati M, Rohani M, Amini E, Ourang A, Azad Z, et al. Frequency and outcome of olfactory impairment and sinonasal involvement in hospitalized patients with COVID-19. Neurol Sci Off J Ital Neurol Soc Ital Soc Clin Neurophysiol. 2020;(drb, 100959175).

76.	Joffily L, Ungierowicz A, David AG, Melo B, Brito CLT, Mello L, et al. The close relationship between sudden loss of smell and COVID-19. Braz J Otorhinolaryngol. 2020;(101207337).

77.	Kai Chua A.J., Yun Chan E.C., Loh J., Charn T.C. Acute olfactory loss is specific for Covid-19 at the Emergency Department. Ann Emerg Med. 2020;

78.	Kandemirli SG, Dogan L, Sarikaya ZT, Kara S, Akinci C, Kaya D, et al. Brain MRI Findings in Patients in the Intensive Care Unit with COVID-19 Infection. Radiology. 2020;201697.

79.	Karadaş Ö, Öztürk B, Sonkaya AR. A prospective clinical study of detailed neurological manifestations in patients with COVID-19. Neurol Sci. 2020 Aug;41(8):1991–5.

80.	Carignan A, Valiquette L, Grenier C, Musonera JB, Nkengurutse D, Marcil-Heguy A, et al. Anosmia and dysgeusia associated with SARS-CoV-2 infection: an age-matched case-control study. CMAJ Can Med Assoc J J Assoc Medicale Can. 2020;192(26):E702–7.

81.	Meini S, Suardi LR, Busoni M, Roberts AT, Fortini A. Olfactory and gustatory dysfunctions in 100 patients hospitalized for COVID-19: sex differences and recovery time in real-life. Eur Arch Oto-Rhino-Laryngol Off J Eur Fed Oto-Rhino-Laryngol Soc EUFOS Affil Ger Soc Oto-Rhino-Laryngol - Head Neck Surg. 2020;(anb, 9002937).

82.	Menni C, Valdes AM, Freidin MB, Sudre CH, Nguyen LH, Drew DA, et al. Real-time tracking of self-reported symptoms to predict potential COVID-19. Nat Med. 2020;26(7):1037–40.

83.	Korkmaz MF, Ture E, Dorum BA, Kilic ZB. The Epidemiological and Clinical Characteristics of 81 Children with COVID-19 in a Pandemic Hospital in Turkey: an Observational Cohort Study. J Korean Med Sci. 2020;35(25):e236.

84.	Mercante G, Ferreli F, De Virgilio A, Gaino F, Di Bari M, Colombo G, et al. Prevalence of Taste and Smell Dysfunction in Coronavirus Disease 2019. JAMA Otolaryngol-- Head Neck Surg. 2020;(101589542).

85.	Speth MM, Singer-Cornelius T, Oberle M, Gengler I, Brockmeier SJ, Sedaghat AR. Mood, anxiety and olfactory dysfunction in COVID-19: evidence of central nervous system involvement?. The Laryngoscope. 2020;(8607378).

86.	Speth MM, Singer-Cornelius T, Oberle M, Gengler I, Brockmeier SJ, Sedaghat AR. Olfactory Dysfunction and Sinonasal Symptomatology in COVID-19: Prevalence, Severity, Timing, and Associated Characteristics. Otolaryngol-Head Neck Surg. 2020 Jul;163(1):114–20.

87.	Kremer S, Lersy F, de Seze J, Ferre J-C, Maamar A, Carsin-Nicol B, et al. Brain MRI Findings in Severe COVID-19: A Retrospective Observational Study. Radiology. 2020;202222.

88.	Spoldi C, Castellani L, Pipolo C, Maccari A, Lozza P, Scotti A, et al. Isolated olfactory cleft involvement in SARS-CoV-2 infection: prevalence and clinical correlates. Eur Arch Oto-Rhino-Laryngol Off J Eur Fed Oto-Rhino-Laryngol Soc EUFOS Affil Ger Soc Oto-Rhino-Laryngol - Head Neck Surg. 2020;(anb, 9002937).

[89.	Sun L., Shen L., Fan J., Gu F., Hu M., An Y., et al. Clinical Features of Patients with Coronavirus Disease 2019 (COVID-19) from a Designated Hospital in Beijing, China. J Med Virol [Internet]. 2020; Available from: http://onlinelibrary.wiley.com/journal/10.1002/(ISSN)1096-9071](https://www.zotero.org/google-docs/?1sevUe)

90.	Sweid A, Hammoud B, Bekelis K, Missios S, Tjoumakaris SI, Gooch MR, et al. Cerebral ischemic and hemorrhagic complications of coronavirus disease 2019. Int J Stroke Off J Int Stroke Soc. 2020;(101274068):1747493020937189.

91.	Chary E, Carsuzaa F, Trijolet J-P, Capitaine A-L, Roncato-Saberan M, Fouet K, et al. Prevalence and Recovery From Olfactory and Gustatory Dysfunctions in Covid-19 Infection: A Prospective Multicenter Study. Am J Rhinol Allergy. 2020;(101490775):1945892420930954.

92.	Lapostolle F, Schneider E, Vianu I, Dollet G, Roche B, Berdah J, et al. Clinical features of 1487 COVID-19 patients with outpatient management in the Greater Paris: the COVID-call study. Intern Emerg Med. 2020;(101263418).

93.	Lechien JR, Cabaraux P, Chiesa-Estomba CM, Khalife M, Plzak J, Hans S, et al. Psychophysical Olfactory Tests and Detection of COVID-19 in Patients With Sudden Onset Olfactory Dysfunction: A Prospective Study. Ear Nose Throat J. 2020;(edf, 7701817):145561320929169.

94.	Lechien JR, Michel J, Radulesco T, Chiesa-Estomba CM, Vaira LA, De Riu G, et al. Clinical and Radiological Evaluations of COVID-19 Patients with Anosmia: Preliminary Report. The Laryngoscope. 2020;(8607378, l1w).

95.	Lechien JR, Chiesa-Estomba CM, Cabaraux P, Mat Q, Huet K, Harmegnies B, et al. Features of Mild-to-Moderate COVID-19 Patients With Dysphonia. J Voice Off J Voice Found. 2020;

96.	Lechien JR, Chiesa‐Estomba CM, Place S, Van Laethem Y, Cabaraux P, Mat Q, et al. Clinical and epidemiological characteristics of 1420 European patients with mild-to-moderate coronavirus disease 2019. J Intern Med. 2020 Sep;288(3):335–44.

97.	Lechien JR, Chiesa-Estomba CM, Hans S, Barillari MR, Jouffe L, Saussez S. Loss of Smell and Taste in 2013 European Patients With Mild to Moderate COVID-19. Ann Intern Med. 2020;(0372351).

98.	Lechien JR, Cabaraux P, Chiesa-Estomba CM, Khalife M, Hans S, Calvo-Henriquez C, et al. Objective olfactory evaluation of self-reported loss of smell in a case series of 86 COVID-19 patients. Head Neck. 2020;42(7):1583–90.

99.	Khan M, Ibrahim RH, Siddiqi SA, Kerolos Y, Al-Kaylani MM, AlRukn SA, et al. COVID-19 and acute ischemic stroke - A case series from Dubai, UAE. Int J Stroke Off J Int Stroke Soc. 2020;(101274068):1747493020938285.

100.	Kihira S, Schefflein J, Chung M, Mahmoudi K, Rigney B, Delman BN, et al. Incidental COVID-19 related lung apical findings on stroke CTA during the COVID-19 pandemic. J Neurointerventional Surg. 2020;12(7):669–72.

101.	Lee DJ, Lockwood J, Das P, Wang R, Grinspun E, Lee JM. Self-reported anosmia and dysgeusia as key symptoms of coronavirus disease 2019. CJEM Can J Emerg Med. 2020 Sep;22(5):595–602.

102.	Lee SA, Jobe MC, Mathis AA. Mental health characteristics associated with dysfunctional coronavirus anxiety. Psychol Med. 2020;1–2.

103.	Lee JY, Kim HA, Huh K, Hyun M, Rhee JY, Jang S, et al. Risk Factors for Mortality and Respiratory Support in Elderly Patients Hospitalized with COVID-19 in Korea. J Korean Med Sci. 2020;35(23):e223.

104.	Lee Y, Min P, Lee S, Kim SW. Prevalence and Duration of Acute Loss of Smell or Taste in COVID-19 Patients. J Korean Med Sci. 2020;35(18):e174.

105.	Li Y, Li M, Wang M, Zhou Y, Chang J, Xian Y, et al. Acute cerebrovascular disease following COVID-19: a single center, retrospective, observational study. Stroke Vasc Neurol. 2020;(101689996).

106.	Merkler AE, Parikh NS, Mir S, Gupta A, Kamel H, Lin E, et al. Risk of Ischemic Stroke in Patients With Coronavirus Disease 2019 (COVID-19) vs Patients With Influenza. JAMA Neurol. 2020;(101589536).

107.	Moein ST, Hashemian SM, Mansourafshar B, Khorram-Tousi A, Tabarsi P, Doty RL. Smell dysfunction: a biomarker for COVID-19. Int Forum Allergy Rhinol. 2020;(101550261).

108.	Moro E, Priori A, Beghi E, Helbok R, Campiglio L, Bassetti CL, et al. The international EAN survey on neurological symptoms in patients with COVID-19 infection. Eur J Neurol. 2020;(daf, 9506311).

109.	Nalleballe K, Reddy Onteddu S, Sharma R, Dandu V, Brown A, Jasti M, et al. Spectrum of neuropsychiatric manifestations in COVID-19. Brain Behav Immun. 2020;

110.	Li Z, Zheng C, Duan C, Zhang Y, Li Q, Dou Z, et al. Rehabilitation needs of the first cohort of post-acute COVID-19 patients in Hubei, China. Eur J Phys Rehabil Med. 2020;56(3):339–44.

111.	Li L, Yang L, Gui S, Pan F, Ye T, Liang B, et al. Association of clinical and radiographic findings with the outcomes of 93 patients with COVID-19 in Wuhan, China. Theranostics. 2020;10(14):6113–21.

112.	Li K, Wu J, Wu F, Guo D, Chen L, Fang Z, et al. The Clinical and Chest CT Features Associated With Severe and Critical COVID-19 Pneumonia. Invest Radiol. 2020;55(6):327–31.

113.	Paterson RW, Brown RL, Benjamin L, Nortley R, Wiethoff S, Bharucha T, et al. The emerging spectrum of COVID-19 neurology: clinical, radiological and laboratory findings. Brain J Neurol. 2020;(0372537).

114.	D’Ascanio L, Pandolfini M, Cingolani C, Latini G, Gradoni P, Capalbo M, et al. Olfactory Dysfunction in COVID-19 Patients: Prevalence and Prognosis for Recovering Sense of Smell. Otolaryngol--Head Neck Surg Off J Am Acad Otolaryngol-Head Neck Surg. 2020;(7909794, 8508176, on7, on8):194599820943530.

115.	Dawson P, Rabold EM, Laws RL, Conners EE, Gharpure R, Yin S, et al. Loss of Taste and Smell as Distinguishing Symptoms of COVID-19. Clin Infect Dis Off Publ Infect Dis Soc Am. 2020;(a4j, 9203213).

116.	Dell’Era V, Farri F, Garzaro G, Gatto M, Aluffi Valletti P, Garzaro M. Smell and taste disorders during COVID-19 outbreak: Cross-sectional study on 355 patients. Head Neck. 2020;42(7):1591–6.

117.	Tabata S, Imai K, Kawano S, Ikeda M, Kodama T, Miyoshi K, et al. Clinical characteristics of COVID-19 in 104 people with SARS-CoV-2 infection on the Diamond Princess cruise ship: a retrospective analysis. Lancet Infect Dis. 2020;(101130150).

[118.	Tian J., Yuan X., Xiao J., Zhong Q., Yang C., Liu B., et al. Clinical characteristics and risk factors associated with COVID-19 disease severity in patients with cancer in Wuhan, China: a multicentre, retrospective, cohort study. Lancet Oncol [Internet]. 2020; Available from: http://www.journals.elsevier.com/the-lancet-oncology/](https://www.zotero.org/google-docs/?1sevUe)

119.	Tostmann A, Bradley J, Bousema T, Yiek W-K, Holwerda M, Bleeker-Rovers C, et al. Strong associations and moderate predictive value of early symptoms for SARS-CoV-2 test positivity among healthcare workers, the Netherlands, March 2020. Euro Surveill Bull Eur Sur Mal Transm Eur Commun Dis Bull. 2020;25(16).

120.	Du N, Chen H, Zhang Q, Che L, Lou L, Li X, et al. A case series describing the epidemiology and clinical characteristics of COVID-19 infection in Jilin Province. Virulence. 2020;11(1):482–5.

121.	Du W., Yu J., Wang H., Zhang X., Zhang S., Li Q., et al. Clinical characteristics of COVID-19 in children compared with adults in Shandong Province, China. Infection. 2020;48(3):445–52.

122.	Trubiano JA, Vogrin S, Kwong JC, Holmes NE. Alterations in smell or taste - Classic COVID-19?. Clin Infect Dis Off Publ Infect Dis Soc Am. 2020;(a4j, 9203213).

123.	Guan W-J, Ni Z-Y, Hu Y, Liang W-H, Ou C-Q, He J-X, et al. Clinical Characteristics of Coronavirus Disease 2019 in China. Zhong NS LL Zeng G, Gao GF, Yuen KY, editor. N Engl J Med. 2020;382(18):1708–20.

124.	Escalard S, Maïer B, Redjem H, Delvoye F, Hébert S, Smajda S, et al. Treatment of Acute Ischemic Stroke due to Large Vessel Occlusion With COVID-19: Experience From Paris. Stroke 00392499. 2020 Aug;51(8):2540–3.

125.	Farahani R.H., Gholami M., Hazrati E., Rouzbahani N.H., Hejripour Z., Soleiman-Meigooni S., et al. Clinical features of icu admitted and intubated novel corona virus-infected patients in iran. Arch Clin Infect Dis. 2020;15(2):e103295.

126.	Freni F, Meduri A, Gazia F, Nicastro V, Galletti C, Aragona P, et al. Symptomatology in head and neck district in coronavirus disease (COVID-19): A possible neuroinvasive action of SARS-CoV-2. Am J Otolaryngol. 2020;41(5):102612.

127.	Galanopoulou AS, Ferastraoaru V, Correa DJ, Cherian K, Duberstein S, Gursky J, et al. EEG findings in acutely ill patients investigated for SARS-CoV-2/COVID-19: A small case series preliminary report. Epilepsia Open. 2020;5(2):314–24.

128.	Tudrej B, Sebo P, Lourdaux J, Cuzin C, Floquet M, Haller DM, et al. Self-Reported Loss of Smell and Taste in SARS-CoV-2 Patients: Primary Care Data to Guide Future Early Detection Strategies. J Gen Intern Med. 2020;(8605834).

129.	Vacchiano V, Riguzzi P, Volpi L, Tappatà M, Avoni P, Rizzo G, et al. Early neurological manifestations of hospitalized COVID-19 patients. Neurol Sci. 2020 Aug;41(8):2029–31.

130.	Vaira LA, Hopkins C, Salzano G, Petrocelli M, Melis A, Cucurullo M, et al. Olfactory and gustatory function impairment in COVID-19 patients: Italian objective multicenter-study. Head Neck. 2020;42(7):1560–9.

131.	Vaira LA, Deiana G, Fois AG, Pirina P, Madeddu G, De Vito A, et al. Objective evaluation of anosmia and ageusia in COVID-19 patients: Single-center experience on 72 cases. Head Neck. 2020;42(6):1252–8.

132.	Boscolo-Rizzo P, Borsetto D, Spinato G, Fabbris C, Menegaldo A, Gaudioso P, et al. New onset of loss of smell or taste in household contacts of home-isolated SARS-CoV-2-positive subjects. Eur Arch Oto-Rhino-Laryngol Off J Eur Fed Oto-Rhino-Laryngol Soc EUFOS Affil Ger Soc Oto-Rhino-Laryngol - Head Neck Surg. 2020;(anb, 9002937).

133.	Burke RM, Killerby ME, Newton S, Ashworth CE, Berns AL, Brennan S, et al. Symptom Profiles of a Convenience Sample of Patients with COVID-19 - United States, January-April 2020. MMWR Morb Mortal Wkly Rep. 2020;69(28):904–8.

134.	Cecchetti G, Vabanesi M, Chieffo R, Fanelli G, Minicucci F, Agosta F, et al. Cerebral involvement in COVID-19 is associated with metabolic and coagulation derangements: an EEG study. J Neurol. 2020;

135.	Chen J, Zhang Z-Z, Chen Y-K, Long Q-X, Tian W-G, Deng H-J, et al. The clinical and immunological features of pediatric COVID-19 patients in China. Genes Dis. 2020;(101635967).

136.	Chen T, Wu D, Chen H, Yan W, Yang D, Chen G, et al. Clinical characteristics of 113 deceased patients with coronavirus disease 2019: retrospective study. BMJ. 2020;368(8900488):m1091.

137.	Denis F, Galmiche S, Dinh A, Fontanet A, Scherpereel A, Benezit F, et al. Epidemiological Observations on the Association Between Anosmia and COVID-19 Infection: Analysis of Data From a Self-Assessment Web Application. J Med Internet Res. 2020;22(6):e19855.

138.	Duan X, Guo X, Qiang J. A retrospective study of the initial 25 COVID-19 patients in Luoyang, China. Jpn J Radiol. 2020;38(7):683–90.

139.	Fraisse M., Logre E., Pajot O., Mentec H., Plantefeve G., Contou D. Thrombotic and hemorrhagic events in critically ill COVID-19 patients: A French monocenter retrospective study. Crit Care. 2020;24:275.

140.	Gaborieau L, Delestrain C, Bensaid P, Vizeneux A, Blanc P, Garraffo A, et al. Epidemiology and Clinical Presentation of Children Hospitalized with SARS-CoV-2 Infection in Suburbs of Paris. J Clin Med. 2020;9(7).

141.	Garazzino S., Montagnani C., Dona D., Meini A., Felici E., Vergine G., et al. Multicentre Italian study of SARS-CoV-2 infection in children and adolescents, preliminary data as at 10 April 2020. Eurosurveillance. 2020;25(18):1–4.

142.	Guo Q, Zheng Y, Shi J, Wang J, Li G, Li C, et al. Immediate psychological distress in quarantined patients with COVID-19 and its association with peripheral inflammation: A mixed-method study. Brain Behav Immun. 2020;

143.	Kaye R, Chang CWD, Kazahaya K, Brereton J, Denneny III JC, Denneny JC 3rd. COVID-19 Anosmia Reporting Tool: Initial Findings. Otolaryngol-Head Neck Surg. 2020 Jul;163(1):132–4.

144.	Li J, Chen Z, Nie Y, Ma Y, Guo Q, Dai X. Identification of Symptoms Prognostic of COVID-19 Severity: Multivariate Data Analysis of a Case Series in Henan Province. J Med Internet Res. 2020;22(6):e19636.

145.	Liang Y, Xu J, Chu M, Mai J, Lai N, Tang W, et al. Neurosensory dysfunction: a diagnostic marker of early COVID-19. Int J Infect Dis IJID Off Publ Int Soc Infect Dis. 2020;(c3r, 9610933).

146.	Liu J-Y, Chen T-J, Hwang S-J. Analysis of Imported Cases of COVID-19 in Taiwan: A Nationwide Study. Int J Environ Res Public Health. 2020;17(9).

147.	Luo Y, Wu J, Lu J, Xu X, Long W, Yan G, et al. Investigation of COVID-19-related symptoms based on factor analysis. Ann Palliat Med. 2020;(101585484).

148.	Mao L, Jin H, Wang M, Hu Y, Chen S, He Q, et al. Neurologic Manifestations of Hospitalized Patients With Coronavirus Disease 2019 in Wuhan, China. JAMA Neurol. 2020;(101589536).

149.	Myrstad M, Ihle-Hansen H, Tveita AA, Andersen EL, Nygard S, Tveit A, et al. National Early Warning Score 2 (NEWS2) on admission predicts severe disease and in-hospital mortality from Covid-19 - a prospective cohort study. Scand J Trauma Resusc Emerg Med. 2020;28(1):66.

150.	Naeini AS, Karimi-Galougahi M, Raad N, Ghorbani J, Taraghi A, Haseli S, et al. Paranasal sinuses computed tomography findings in anosmia of COVID-19. Am J Otolaryngol. 2020;41(6):102636.

151.	Nguyen HC, Nguyen MH, Do BN, Tran CQ, Nguyen TTP, Pham KM, et al. People with Suspected COVID-19 Symptoms Were More Likely Depressed and Had Lower Health-Related Quality of Life: The Potential Benefit of Health Literacy. J Clin Med. 2020;9(4).

152.	Pati S., Toth E., Chaitanya G. Quantitative EEG markers to prognosticate critically ill patients with COVID-19: A retrospective cohort study. Clin Neurophysiol. 2020;131(8):1824–6.

153.	Paz C, Mascialino G, Adana-Diaz L, Rodriguez-Lorenzana A, Simbana-Rivera K, Gomez-Barreno L, et al. Anxiety and depression in patients with confirmed and suspected COVID-19 in Ecuador. Psychiatry Clin Neurosci. 2020;

154.	Petrescu A-M, Taussig D, Bouilleret V. Electroencephalogram (EEG) in COVID-19: A systematic retrospective study. Neurophysiol Clin Clin Neurophysiol. 2020;(nec, 8804532).

155.	Petrocelli M, Ruggiero F, Baietti AM, Pandolfi P, Salzano G, Salzano FA, et al. Remote psychophysical evaluation of olfactory and gustatory functions in early-stage coronavirus disease 2019 patients: the Bologna experience of 300 cases. J Laryngol Otol. 2020;(8706896, iwn):1–12.

156.	Peyrony O, Marbeuf-Gueye C, Truong V, Giroud M, Riviere C, Khenissi K, et al. Accuracy of Emergency Department Clinical Findings for Diagnosis of Coronavirus Disease 2019. Ann Emerg Med. 2020;(8002646).

157.	Pinna P, Grewal P, Hall JP, Tavarez T, Dafer RM, Garg R, et al. Neurological manifestations and COVID-19: Experiences from a tertiary care center at the Frontline. J Neurol Sci. 2020;415:116969.

158.	Pons-Escoda A, Naval-Baudin P, Majos C, Camins A, Cardona P, Cos M, et al. Neurologic Involvement in COVID-19: Cause or Coincidence? A Neuroimaging Perspective. AJNR Am J Neuroradiol. 2020;(3).

159.	Porta-Etessam J, Matias-Guiu JA, Gonzalez-Garcia N, Gomez Iglesias P, Santos-Bueso E, Arriola-Villalobos P, et al. Spectrum of Headaches Associated With SARS-CoV-2 Infection: Study of Healthcare Professionals. Headache. 2020;(2985091).

160.	Qin C, Zhou L, Hu Z, Yang S, Zhang S, Chen M, et al. Clinical Characteristics and Outcomes of COVID-19 Patients With a History of Stroke in Wuhan, China. Stroke. 2020;51(7):2219–23.

161.	Qiu C, Cui C, Hautefort C, Haehner A, Zhao J, Yao Q, et al. Olfactory and Gustatory Dysfunction as an Early Identifier of COVID-19 in Adults and Children: An International Multicenter Study. Otolaryngol--Head Neck Surg Off J Am Acad Otolaryngol-Head Neck Surg. 2020;(7909794, 8508176, on7, on8):194599820934376.

162.	Radmanesh A, Derman A, Lui YW, Raz E, Loh JP, Hagiwara M, et al. COVID-19 -associated Diffuse Leukoencephalopathy and Microhemorrhages. Radiology. 2020;(qsh, 0401260):202040.

163.	Reddy ST, Garg T, Shah C, Nascimento FA, Imran R, Kan P, et al. Cerebrovascular Disease in Patients with COVID-19: A Review of the Literature and Case Series. Case Rep Neurol. 2020;12(2):199–209.

164.	Shi M, Chen L, Yang Y, Zhang J, Xu J, Xu G, et al. Analysis of clinical features and outcomes of 161 patients with severe and critical COVID-19: A multicenter descriptive study. J Clin Lab Anal. 2020;e23415.

165.	Soltani J, Sedighi I, Shalchi Z, Sami G, Moradveisi B, Nahidi S. Pediatric coronavirus disease 2019 (COVID-19): An insight from west of Iran. North Clin Istanb. 2020;7(3):284–91.

166.	Spinato G, Fabbris C, Polesel J, Cazzador D, Borsetto D, Hopkins C, et al. Alterations in Smell or Taste in Mildly Symptomatic Outpatients With SARS-CoV-2 Infection. JAMA. 2020;(7501160).

167.	Su L, Ma X, Yu H, Zhang Z, Bian P, Han Y, et al. The different clinical characteristics of corona virus disease cases between children and their families in China - the character of children with COVID-19. Emerg Microbes Infect. 2020;9(1):707–13.

168.	Sun H., Ning R., Tao Y., Yu C., Deng X., Zhao C., et al. Risk Factors for Mortality in 244 Older Adults With COVID-19 in Wuhan, China: A Retrospective Study. J Am Geriatr Soc. 2020;68(6):E19–23.

169.	Tenforde MW, Rose EB, Lindsell CJ, Shapiro NI, Files DC, Gibbs KW, et al. Characteristics of Adult Outpatients and Inpatients with COVID-19 - 11 Academic Medical Centers, United States, March-May 2020. MMWR Morb Mortal Wkly Rep. 2020 Jul 3;69(26):841–6.

170.	Vaira LA, Salzano G, Petrocelli M, Deiana G, Salzano FA, De Riu G. Validation of a self-administered olfactory and gustatory test for the remotely evaluation of COVID-19 patients in home quarantine. Head Neck. 2020;42(7):1570–6.

171.	Varatharaj A, Thomas N, Ellul MA, Davies NWS, Pollak TA, Tenorio EL, et al. Neurological and neuropsychiatric complications of COVID-19 in 153 patients: a UK-wide surveillance study. Lancet Psychiatry. 2020;(101638123).

172.	Vargas-Gandica J, Winter D, Schnippe R, Rodriguez-Morales AG, Mondragon J, Escalera-Antezana JP, et al. Ageusia and anosmia, a common sign of COVID-19? A case series from four countries. J Neurovirol. 2020;

173.	Vespignani H, Colas D, Lavin BS, Soufflet C, Maillard L, Pourcher V, et al. Report on Electroencephalographic Findings in Critically Ill Patients with COVID-19. Ann Neurol. 2020 Sep;88(3):626–30.

174.	Wang X, Xu H, Jiang H, Wang L, Lu C, Wei X, et al. The Clinical Features and Outcomes of Discharged Coronavirus Disease 2019 Patients:A Prospective Cohort Study. QJM Mon J Assoc Physicians. 2020;(9438285, B4V).

175.	Wu J, Chen X, Yao S, Liu R. Anxiety persists after recovery from acquired COVID-19 in anaesthesiologists. J Clin Anesth. 2020;67:109984.

176.	Xiao M, Zhang Y, Zhang S, Qin X, Xia P, Cao W, et al. Brief Report: Anti-phospholipid antibodies in critically ill patients with Coronavirus Disease 2019 (COVID-19). Arthritis Rheumatol Hoboken NJ. 2020;(101623795).

177.	Xie S, Zhang G, Yu H, Wang J, Wang S, Tang G, et al. The epidemiologic and clinical features of suspected and confirmed cases of imported 2019 novel coronavirus pneumonia in north Shanghai, China. Ann Transl Med. 2020;8(10):637.

178.	Xiong W, Mu J, Guo J, Lu L, Liu D, Luo J, et al. New onset neurologic events in people with COVID-19 infection in three regions in China. Neurology. 2020;(401060).

179.	Yan CH, Faraji F, Prajapati DP, Boone CE, DeConde AS. Association of chemosensory dysfunction and COVID-19 in patients presenting with influenza-like symptoms. Int Forum Allergy Rhinol. 2020;10(7):806–13.

180.	Yan CH, Prajapati DP, Ritter ML, DeConde AS. Persistent Smell Loss Following Undetectable SARS-CoV-2. Otolaryngol--Head Neck Surg Off J Am Acad Otolaryngol-Head Neck Surg. 2020;(7909794, 8508176, on7, on8):194599820934769.

181.	Yang Q, Xie L, Zhang W, Zhao L, Wu H, Jiang J, et al. Analysis of the clinical characteristics, drug treatments and prognoses of 136 patients with coronavirus disease 2019. J Clin Pharm Ther. 2020;45(4):609–16.

182.	Yin Z, Kang Z, Yang D, Ding S, Luo H, Xiao E. A Comparison of Clinical and Chest CT Findings in Patients With Influenza A (H1N1) Virus Infection and Coronavirus Disease (COVID-19). AJR Am J Roentgenol. 2020;(3):1–7.

[183.	Yu J., He W., Chen L., Yuan G., Dong F., Chen W., et al. Risk factors for death in 1859 subjects with COVID-19. Leukemia [Internet]. 2020; Available from: http://www.nature.com/leu/index.html](https://www.zotero.org/google-docs/?1sevUe)

184.	Yu T, Tian C, Chu S, Zhou H, Zhang Z, Luo S, et al. COVID-19 patients benefit from early antiviral treatment: a comparative, retrospective study. J Med Virol. 2020;(i9n, 7705876).

185.	Yuan B, Li W, Liu H, Cai X, Song S, Zhao J, et al. Correlation between immune response and self-reported depression during convalescence from COVID-19. Brain Behav Immun. 2020;(bbi, 8800478).

186.	Zarghami A, Farjam M, Fakhraei B, Hashemzadeh K, Yazdanpanah MH. A Report of the Telepsychiatric Evaluation of SARS-CoV-2 Patients. Telemed J E-Health Off J Am Telemed Assoc. 2020;(dyh, 100959949).

187.	Zayet S, Kadiane-Oussou NJ, Lepiller Q, Zahra H, Royer P-Y, Toko L, et al. Clinical features of COVID-19 and influenza: a comparative study on Nord Franche-Comte cluster. Microbes Infect. 2020;(dj1, 100883508).

188.	Zayet S, Klopfenstein T, Mercier J, Kadiane-Oussou NJ, Lan Cheong Wah L, Royer P-Y, et al. Contribution of anosmia and dysgeusia for diagnostic of COVID-19 in outpatients. Infection. 2020;

189.	Zhang G., Hu C., Luo L., Fang F., Chen Y., Li J., et al. Clinical features and short-term outcomes of 221 patients with COVID-19 in Wuhan, China. J Clin Virol. 2020;127:104364.

190.	Zhang H, Shang W, Liu Q, Zhang X, Zheng M, Yue M. Clinical characteristics of 194 cases of COVID-19 in Huanggang and Taian, China. Infection. 2020;

191.	Zhang J, Wang M, Zhao M, Guo S, Xu Y, Ye J, et al. The Clinical Characteristics and Prognosis Factors of Mild-Moderate Patients With COVID-19 in a Mobile Cabin Hospital: A Retrospective, Single-Center Study. Front Public Health. 2020;8(101616579):264.

192.	Zhao J., Gao H.-Y., Feng Z.-Y., Wu Q.-J. A Retrospective Analysis of the Clinical and Epidemiological Characteristics of COVID-19 Patients in Henan Provincial People’s Hospital, Zhengzhou, China. Front Med. 2020;7:286.

193.	Zhao D, Yao F, Wang L, Zheng L, Gao Y, Ye J, et al. A comparative study on the clinical features of COVID-19 pneumonia to other pneumonias. Clin Infect Dis Off Publ Infect Dis Soc Am. 2020;(a4j, 9203213).

194.	Zheng F., Tang W., Li H., Huang Y.-X., Xie Y.-L., Zhou Z.-G. Clinical characteristics of 161 cases of corona virus disease 2019 (COVID-19) in Changsha. Eur Rev Med Pharmacol Sci. 2020;24(6):3404–10.

195.	Zheng Y., Sun L.-J., Xu M., Pan J., Zhang Y.-T., Fang X.-L., et al. Clinical characteristics of 34 COVID-19 patients admitted to intensive care unit in Hangzhou, China. J Zhejiang Univ Sci B. 2020;21(5):378–87.

196.	Zhou H., Lu S., Chen J., Wei N., Wang D., Lyu H., et al. The landscape of cognitive function in recovered COVID-19 patients. J Psychiatr Res. 2020;129:98–102.

[197.	Zhou Y., Han T., Chen J., Hou C., Hua L., He S., et al. Clinical and Autoimmune Characteristics of Severe and Critical Cases with COVID-19. Clin Transl Sci [Internet]. 2020; Available from: http://onlinelibrary.wiley.com/journal/10.1111/(ISSN)1752-8062](https://www.zotero.org/google-docs/?1sevUe)

198.	Mahammedi A, Saba L, Vagal A, Leali M, Rossi A, Gaskill M, et al. Imaging in Neurological Disease of Hospitalized COVID-19 Patients: An Italian Multicenter Retrospective Observational Study. Radiology. 2020;201933.

199.	Solomon IH, Normandin E, Bhattacharyya S, Mukerji SS, Keller K, Ali AS, et al. Neuropathological Features of Covid-19. N Engl J Med. 2020;(0255562, now).

200.	Levinson R, Elbaz M, Ben-Ami R, Shasha D, Levinson T, Choshen G, et al. Time course of anosmia and dysgeusia in patients with mild SARS-CoV-2 infection. Infect Dis Lond Engl. 2020;52(8):600–2.

201.	Kotabagi P, Fortune L, Essien S, Nauta M, Yoong W. Anxiety and depression levels among pregnant women with COVID-19. Acta Obstet Gynecol Scand. 2020;99(7):953–4.

202.	Yaghi S, Ishida K, Torres J, Mac Grory B, Raz E, Humbert K, et al. SARS-CoV-2 and Stroke in a New York Healthcare System. Stroke. 2020;51(7):2002–11.

203.	Yan CH, Faraji F, Prajapati DP, Ostrander BT, DeConde AS. Self-reported olfactory loss associates with outpatient clinical course in COVID-19. Int Forum Allergy Rhinol. 2020;10(7):821–31.

204.	Tian S, Hu N, Lou J, Chen K, Kang X, Xiang Z, et al. Characteristics of COVID-19 infection in Beijing. J Infect. 2020;80(4):401–6.

205.	Helms J, Kremer S, Merdji H, Clere-Jehl R, Schenck M, Kummerlen C, et al. Neurologic Features in Severe SARS-CoV-2 Infection. N Engl J Med. 2020;382(23):2268–70.

206.	Suleyman G, Fadel RA, Malette KM, Hammond C, Abdulla H, Entz A, et al. Clinical Characteristics and Morbidity Associated With Coronavirus Disease 2019 in a Series of Patients in Metropolitan Detroit. JAMA Netw Open. 2020 Jun 16;3(6):e2012270–e2012270.

207.	Ziehr DR, Alladina J, Petri CR, Maley JH, Moskowitz A, Medoff BD, et al. Respiratory Pathophysiology of Mechanically Ventilated Patients with COVID-19: A Cohort Study. Am J Respir Crit Care Med. 2020 Jun 15;201(12):1560–4.

208.	Dogra S, Jain R, Cao M, Bilaloglu S, Zagzag D, Hochman S, et al. Hemorrhagic stroke and anticoagulation in COVID-19. J Stroke Cerebrovasc Dis. 2020 Aug;29(8):N.PAG-N.PAG.

209.	Lechien JR, Chiesa-Estomba CM, De Siati DR, Horoi M, Le Bon SD, Rodriguez A, et al. Olfactory and gustatory dysfunctions as a clinical presentation of mild-to-moderate forms of the coronavirus disease (COVID-19): a multicenter European study. Eur Arch Oto-Rhino-Laryngol Off J Eur Fed Oto-Rhino-Laryngol Soc EUFOS Affil Ger Soc Oto-Rhino-Laryngol - Head Neck Surg. 2020;277(8):2251–61.

210.	Parma V, Ohla K, Veldhuizen MG, Niv MY, Kelly CE, Bakke AJ, et al. More than smell - COVID-19 is associated with severe impairment of smell, taste, and chemesthesis. Chem Senses. 2020;(b4b, 8217190).

211.	De Maria A, Varese P, Dentone C, Barisione E, Bassetti M. High prevalence of olfactory and taste disorder during SARS-CoV-2 infection in outpatients. J Med Virol. 2020;(i9n, 7705876).

212.	Liguori C, Pierantozzi M, Spanetta M, Sarmati L, Cesta N, Iannetta M, et al. Subjective neurological symptoms frequently occur in patients with SARS-CoV2 infection. Brain Behav Immun. 2020;(bbi, 8800478).

213.	Wee LE, Chan YFZ, Teo NWY, Cherng BPZ, Thien SY, Wong HM, et al. The role of self-reported olfactory and gustatory dysfunction as a screening criterion for suspected COVID-19. Eur Arch Oto-Rhino-Laryngol Off J Eur Fed Oto-Rhino-Laryngol Soc EUFOS Affil Ger Soc Oto-Rhino-Laryngol - Head Neck Surg. 2020;277(8):2389–90.

214.	Giacomelli A, Pezzati L, Conti F, Bernacchia D, Siano M, Oreni L, et al. Self-reported olfactory and taste disorders in SARS-CoV-2 patients: a cross-sectional study. Clin Infect Dis Off Publ Infect Dis Soc Am. 2020;(a4j, 9203213).

215.	Gelardi M, Trecca E, Cassano M, Ciprandi G. Smell and taste dysfunction during the COVID-19 outbreak: a preliminary report. Acta Bio-Medica Atenei Parm. 2020;91(2):230–1.
