## supplementary table 4 for "The neurology and neuropsychiatry of COVID-19: a systematic review and meta-analysis of the early literature reveals frequent CNS manifestations and key emerging narratives"

**Supplementary Table 4: Comparison of proportion estimation models**

|  | GLMM | | | Double-arcsine transformation | | |
| --- | --- | --- | --- | --- | --- | --- |
| Symptom/Syndrome | Prevalence (%) | 95% CI | *I*^2^ | Prevalence (%) | 95% CI | *I*^2^ |
| Headache | 20.7 | 16.1 - 26.1 | 99.0% | 24.3 | 19.6 - 29.4 | 99.2% |
| Myalgia | 25.1 | 19.8-31.3 | 99.1% | 28.6 | 23.5 - 33.9 | 99.3% |
| Anosmia | 43.1 | 35.2 – 51.3 | 98.8% | 45.0 | 37.8 - 51.5 | 98.6% |
| Fatigue | 35.7 | 29.2 – 42.9 | 98.4% | 37.1 | 30.9 - 43.5 | 98.4% |
| Dysgeusia | 37.2 | 29.8 – 45.3 | 98.6% | 39.5 | 32.8 - 46.5 | 98.5% |
