## supplementary table 5 for "The neurology and neuropsychiatry of COVID-19: a systematic review and meta-analysis of the early literature reveals frequent CNS manifestations and key emerging narratives"

**Supplementary table 5: Subgroup analysis by country of origin**

| Outcome | Country (*N* studies) | Prevalence % | 95% CI | *p* |
| --- | --- | --- | --- | --- |
| Headache (84 studies) | China (32) | 7.8 | 5.7-10.4 | (reference) |
|  | Italy (12) | 28.0 | 19.4-38.6 | <0.001 |
|  | USA (12) | 32.6 | 18.4-51.0 | <0.001 |
| Myalgia (76 studies) | China (27) | 10.4 | 7.1-15.1 | (reference) |
|  | USA (13) | 34.0 | 19.1-52.9 | 0.001 |
|  | Italy (10) | 26.4 | 18.7-35.9 | <0.001 |
| Fatigue (67 studies) | China (30) | 31.2 | 24.5-38.8 | (reference) |
|  | Italy (11) | 31.1 | 18.9-46.8 | 1.00 |
|  | USA (11) | 64.2 | 56.7-70.9 | <0.001 |
| Anosmia (63 studies) | Italy (15) | 41.1 | 26.7-57.1 | (reference) |
|  | France (9) | 35.5 | 18.4-57.4 | 0.67 |
|  | USA (8) | 27.2 | 12.1-50.5 | 0.31 |
| Dysgeusia (52 studies) | Italy (13) | 39.1 | 26.0-54.1 | (reference) |
|  | France (6) | 39.1 | 21.3-60.4 | 1.00 |
|  | USA (6) | 45.0 | 27.8-63.4 | 0.63 |
