## Supplementary figures and images for "The neurology and neuropsychiatry of COVID-19: a systematic review and meta-analysis of the early literature reveals frequent CNS manifestations and key emerging narratives"

### supplementary figures

#### Proportion of patients reporting headache in SARS−CoV−2 infection

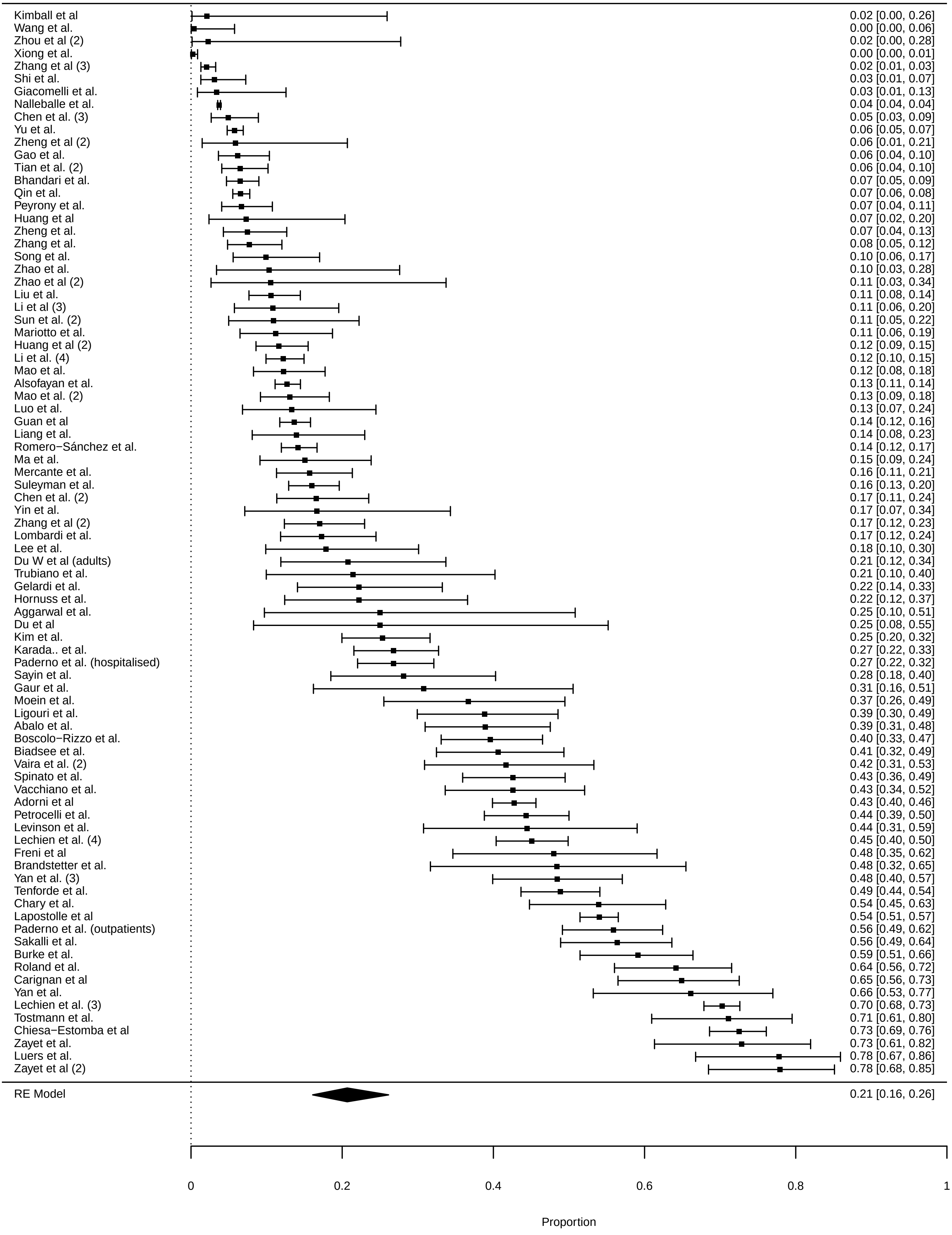

### supplementary figures

#### Proportion of patients reporting myalgia in SARS−CoV−2 infection

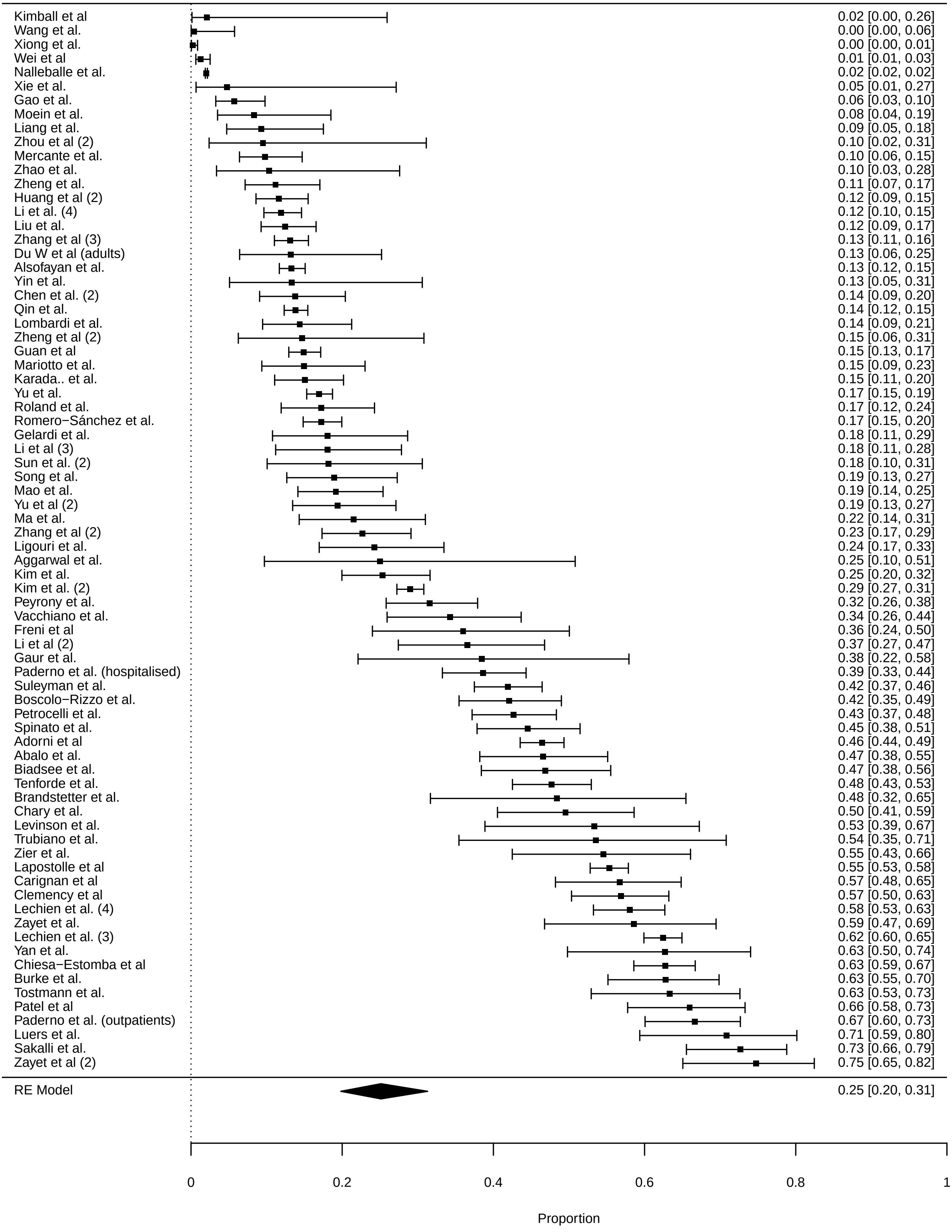

### supplementary figures

#### Proportion of patients reporting anosmia in SARS−CoV−2 infection

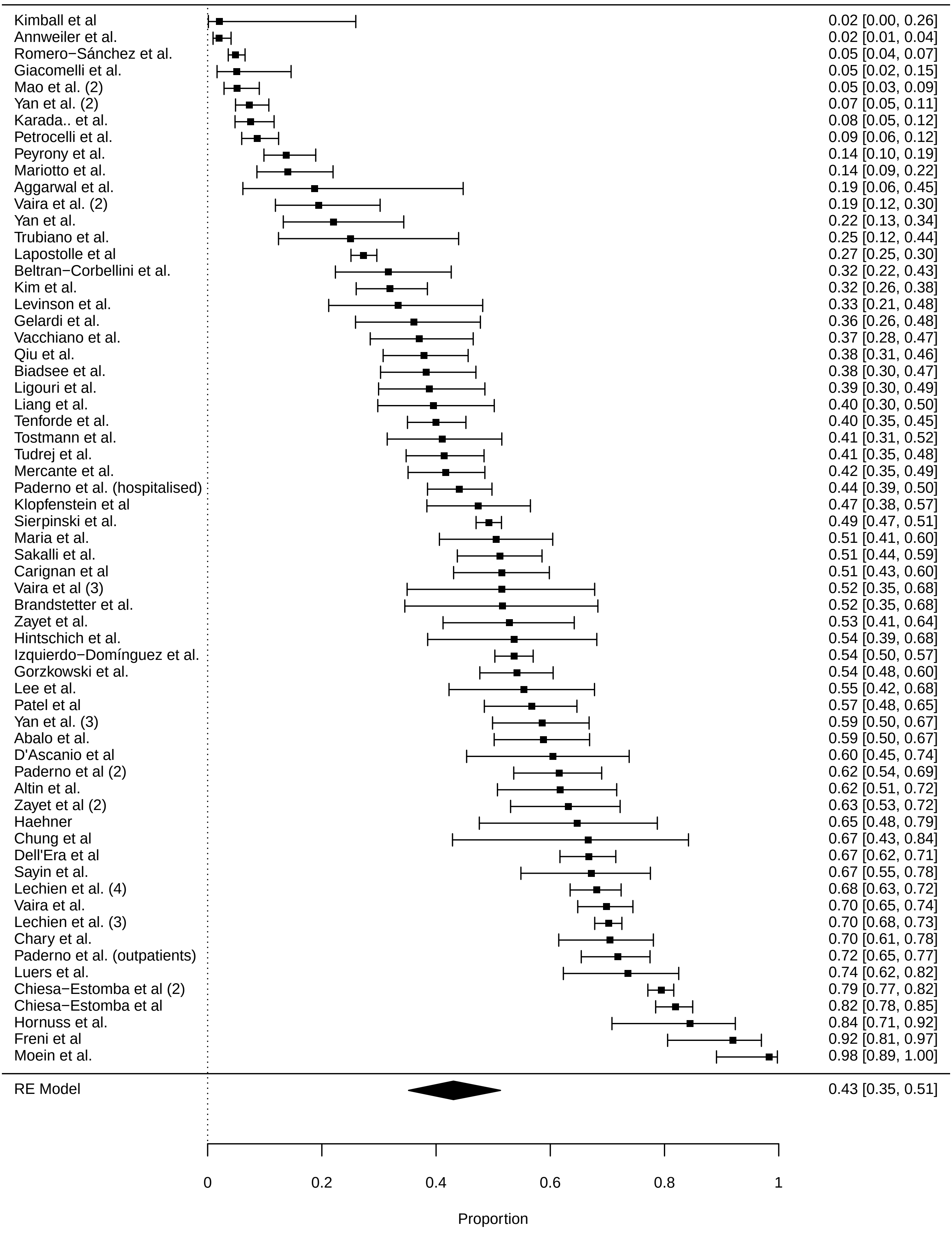

### supplementary figures

#### Proportion of patients reporting fatigue in SARS−CoV−2 infection

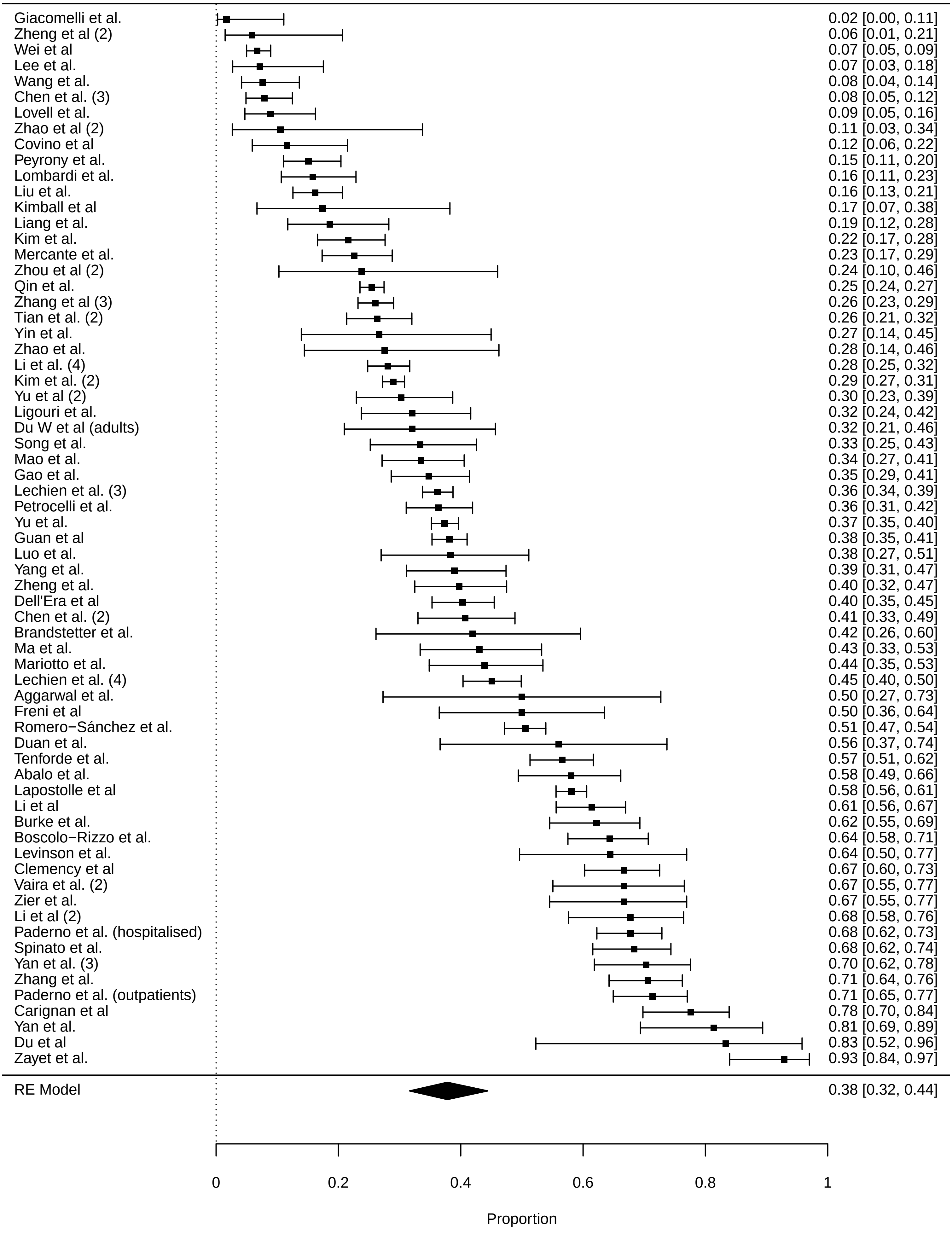

### supplementary figures

#### Proportion of patients reporting dysguesia in SARS−CoV−2 infection

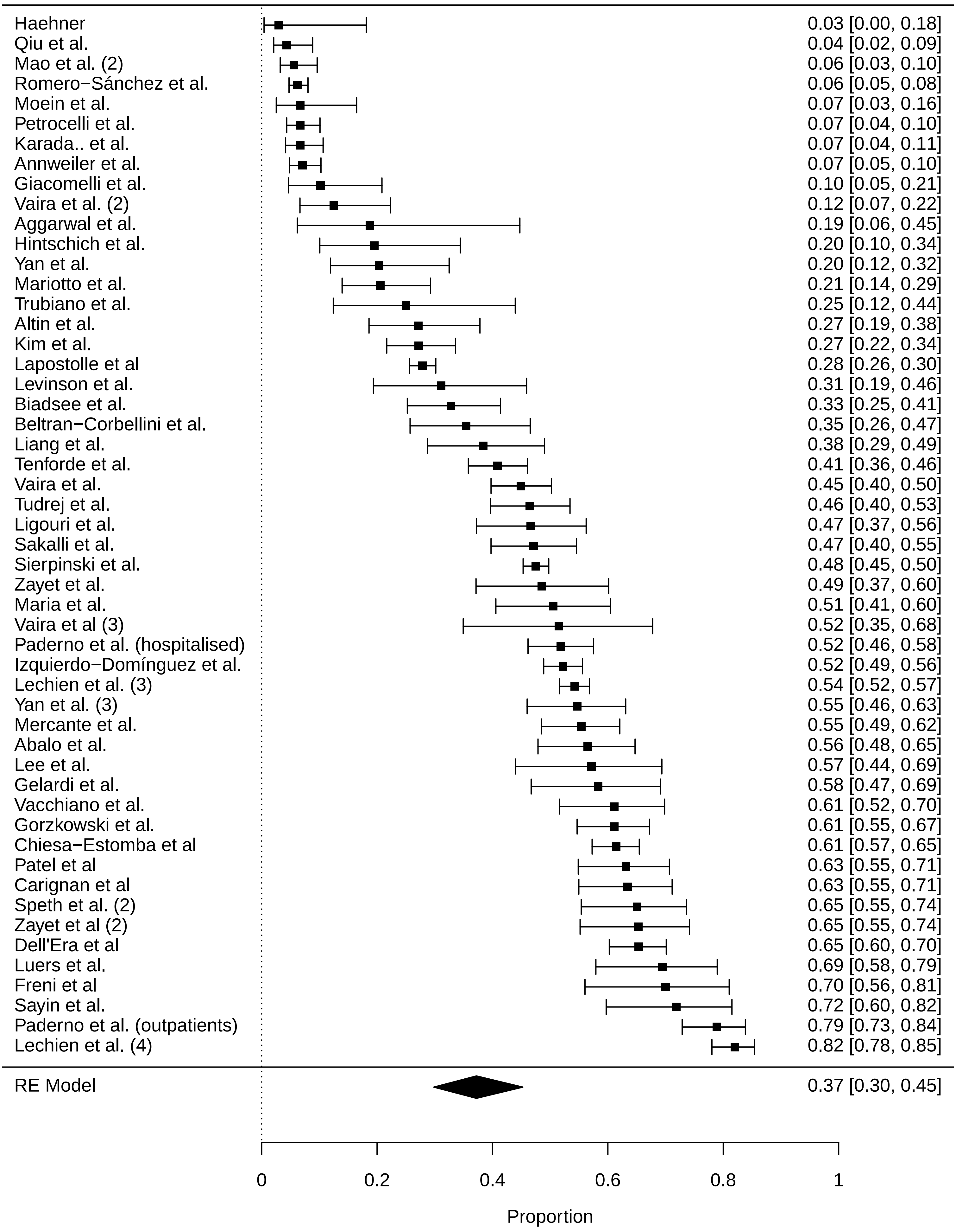

### supplementary figures

##### Proportion of patients reporting dizziness/vertigo in SARS−CoV−2 infection

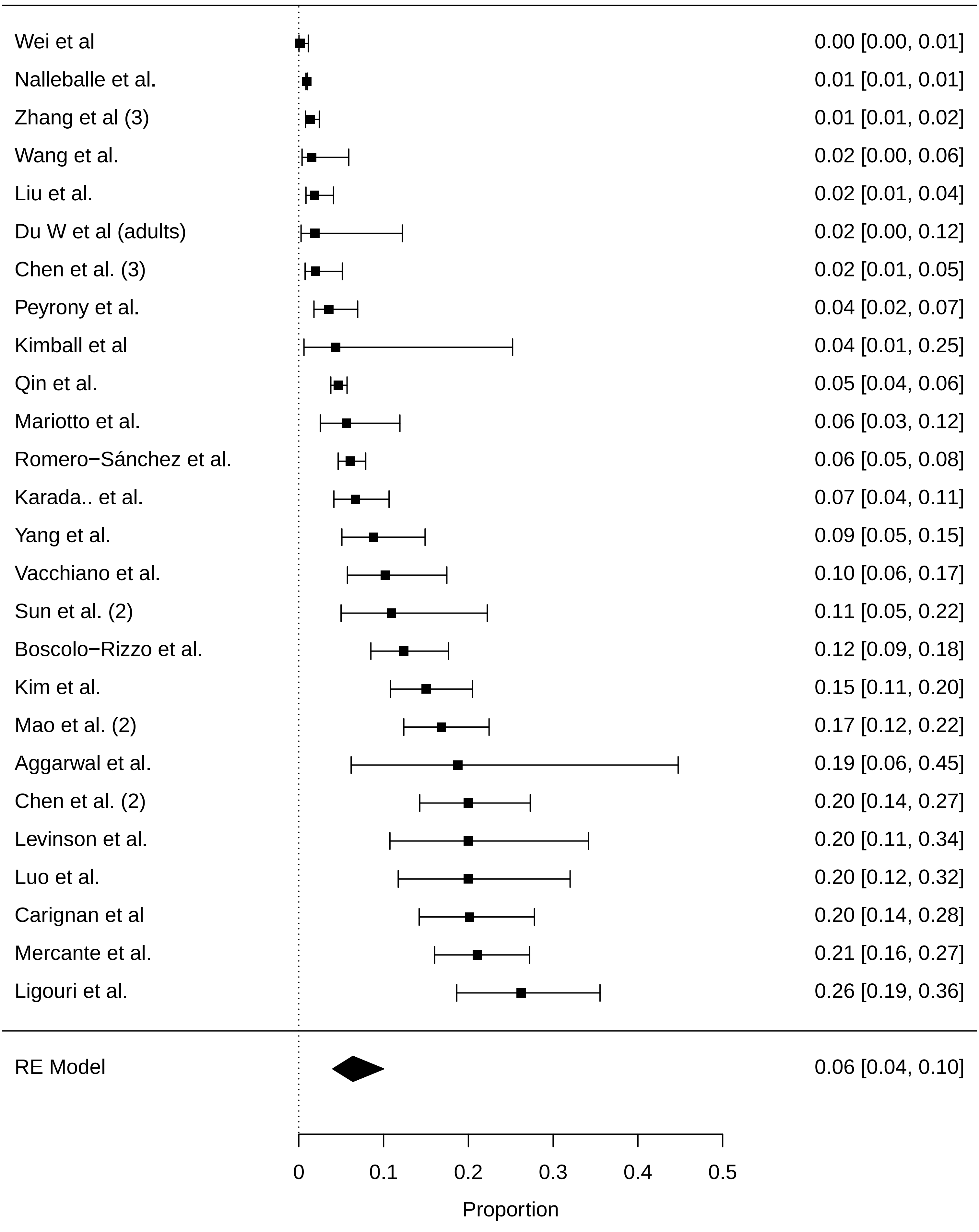

### supplementary figures

##### Proportion of patients reporting altered mental status in SARS−CoV−2 infection

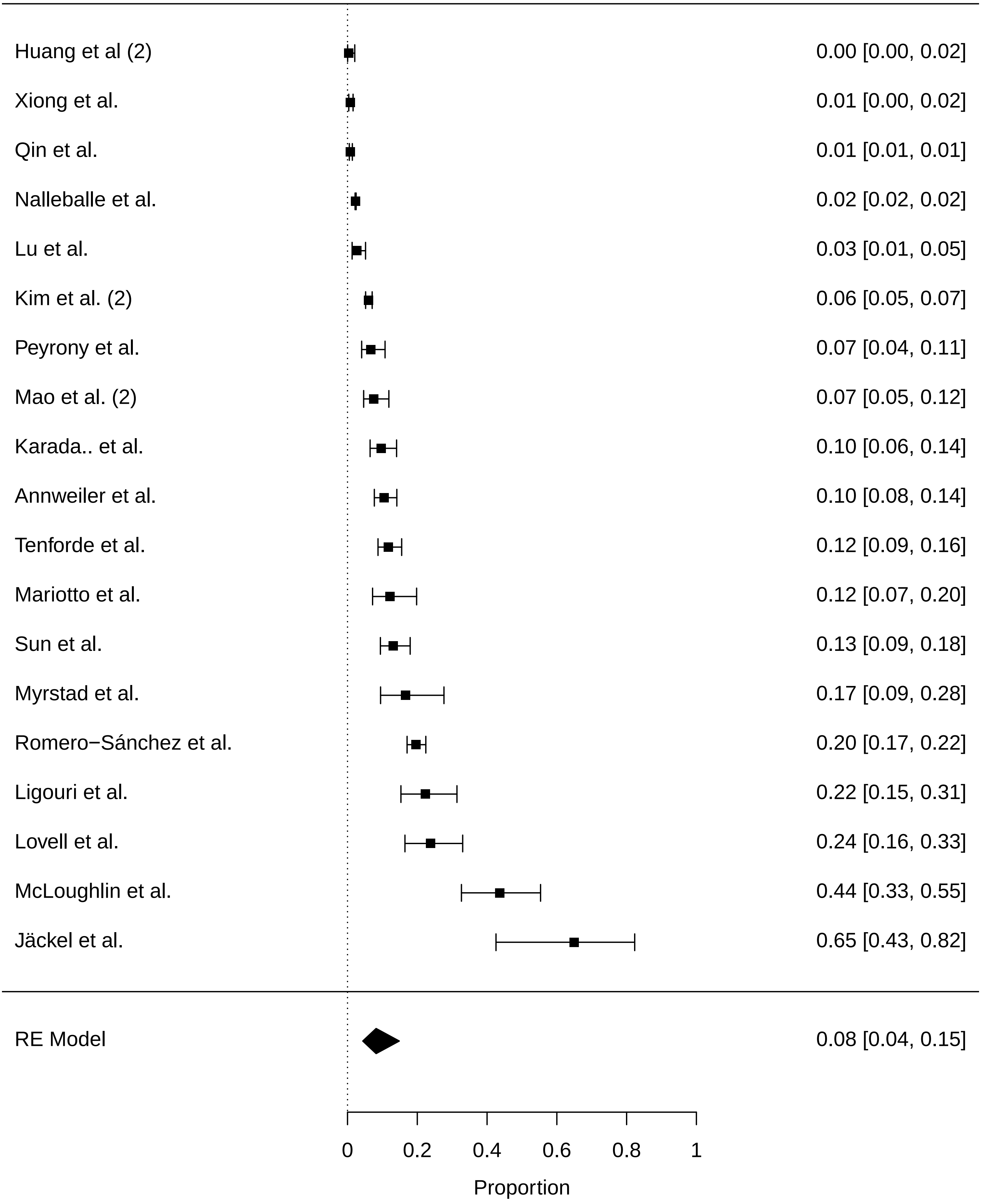

### supplementary figures

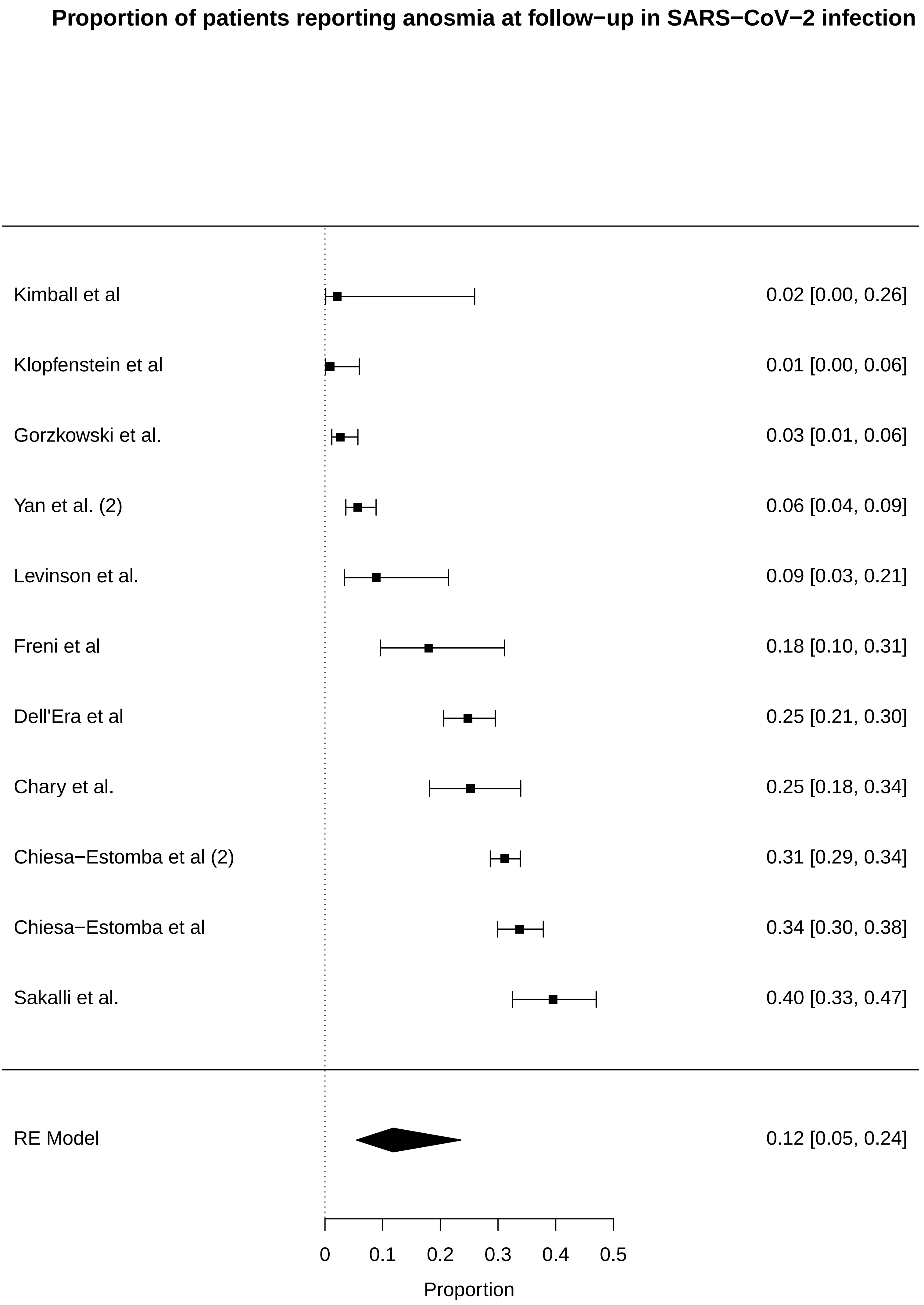

### supplementary figures

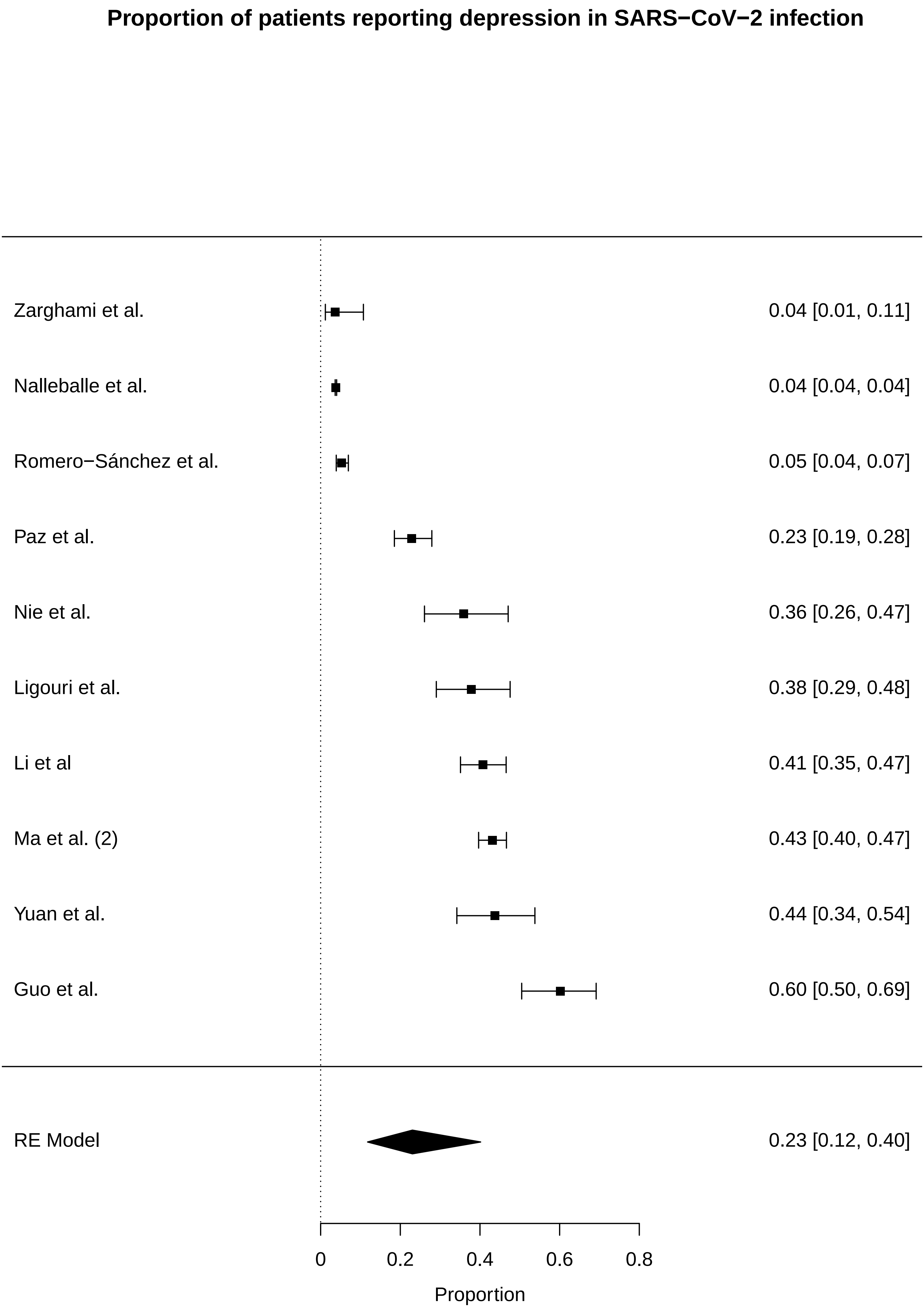

### supplementary figures

##### Proportion of patients reporting anxiety in SARS−CoV−2 infection

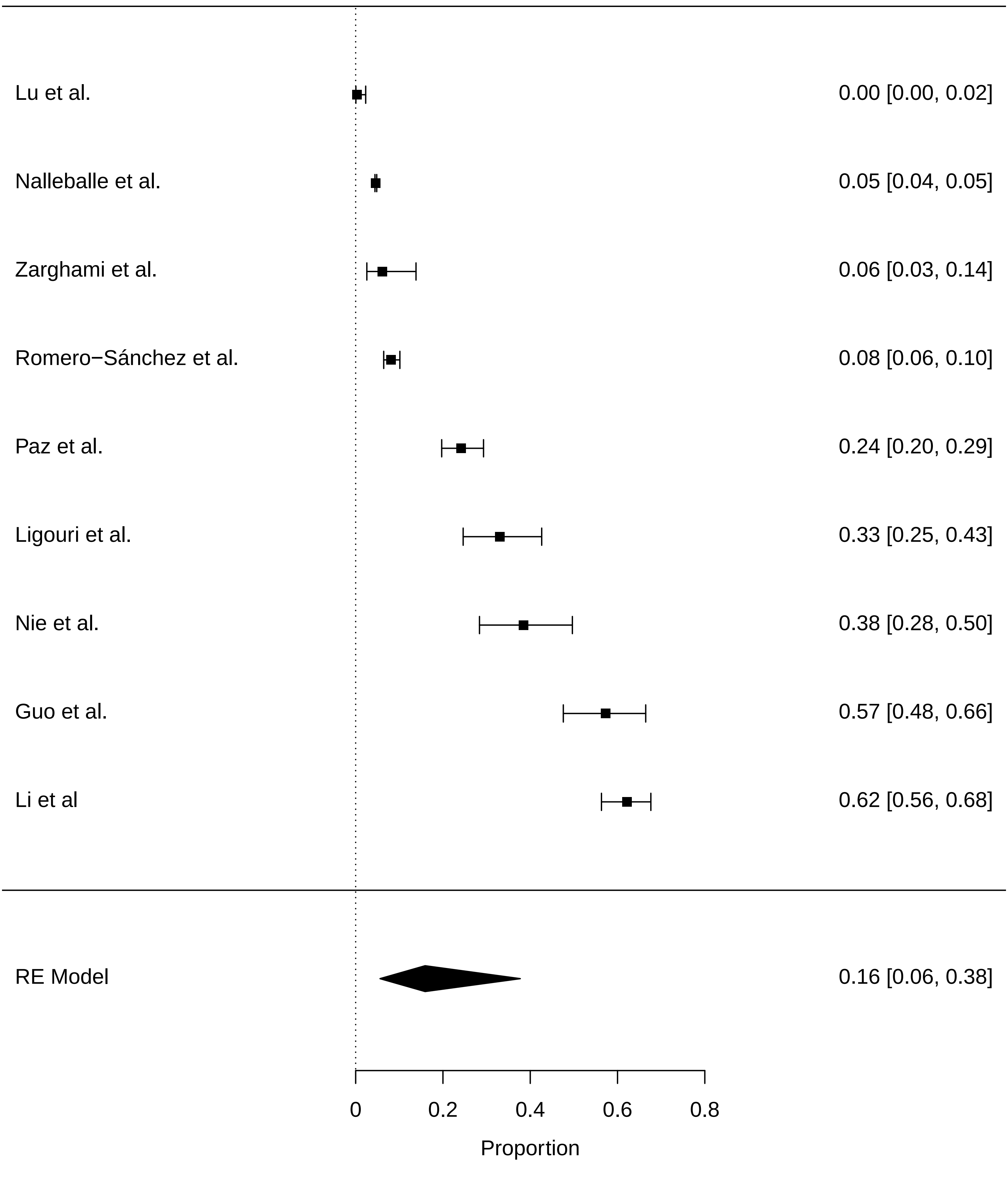

### supplementary figures

**Proportion of patients reporting sleep disorder in SARS−CoV−2 infection**

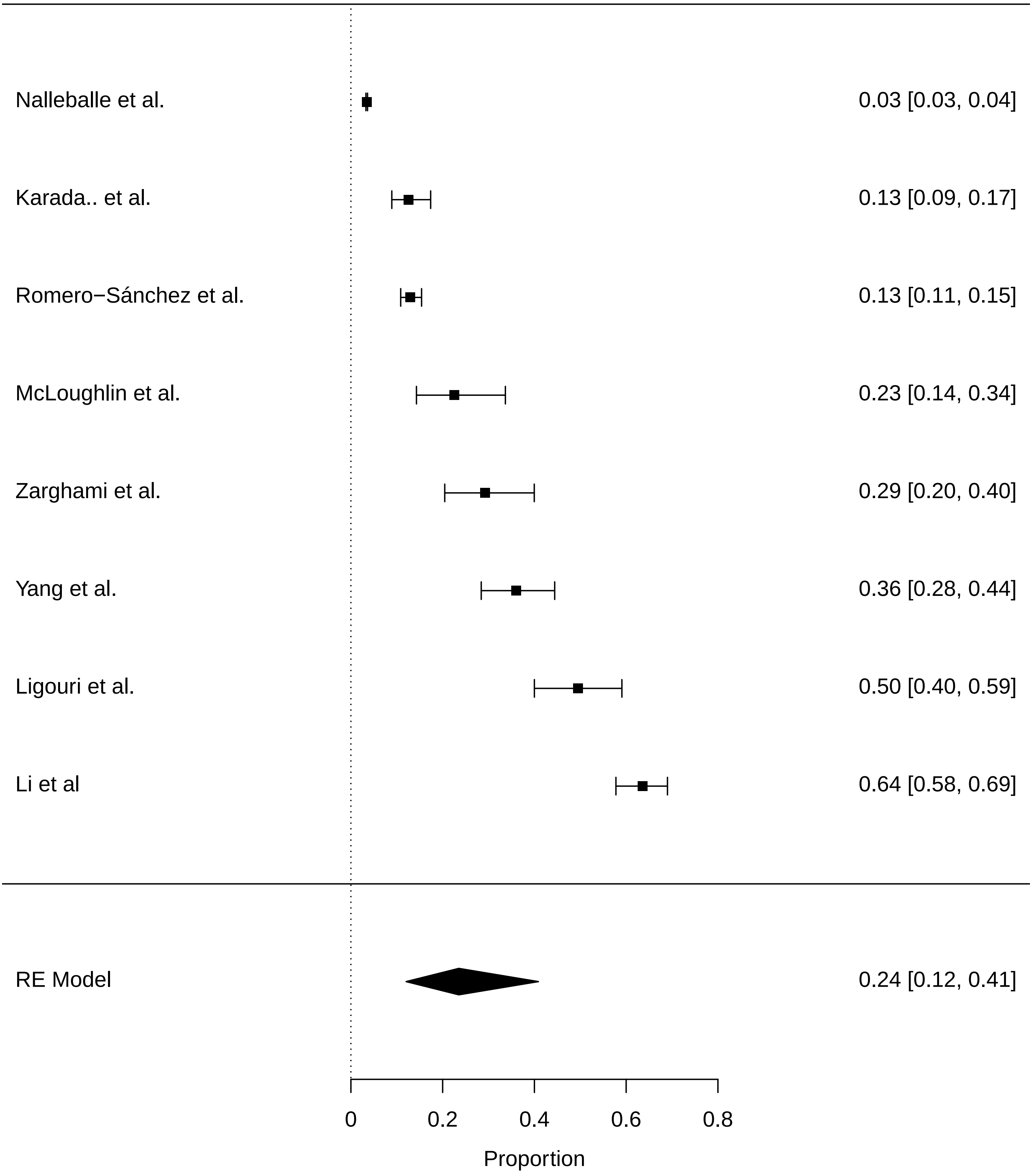

### supplementary figures

#### Proportion of patients with ischaemic stroke in SARS−CoV−2 infection

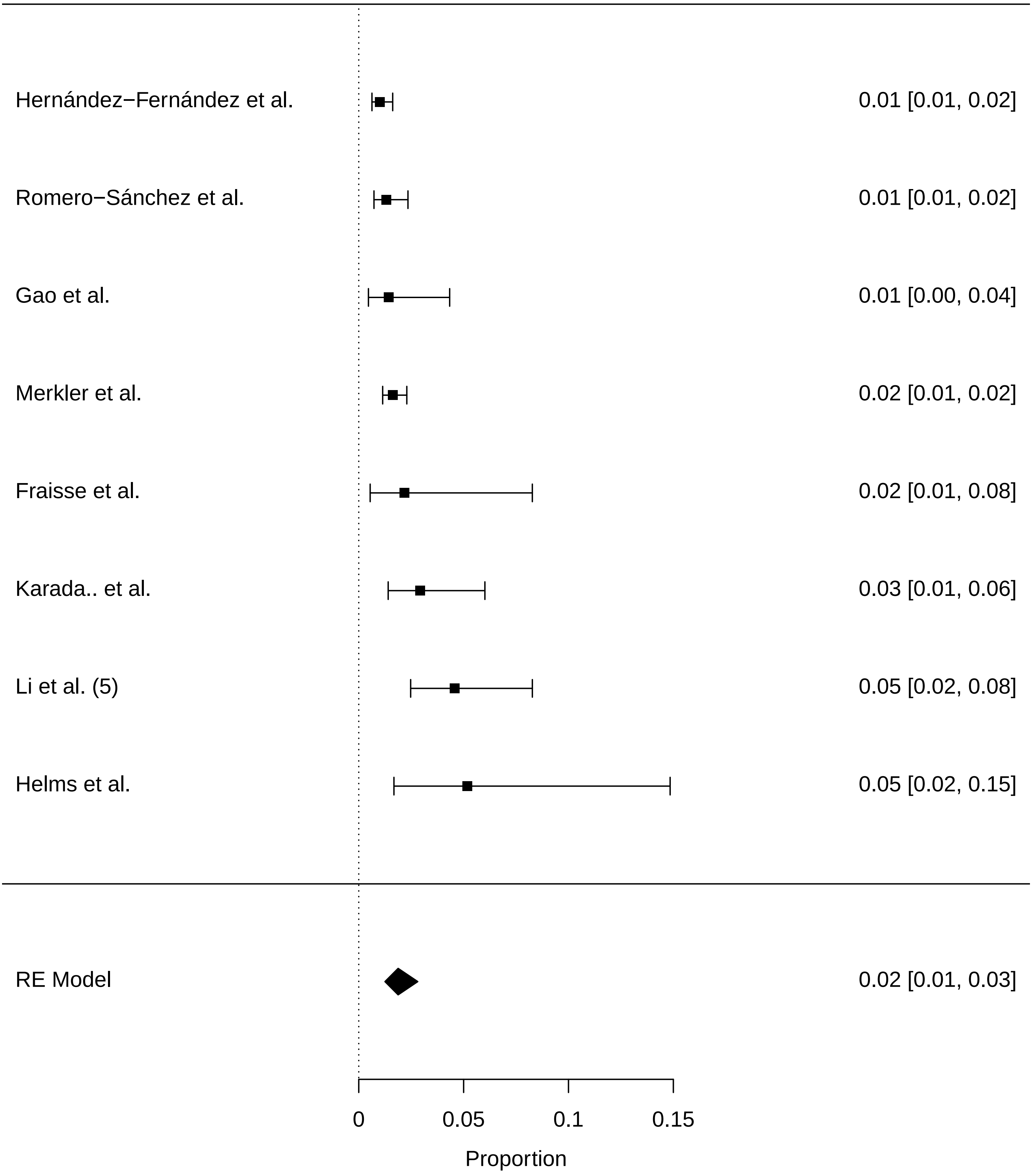

### supplementary figures

##### roportion of patients with unsecified cerebrovascular accident stroke in SARS−CoV−2 inf

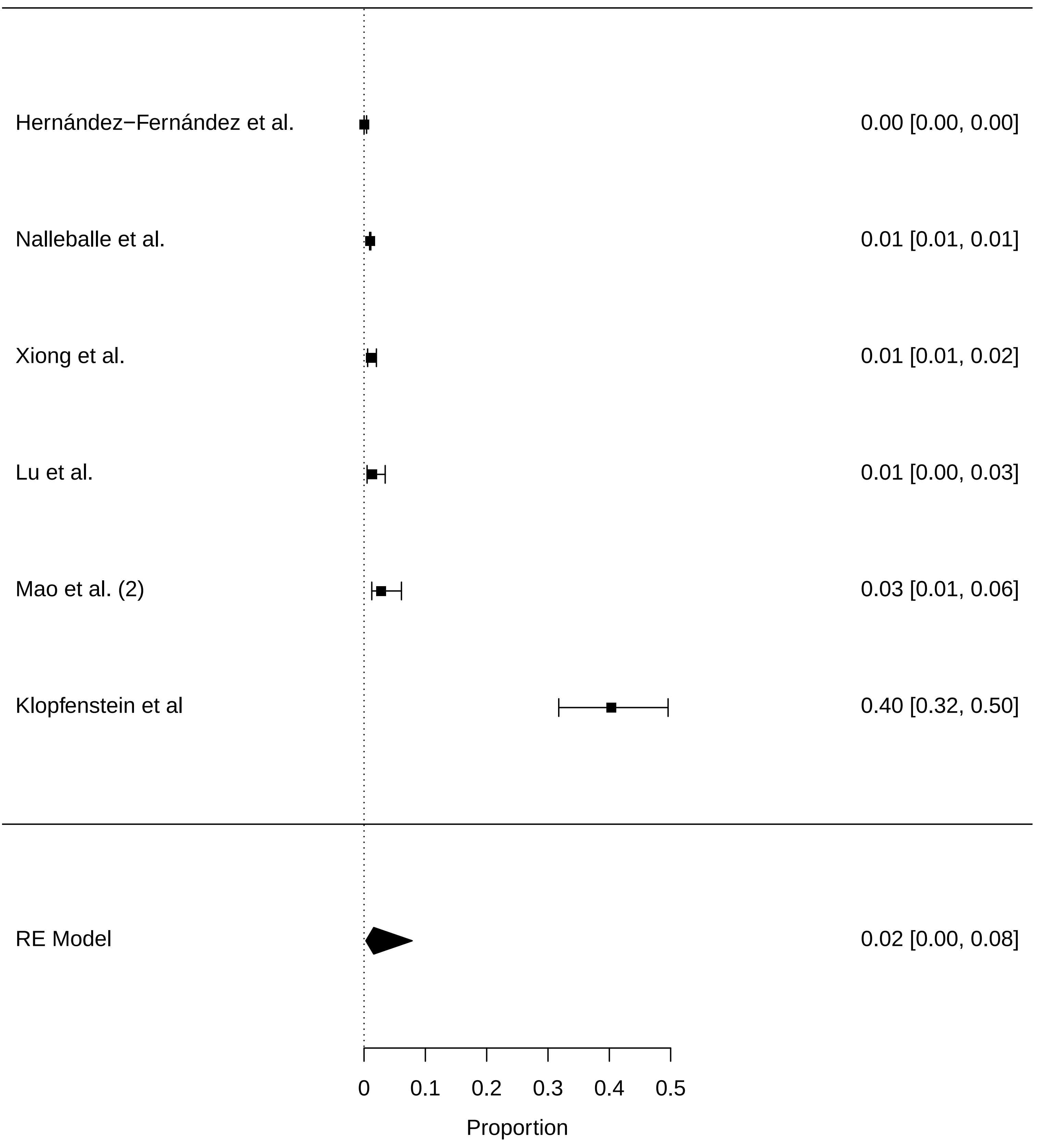

### supplementary figures

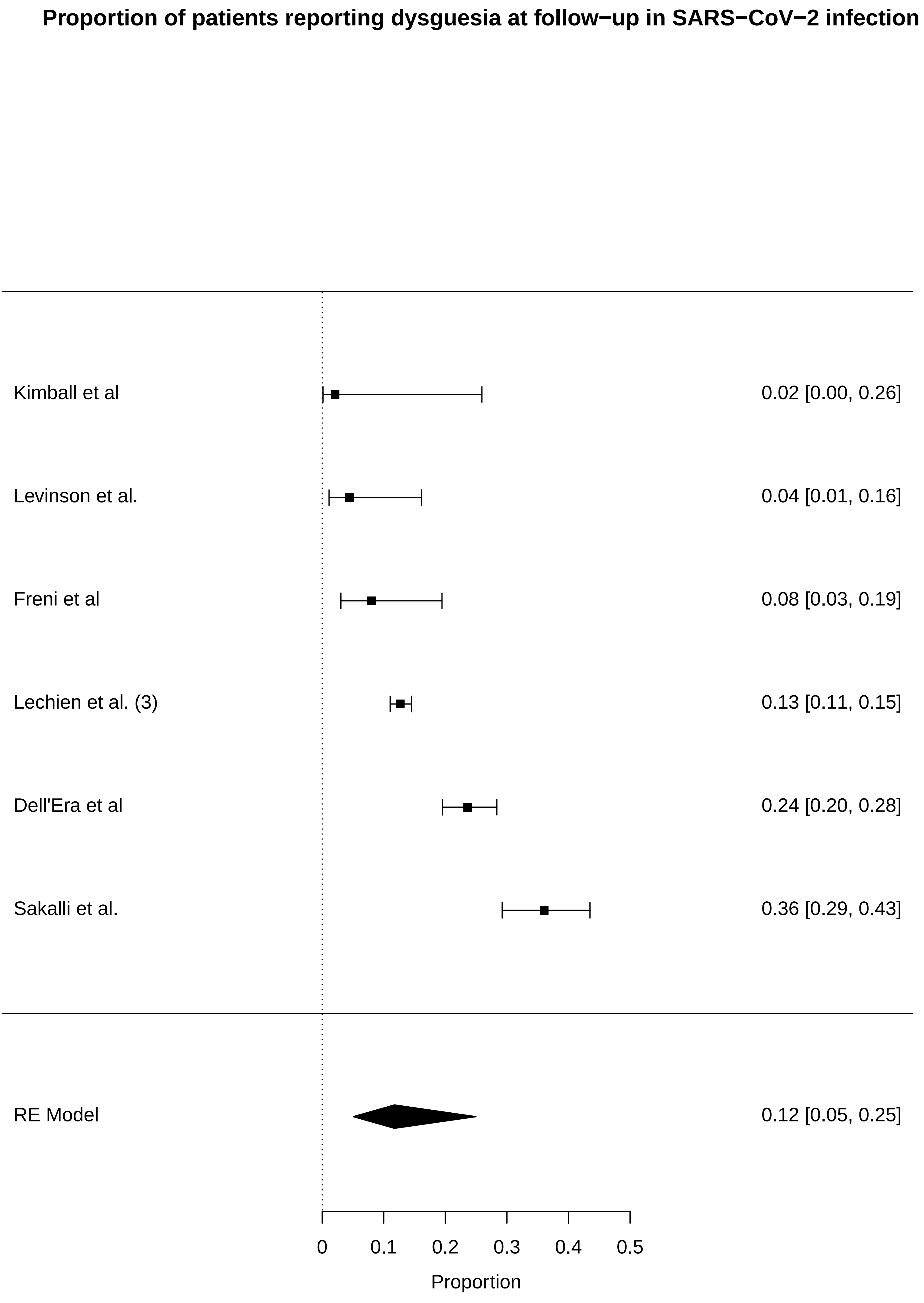

### supplementary figures

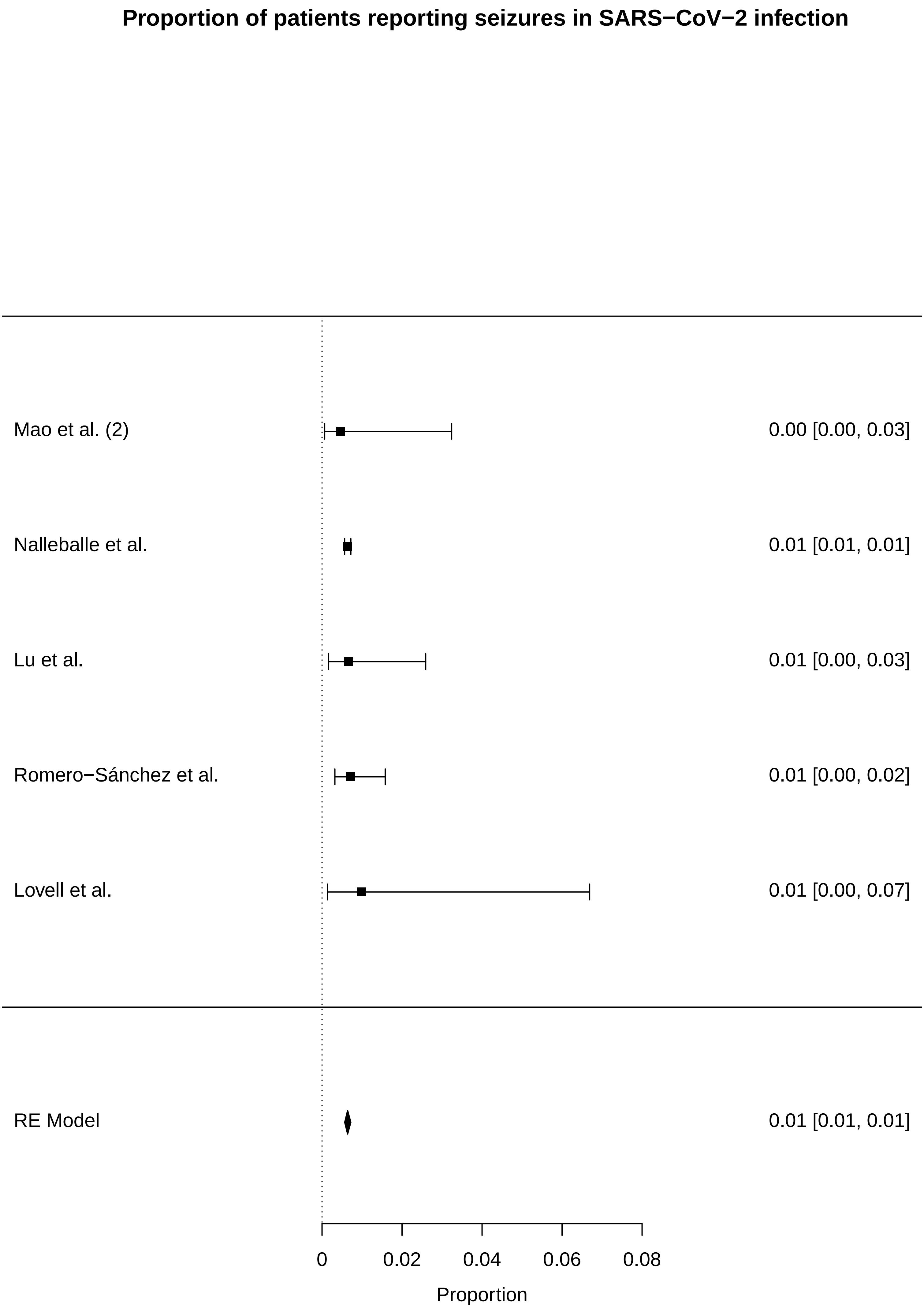

### supplementary figures

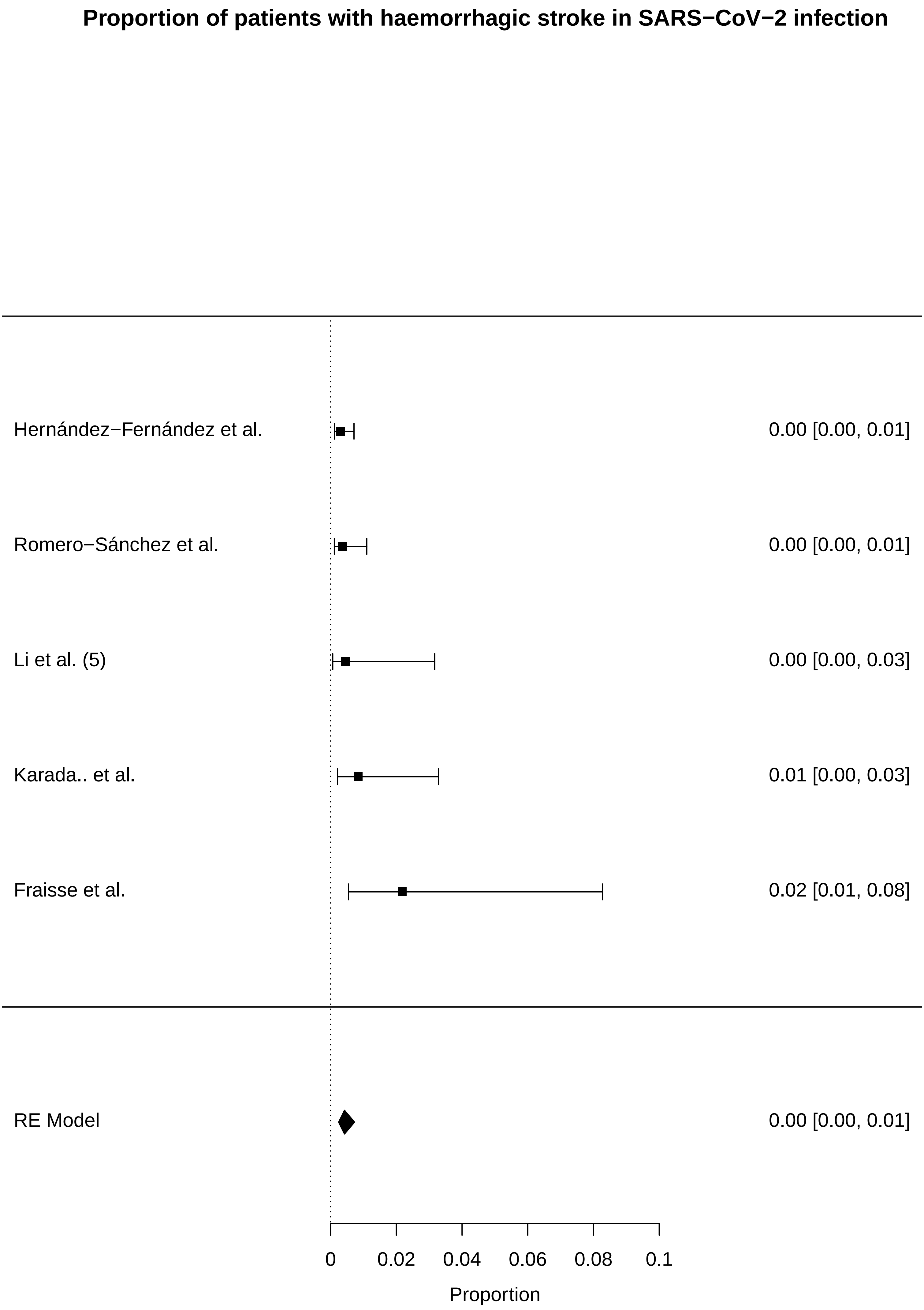

### supplementary figures

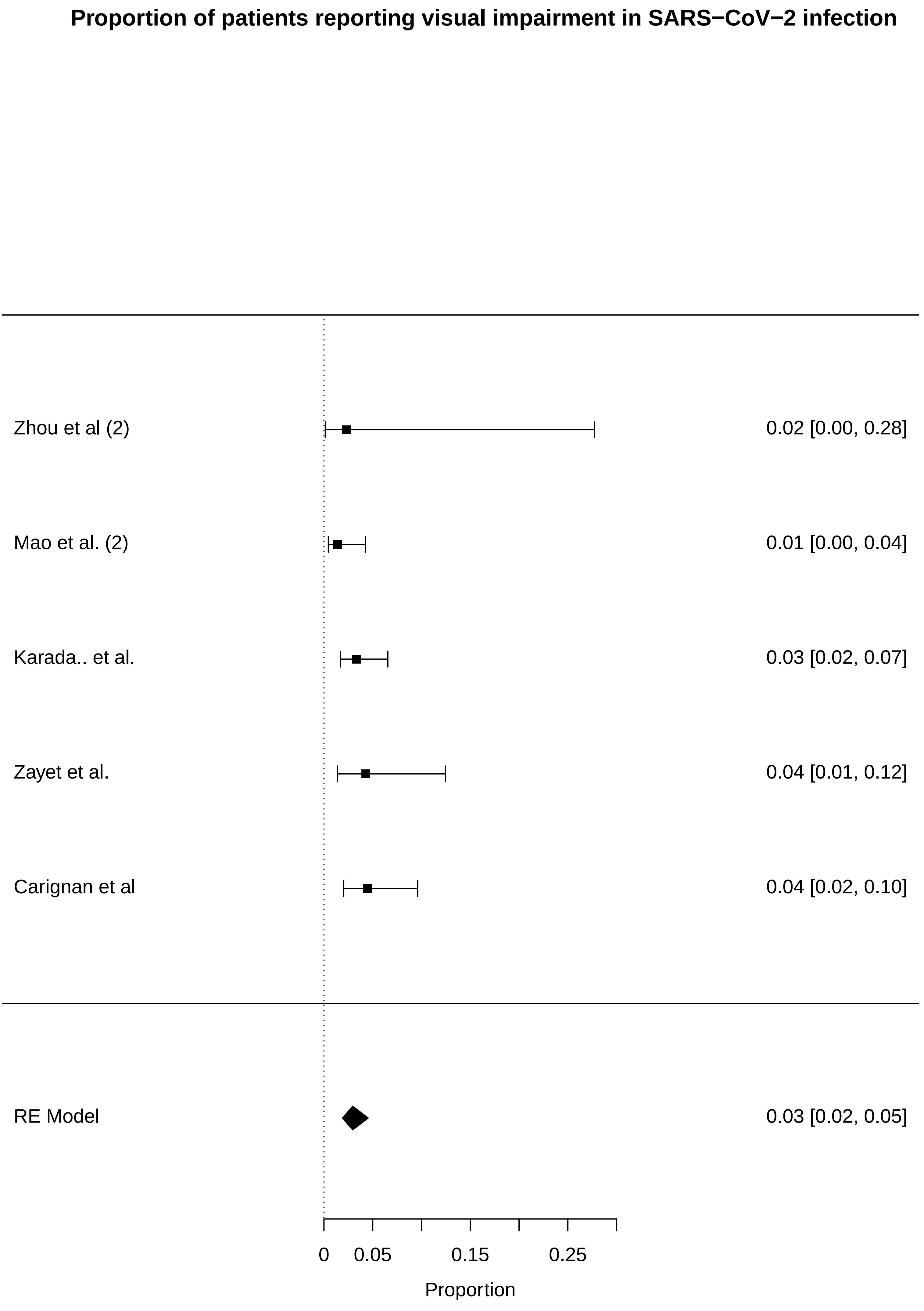

### supplementary figures

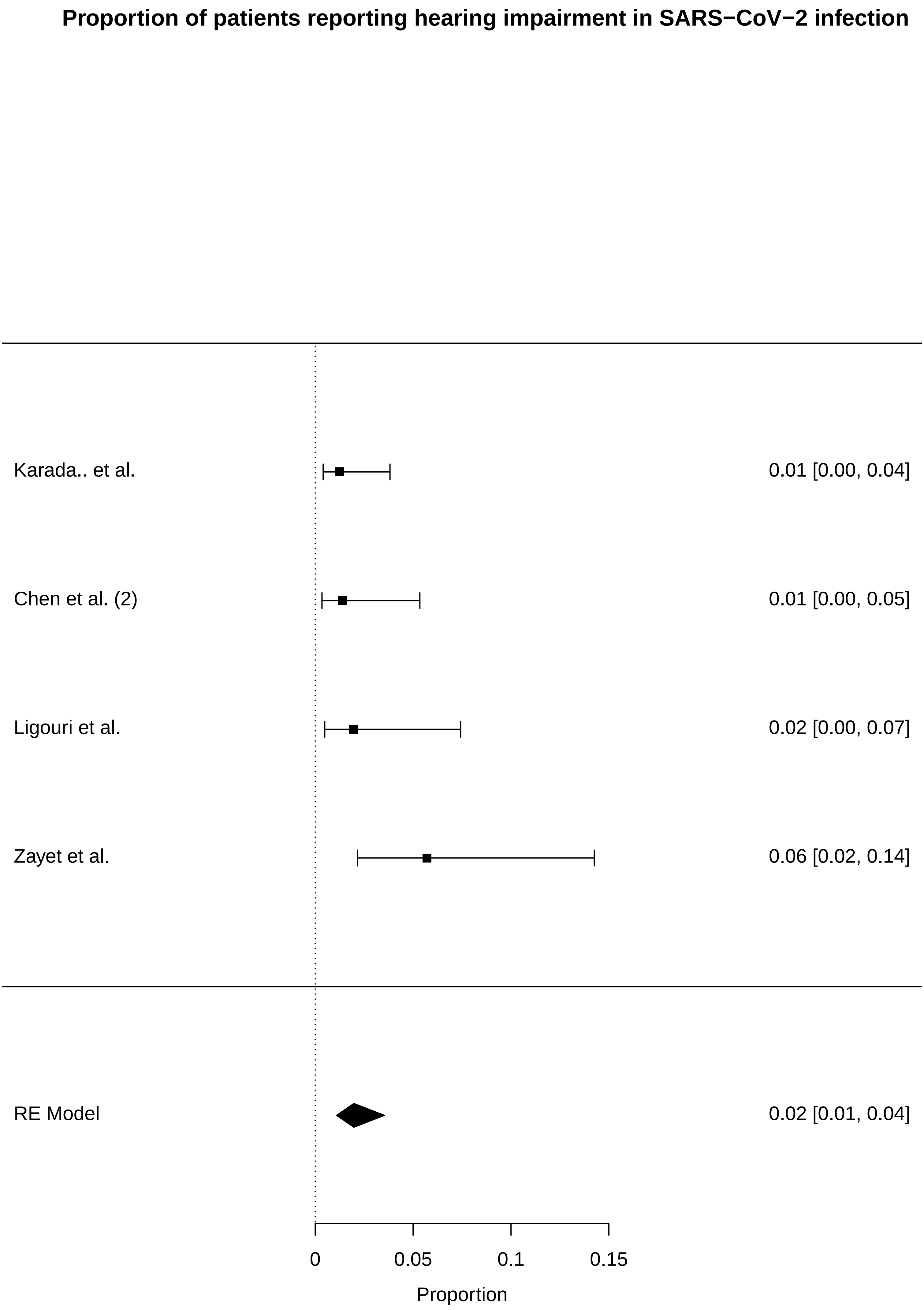

### supplementary figures

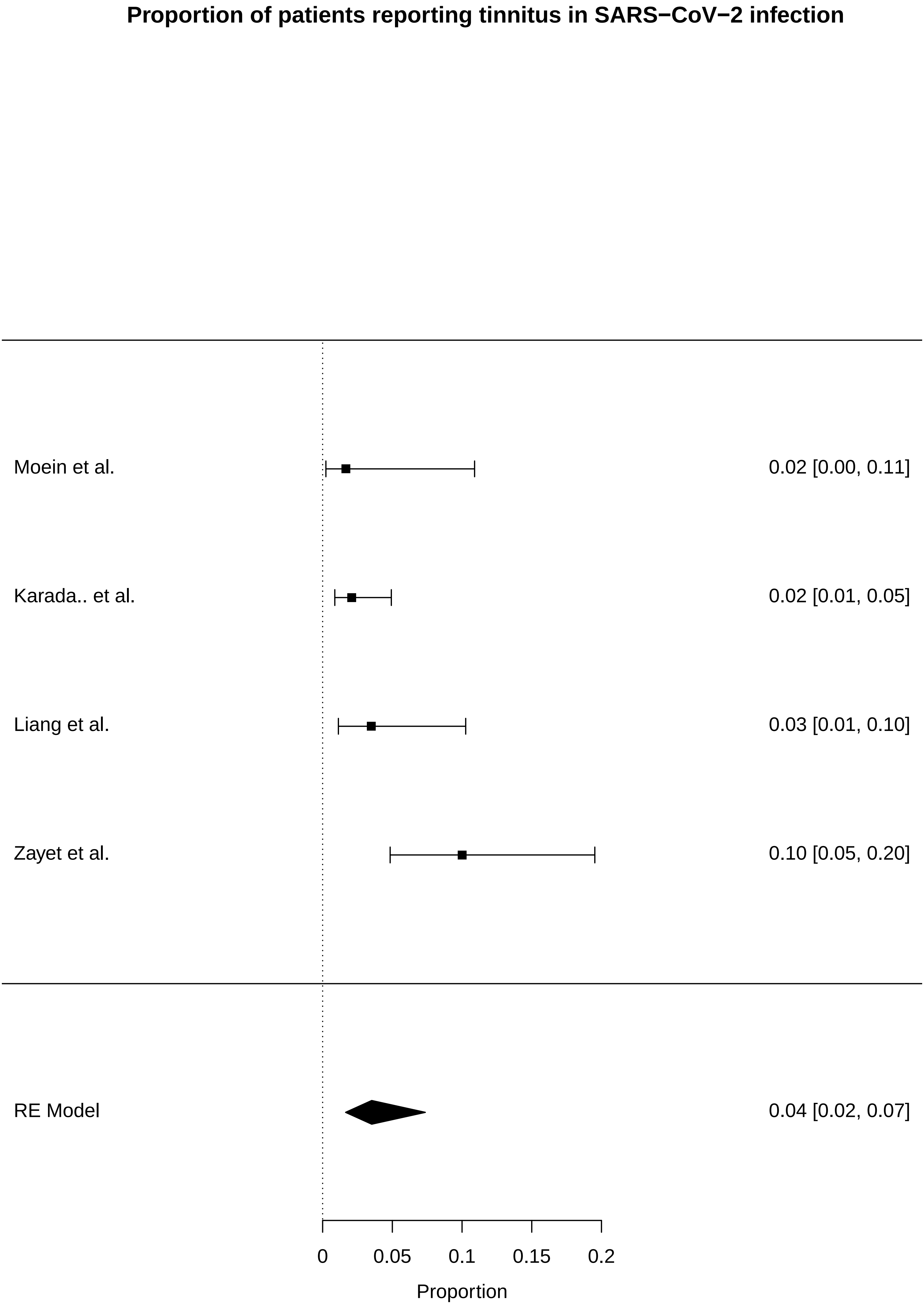

### supplementary figures

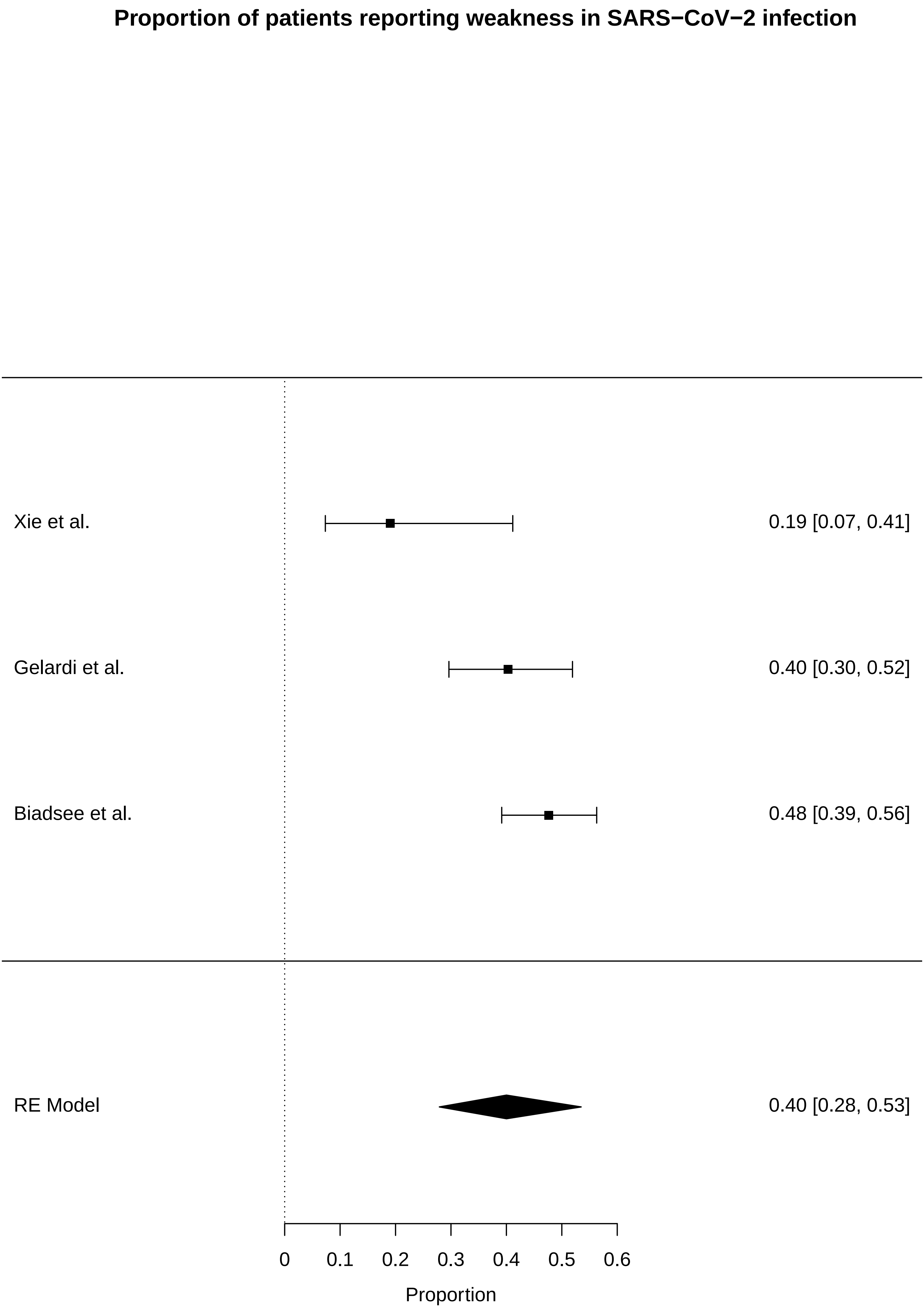
